# Single cell sequencing analysis uncovers genetics-influenced CD16+monocytes and memory CD8+T cells involved in severe COVID-19

**DOI:** 10.1101/2022.02.06.21266924

**Authors:** Yunlong Ma, Fei Qiu, Chunyu Deng, Jingjing Li, Yukuan Huang, Zeyi Wu, Yijun Zhou, Yaru Zhang, Yichun Xiong, Yinghao Yao, Yigang Zhong, Jia Qu, Jianzhong Su

## Abstract

**Background:** Understanding the host genetic architecture and viral immunity contributes to the development of effective vaccines and therapeutics for controlling the COVID-19 pandemic. Alterations of immune responses in peripheral blood mononuclear cells play a crucial role in the detrimental progression of COVID-19. However, the effects of host genetic factors on immune responses for severe COVID-19 remain largely unknown.

**Methods:** We constructed a powerful computational framework to characterize the host genetics-influenced immune cell subpopulations for severe COVID-19 by integrating GWAS summary statistics (N = 969,689 samples) with four independent scRNA-seq datasets (N = 606,534 cells).

**Results:** We found that 34 risk genes were significantly associated with severe COVID-19, and the number of highly-expressed genetics-risk genes increased with the severity of COVID-19. Three cell-subtypes that are CD16+monocytes, megakaryocytes, and memory CD8+T cells were significantly enriched by COVID-19-related genetic association signals. Notably, three causal risk genes of *CCR1, CXCR6*, and *ABO* were specifically expressed in these three cell types, respectively. *CCR1*^+^CD16+monocytes and *ABO*^+^ megakaryocytes with significant up-regulated genes including *S100A12, S100A8, S100A9*, and *IFITM1* confer higher risk to the cytokine storms among severe patients. *CXCR6*^+^ memory CD8+ T cells exhibit a notable polyfunctionality of multiple immunologic features, including elevation of proliferation, migration, and chemotaxis. Moreover, we observed a prominent increase in cell-cell interactions of both *CCR1*^+^ CD16+monocytes and *CXCR6*^+^ memory CD8+T cells in severe patients compared to normal controls among both PBMCs and lung tissues, and elevated interactions with epithelial cells could contribute to enhance the resident to lung airway for against COVID-19 infection.

**Conclusions:** We uncover a major genetics-modulated immunological shift between mild and severe infection, including an increase in up-regulated genetic-risk genes, excessive secreted inflammatory cytokines, and functional immune cell subsets contributing high risk to severity, which provides novel insights in parsing the host genetics-influenced immune cells for severe COVID-19.

## Background

The coronavirus disease 2019 (COVID-19) outbreak, caused by severe acute respiratory syndrome coronavirus 2 (SARS-CoV-2), has widely and severely jeopardized the health and economy systems of most countries worldwide. As of July 21^th^, 2021, there were more than 192.2 million confirmed patients with more than 4.12 million deaths in the whole world [1]. COVID-19 has distinct clinical manifestations ranging from asymptomatic to severe respiratory failure [2]. Mortalities of COVID-19 are largely derived from severe patients with interstitial pneumonia in both lungs and acute respiratory distress syndrome [3]. Many earlier studies [4-6] have shown that the number of severe COVID-19 patients who are elders and have comorbidities, such as diabetes and hypertension, has increased. In this connection, understanding the immunologic mechanism of severe COVID-19 and identifying novel vaccine targets to control the pandemic are of considerable interest.

Accumulating evidence have suggested that alterations of immune responses in peripheral blood mononuclear cells (PBMCs) and bronchoalveolar lavage fluid (BALF) play a crucial role in the detrimental progression of COVID-19 [7, 8]. There has been evidence that cytokine storm, usually found in severe COVID-19 patients, causes the adverse progression of COVID-19 [7]. Increased circulating levels of proinflammatory cytokine, including IL-10, IL-6 and TNF-α, have been reported to be associated with severe COVID-19 [7, 9]. Single-cell RNA sequencing (scRNA-seq) has been extensively utilized to reveal the immune responses of COVID-19 patients in both lung and peripheral blood [10-18]. Megakaryocytes and monocytes [11, 12], T cells exhaustion [14], lymphopenia [19], and increased levels of cytokines [20] may cause aberrant peripheral immune activities in severe COVID-19 patients. Based on large-scale samples, previous studies identified that dysregulation of mTOR signaling pathway in dendritic cells [21] and aberrant myeloid cell subpopulations [16, 17] implicated in severe COVID-19. Su et al. [10] revealed an increase in inflammation and a sharp drop in blood nutrients between mild and moderate-to-severe COVID-19, and new subsets of immune cells emerged in moderate COVID-19 patients.

Genome-wide association study (GWAS) has emerged as a powerful approach to identify risk genes and genetic variants for complex diseases. By gathering population-based GWAS data worldwide, the COVID-19 Host Genetic Consortium has launched the “COVID-19 Host Genetics Initiative” project to facilitate COVID-19 host genetic research and identify genetic determinants of COVID-19 [22]. Subsequently, a growing number of GWASs have identified numerous significant genetic variants associated with COVID-19 susceptibility and severity [23-28]. Ellinghaus et al. [27] performed a meta-analysis of two independent GWAS datasets with 1,610 severe COVID-19 patients and 2,205 matched controls at seven hospitals in the Italian and Spanish epicenters, and identified two susceptibility loci of 3p21.31 and 9q34.2 to be significantly associated with severe COVID-19 at the genome-wide level. Based on a large-scale meta-analysis (N = 680,128), our group found that the *IFNAR2-IL10RB* gene cluster were significantly associated with COVID-19 susceptibility, and suggested that *IFNAR2* and *IL10RB* might have regulatory roles in the pulmonary immune response based on scRNA-seq data [25]. Consistently, Pairo-Gastineira et al. [24] conducted a GWAS study based on 2,244 critically ill COVID-19 patients and highlighted that several genes including *IFNAR2, DPP9*, and *OAS1* were significantly associated with severe COVID-19 at a genome-wide significance.

Two primary hypotheses were proposed for the involvement of immune genes in severe COVID-19 susceptibility. Whether the severe COVID-19-related risk genes associated with defective innate immune responses would induce persistent viral replication and resultant high viral loads, and whether an exaggerated genetically-mediated cytokine production contributes to the hyper-inflammation and poor outcome among severe COVID-19. However, the effects of these genetic determinants on the peripheral immune cells for severe COVID-19 remain largely unknown. In view of a purely genetic study or single cell sequencing study cannot address this critical question, we here leveraged comprehensive computational methods to combine a large-scale GWAS summary dataset with scRNA-seq data for identifying host genetics-influenced immune cell subpopulations involved in the etiology of severe COVID-19.

## Methods

### Single cell RNA-seq data on severe COVID-19

In this study, we downloaded four independent scRNA-seq datasets on COVID-19 in PBMC and BALF from the ArrayExpress database (Dataset #1, the accession number is E-MTAB-9357 from Su et al. study [10]), and the Gene Expression Omnibus (GEO) database (Dataset #2, the accession number is GSE149689 from Lee et al. study [18], Dataset #3, the accession number is GSE150861 from Guo et al. study [11], and Dataset #4, the accession number is GSE158055 [29]). The first dataset contained 270 peripheral blood samples including 254 samples with different COVID-19 severity (i.e., mild N = 109, moderate N = 102, and severe N = 50) and 16 healthy controls for scRNA-seq analysis. There were eight patients in dataset #2 with COVID-19 of varying clinical severity, including asymptomatic, mild, and severe, and four healthy controls with PBMCs. The dataset #3 included five peripheral blood samples collected from two severe COVID-19 patients at three different time points during tocilizumab treatment, containing two different stages: severe stage and remission stage. Within the dataset #4, 12 BALF samples were collected from lung tissues, including three moderate and nine severe patients. For all the datasets, the sample collection process were reviewed and approved by Institutional Review Board at the institutions where samples were originally collected. The COVID-19 severity was evaluated by using the World Health Organization (WHO) ordinal scale (WOS), the National Early Warning Score (NEWS), or the Diagnosis and Treatment of COVID-19 (Trail Version 6). Single-cell transcriptomes for these four datasets were gathered using the 10× Genomics scRNA-seq platform.

### Single cell RNA sequencing data processing

We performed normalization, clustering, and dimensionality reduction, differential expression gene (DEG) analysis, and visualization on these four independent scRNA-seq datasets with the Seurat R package [30]. The *SCTransform* function was used to scale and transform data, and linear regression model was applied to omit redundant variations caused by cellular complexity (i.e., cells expressed less than 200 genes or more than 2,500 genes were removed) or cellular quality (i.e., cells that had UMIs more than 10,000 and expressed reads of mitochondrial genes greater than 10% were removed). The *CellCycleSoring* function was applied to remove the effects of confounding factors. Principal component analysis (PCA) was carried out to extract principal components (PCs) that could explain most of datasets via using high variable genes. Top 20 PCs were utilized to conduct uniform manifold approximation and projection (UMAP) to embed the dataset into two dimensions. Subsequently, we constructed a shared nearest-neighbor graph (SNN) using the *FindNeighbors* function based on the top 20 PCs, and applied a graph-based modularity-optimization algorithm from the Louvain method [31] on this SNN for clustering the dataset with the cluster resolution set to 0.5. We used the *RunHarmony* function with PCA reduction method from harmony R package [32] to integrate samples to correct batch effects. The *FindConservedMarkers* function in Seurat was implemented to find differential expressed genes for determining cellular identity. Well-defined markers were used to annotate clusters, and uncharacterized clusters in the first round of clustering were extracted to run the second round of clustering (Supplemental Table S2). A total of 606,534 cells with 563,856 PBMC cells and 42,678 BALF cells were yielded from 300 samples based on the four independent scRNA-seq datasets (Supplemental Table S1 and Figure 1A). To allow comparison across samples and datasets, we used a common dictionary of gene symbols to annotate genes and these unrecognized symbols were removed.

**Figure 1.**
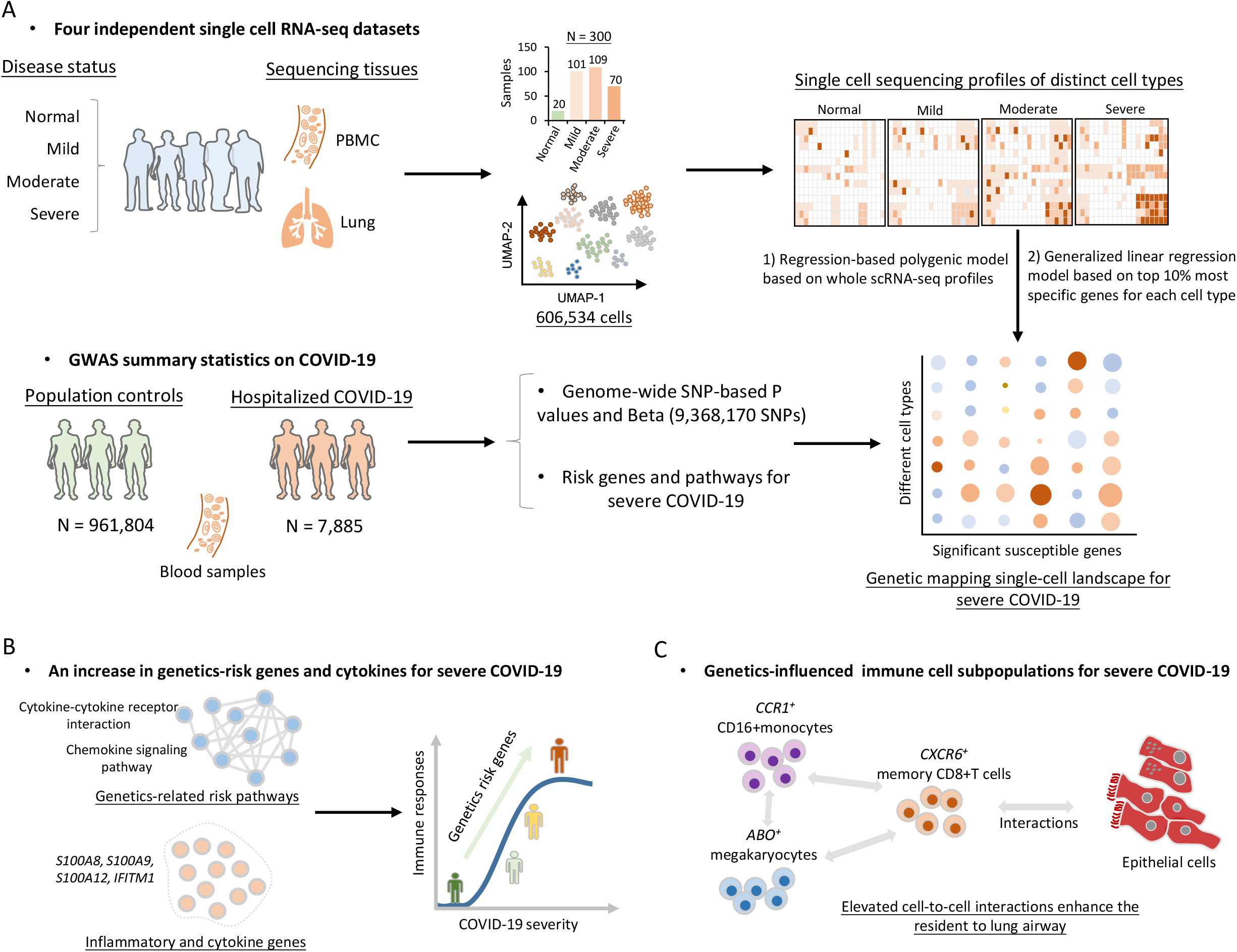
The workflow for this integrative genomic analysis. A) Combination of single cell RNA sequencing data and GWAS summary statistics on severe COVID-19 based on two independent methods. One method is regression-based polygenic model based on whole scRNA-seq profiles, and another is generalized linear regression model based on top 10% most specific genes for each cell type. B) An increase in genetics-risk genes and cytokines for severe COVID-19. C) Cellular interaction analysis of genetics-influenced immune cell subsets with epithelial cells.

### GWAS summary data on hospitalized COVID-19

The meta-GWAS summary data on severe COVID-19 round 4 (B2_ALL, Susceptibility [Hospitalized COVID-19 vs. Population]) were downloaded from the official website of the COVID-19 Host Genetic Consortium [22] (https://www.covid19hg.org/; analyzed file named: “COVID19_HGI_B2_ALL_leave_23andme_20201020.txt.gz”; released date of October 4 2020). There were 7,885 hospitalized COVID-19 patients and 961,804 control participants from 21 independent contributing studies. There was an overwhelming majority of participants in these contributing studies with European ancestry (93%). The meta-GWAS summary statistics contained P values, Wald statistic, inverse-variance meta-analyzed log Odds Ratio (OR) and related standard errors. The 1,000 Genomes Project European Phase 3 [33] was used as a panel for pruning. Results from 23&Me cohort GWAS summary statistics were excluded from our current analysis. Genetic variants without RefSNP number in the Human Genome reference builds 37 were filtered out, giving a total of 9,368,170 genetic variants satisfying the major allele frequency (MAF) over 0.0001 and the imputation score of greater tha 0.6. We used the *qqman* R package to figure both Manhattan plot and quantile-quantile (QQ) plot, and the web-based software of *LocusZoom* (http://locuszoom.sph.umich.edu/)[34] to visualize the regional association plots for significant risk loci.

### Hierarchical clustering analysis

To examine the similarity of the transcriptome profiles between cell types across different COVID-19 severities (Supplemental Figure S4), we merged the counts of UMI for each cell type according to normal, mild, moderate, and severe COVID-19. In order to normalize gene expression, we divided the counts of UMI for each gene by the counts of total UMI for all genes in each cell type and then multiplied by 100,000, as refer to the method in a previous study [18]. A median expression value of greater than 0.5 was used to calculate the relative change in each gene expression by dividing it by the median value for each gene, and the Pearson correlation coefficient (PPC) of the relative change in gene expression was used for current hierarchical clustering analysis.

### Gene-based association analysis

To perform a gene-based genetic association analysis of the meta-GWAS summary statistics on severe COVID-19, we leveraged the updated SNP-wise Mean model of MAGMA [35]. In this model, MAGMA computes a test statistic:

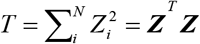

where N is the number of SNPs mapped in a gene and *Z*_*i*_ = **ϕ**(*p*_*i*_). Of note, **ϕ** is the cumulative normal distribution function and *p*_*i*_ is the marginal P value for a given SNP *i*. SNPs belonging to a specific gene were based on whether located in the gene body or within the +/- 20 kb upstream or downstream region of the gene. Furthermore, the model assumes ***Z*** ∼ MVN(***0, S***), where ***S*** is the LD matrix of the SNP genotypes. The LD matrix can be diagonalized and hence written as ***S*** = ***QAQ***^*T*^, where ***Q*** is an orthogonal matrix and ***A*** = diag(*λ*_1_, *λ*_2_,…, *λ*_*N*_) with *λ*_*j*_ being the *j*th eigenvalue of ***S***. The 1,000 Genomes Project Phase 3 European Panel [33] was used for calculating the LD information among SNPs extracted from GWAS summary data on COVID-19. ***D*** ∼ MVN(***0, I***_*K*_) is a random variable, where ***D*** = ***A***^***-0*.*5***^***Q***^***T***^ ***Z***. Then the sum of squared SNP Z-statistics as the following formula:

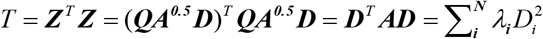

with *D*_*i*_ ∼ N(0,1) and 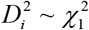. Namely, *T* follows a mixture distribution of independent 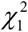 random variables. A total of 19,138 genes were included in the current analysis. We used the Benjamini-Hochberg false discovery rate (FDR) method, in which a gene with a FDR ≤ 0.05 (P ≤ 6.8×10^−5^) was interpreted as significant, to adjust for multiple testing.

### Pathway enrichment analysis

We applied the *built-in* functions of MAGMA [35], using the results from GWAS summary statistics as its input, to examine genome-wide enriched biological pathways for severe COVID-19. We calculated competitive P values by examining the results that the combined effect of genes within a pathway is significantly greater than the combined effect of all other genes, and 10,000 permutations was used to adjust competitive P values. Additionally, we leveraged the over-representation algorithm of the WebGestalt (http://www.webgestalt.org) [36] along with the significant genes as an input list to conduct a pathway enrichment analysis using the KEGG pathway resource [37]. The number of genes in each pathway was set to between 5 and 2,000, and the Benjamini-Hochberg FDR was used for multiple correction. To cluster these identified KEGG pathways, we performed a multidimensional scaling (MDS) analysis based on the Jaccard distance method [38], and constructed a pathway-pathway interaction network for these significantly enriched pathways setting the Jaccard distance > 0.1.

### Combining GWAS-based genetic signals with eQTL data

To uncover genetically-regulatory expression of genes associated with severe COVID-19, we conducted an integrative genomics analysis by using the S-PrediXcan [39] by combining meta-GWAS summary statistics with expression quantitative trait loci (eQTL) data for 49 tissues from the GTEx Project (version 8). S-PrediXcan mainly uses two linear regression models to analyze the association between predicted gene expression and severe COVID-19:

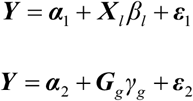

where ***α***_1_ and ***α***_2_ are intercepts, ***ε***_1_ and ***ε***_2_ are independent error terms, ***Y*** is the *n* dimensional vector for *n* individuals, ***X***_*l*_ is the allelic dosage for SNP *l* in n individuals, *β*_*l*_ is the effect size of SNP *l*, ***G***_*g*_ = Σ_*i*∈*gene*(*g*)_ *ω*_*ig*_ ***X***_*i*_ is the predicted expression calculated by *ω*_*lg*_ and ***X***_*l*_, in which *ω*_*lg*_ is derived from the GTEx Project, and *γ*_*g*_ is the effect size of ***G***_*g*_. The Z-score (Wald-statistic) of the association between predicted gene expression and severe COVID-19 can be transformed as:

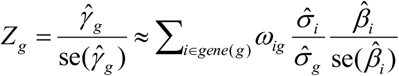

where 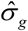 is the standard deviation of ***G***_*g*_ and can be calculated from the 1,000 Genomes Project European Phase 3 Panel, 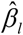 is the effect size from GWAS on COVID-19 and 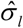 is the standard deviation of 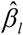. S-PrediXcan was run for each of 49 tissues with 659,158 gene-tissue pairs.

Furthermore, to increase the power to discover significant genes whose expression has similar regulations across multi-tissues, we utilized the S-MultiXcan [40] to meta-analyze these results from above S-PrediXcan analysis. S-MultiXcan fits a linear regression model of severe COVID-19 on predicted expression from multiple tissue models jointly:

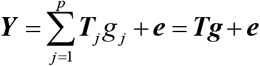

where 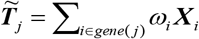 is the predicted expression of tissue *j*, and ***T***_*j*_ is the standardization of 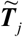 to *mean* = 0 and *standard deviation* = 1. *g*_*j*_ is the effect size for the predicted gene expression in tissue *j*, ***e*** is an error term with variance 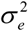, and *p* is the number of included tissues. There were 22,326 genes across 49 GTEx tissues with integrated convergent evidence in S-MultiXcan, and a gene with a value of FDR ≤ 0.05 (P ≤ 3.8×10^−5^) is considered to be significant.

### In silico permutation analysis

To explore the concordance of results from both MAGMA analysis (Gene set #1: N = 944, P ≤ 0.05) and S-MultiXcan analysis (Gene set #2: N =1,274, P ≤ 0.05), we performed an *in silico* permutation analysis which consisted 100,000 times (*N* _Total_) random selections [41, 42]. We first calculated the number of overlapped genes between Gene Set #1 and #2 (*N* _Observation_ = 302), then employed the total number of genes in S-MultiXcan analysis as background genes (*N* _Background_ = 22,326). By randomly selecting the same number of genes as Gene set #2 (N = 1,274) from the background genes, and after repeating it 100,000 times, we calculated the number of overlapped genes between Gene Set #1 and the sample we selected each time (*N* _Random_). Finally, we calculated the empirically permuted P value using the following formula: 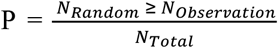, and empirical P value ≤ 0.05 is considered to be significant.

### Drug-gene interaction analysis

We conducted a drug-gene interaction analysis for identified genetics-risk genes by using protein-chemical interactions in the context of STRING-based PPI networks [43] and STITCH-based drug annotation information (v5.0, http://stitch.embl.de/) [44]. Only experimentally-validated gene-drug interactions with ranked confidence score were selected for constructing a drug-gene interaction network. To examine the potential therapeutic effects of highly-expressed genes in each immune cell, we conducted an enrichment analysis of 43 druggable categories based on the DGIdb database (https://www.dgidb.org/druggable_gene_categories). Additionally, we collected 1,263 human druggable proteins, which are therapeutic targets of clinical stage or approved drugs, from a previous study [26]. Among them, 704 proteins are targets for potential COVID-19-relevant drugs based on registers of clinical trials for COVID-19, approved immunomodulatory/anticoagulant drugs, or have biological functions associated with SARS-CoV-2 infection (Supplemental Table S11).

### Integrated analysis of GWAS summary statistics and scRNA-seq data

To identify genetically regulatory-related peripheral immune cells for severe COVID-19, we implemented the RolyPoly algorithm [45] to incorporate GWAS summary statistics with scRNA-seq data. Let *g*(*i*) stands for the gene associated with SNP *i, S*_*j*_ = {*i* : *g*(*i*) = *j*} be the SNP set with multiple SNPs associated with the gene *j*, and 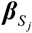 be a GWAS-based effect-size vector of *S*_*j*_ with a *priori* assumption that 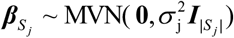. Following the *priori*, RolyPoly gives a polygenic linear model for 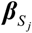:

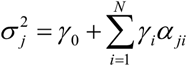

where *γ*_0_ is an intercept term, *α*_*ji*_(*i* = 1,2,…, *N*) are annotations such as cell-type-specific gene expression, and *γ*_*i*_ are annotation coefficients for *α*_*ji*_. To fit the observed and expected sum squared SNP effect sizes related to each gene by using the method-of-moments estimators, RolyPoly estimates *γ*_*i*_ by the following equation:

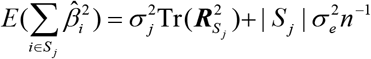

where 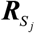 is the LD matrix of *S*_*j*_ and Tr represents the trace of a matrix. Finally, RolyPoly applies the block bootstrap method with 1,000 iterations to estimate standard errors 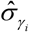 for calculating a *t*-statistic and corresponding P values. The PLINK (v1.90) [46] was used to calculate the LD between SNPs within the 1 Mb window based on the 1,000 Genome Project European Phase 3 panel [33]. We restricted the analysis to SNPs in the autosomes, and any SNPs with MAF ≤ 5% were excluded. The major histocompatibility complex region (Chr6: 25-35 Mbp) was also excluded due to the extensive LD in this region.

### Defining cell state scores

We leveraged cell state scores to assess the immunological degree of each immune cell type expressed a pre-curated expression gene set [11, 14, 29]. The cell state scores (CTS) were calculated based on the average expression of genes from the pre-curated gene set in the respective cell with the following formula: CTS*k*(*m*) = average(RE(GS*k, m*)) - average(RE(GS*n*, m)), where GS*k* is a pre-defined gene set *k* in a given cell *m*, and GS*n* is a control gene set that was randomly chosen on the basis of aggregate expression levels bins, which obtain a comparable distribution of expression levels and over size to that of the pre-curated gene set. RE represents the relative expression of GS*k* or GS*n*. The *AddModuleScore* function in Seurat [30] was applied to calculate the CTS with default parameters. We used the inflammatory and cytokine genes (N = 324 genes, Supplemental Table S10), cytokine-cytokine receptor interactions (N = 294 genes), chemokine signaling pathway (N = 189 genes), T cell activation (GO: 0042110), response to interferon alpha (GO: 0035455), response to interferon beta (GO: 0035456), leukocyte migration (GO: 0050900), 5 well-defined proliferating markers (*MK167, TYMS, NKG7, IL7R*, and *CCR7*), 6 well-defined exhaustion markers (*LAG3, TIGIT, PDCD1, CTLA4, HAVCR2*, and *TOX*), and 12 cytotoxicity-associated genes (*PRF1, IFNG, GNLY, NKG7, GZMB, GZMA, GZMH, KLRK1, KLRB1, KLRD1, CTSW*, and *CST7*) to define inflammatory cytokine, chemokine, T cell activation, IFN-α/β response, migration, proliferation, exhaustion, and cytotoxicity score, respectively.

### Cell-to-cell interaction analysis

To identify potential cellular interactions of *CCR1*^+^ CD16+monocytes and *CXCR6*^+^ memory CD8+T cells with other immune cells, we utilized the CellChat R package [47] for inferring the predicted cell-to-cell communications based on two normalized scRNA-seq datasets (dataset #1 of PBMC and dataset #4 of BALF). CellChat algorithm could examine the significance of ligand-receptor interactions between two cell types depending on the expression of important factors, including stimulatory and inhibitory membrane-bound co-receptors, soluble agonists and antagonists. The communication probability of a signaling pathway was derived from the sum of probabilities of their ligand-receptor interactions. We only concentrated on the ligand-receptor interactions that significantly associated with severe COVID-19 compared with normal control.

### Statistical analysis

The Wilcoxon sum-rank test was used to assess DEGs in mild, moderate, and severe COVID-19 group compared with normal control. The Mann-Kendall trend analysis was applied to evaluate the significance of cell state cells with elevated severities of COVID-19. Pathway- and disease-based enrichment analyses used the hypergeometric test to identify remarkable biological pathways and disease-terms. The Pearson correlation analysis was used to calculate the correlation coefficient of highly-expressed genes in *CCR1*^+^ CD16+monocytes between moderate and severe patients. The paired Student’s t test was used to calculate the significance of ligand/receptor interactions of *CCR1*^+^ CD16+monocytes and *CXCR6*^+^ memory CD8+T cells with other immune cells between normal control and severe COVID-19.

## Results

### The computational framework for integrating single-cell transcriptomes and GWAS on COVID-19

As shown in Figure 1, we devised a computational framework to parse the host genetics-modulated immune cell subpopulations implicated in severe COVID-19. It included three main parts: 1) integrative analysis that combined GWAS summary statistics with scRNA-seq data to genetically map single-cell landscape for severe COVID-19 (Figure 1A); 2) identifying genetics-risk genes, pathways, and immune cell subpopulations that contributed to cytokine storms among severe patients (Figure 1B); and 3) uncovering the cellular interactions of genetics-modulated immune cell subsets, as well as their functions with cells in lung tissues (Figure 1C).

### Identification of immune cell types associated with severe COVID-19

To parse the host genetics-influenced immune responses at single cellular level in PBMCs for severe COVID-19, we subjected three independent scRNA-seq datasets with 563,856 cells to UMAP based on highly variable genes using the Seurat (See Methods) [30]. There was identification of 13 distinct clusters unbiased by patients with different severities (Supplemental Figure S1). We leveraged well-known marker genes to assign these clusters to 13 distinct cell types, including mature B cells, megakaryocytes, naïve B cells, CD34+progenitors, dendritic cells, natural killer (NK) cells, CD14+monocytes, CD16+monocytes, memory CD4+T cells, naïve CD4+T cells, naïve C D8+T cells, memory CD8+T cells, and effector CD8+T cells (Supplemental Figures S2-S3).

While performing the hierarchical clustering analysis on the scRNA-seq profiles, we discovered that cell types were the primary determinants of their clustering, followed by disease severities, indicating both COVID-19 pathology and immune cell types might have crucial roles in altered patterns of immune transcriptome instead of technical artifacts (Supplemental Figure S4). As a vital feature for reflecting the alterations of immune responses, we examined the relative proportions of peripheral immune cells across different COVID-19 groups in comparison with normal group. The proportions of CD14+monocytes, megakaryocytes, and CD34+progenitors were significantly elevated in moderate and severe patients, whereas the proportions of CD16+monocytes, effector CD8+ T cells, memory CD8+T cells, memory CD4+T cells, naïve CD4+T cells, and NK cells were significantly decreased with the increased severities (Supplemental Figure S5).

### Identification of genetic risk loci associated with severe COVID-19

Through performing a meta-analysis of 21 independent GWAS studies from the COVID-19 Host Genetic Consortium, eight genomic loci were identified to be associated with hospitalized COVID-19 at a genome-wide significant level, including 1p22.2 (rs2166172, P = 2.74×10^−8^), 3p21.31 (rs35081325, P = 3.32×10^−58^, and rs33998492, P = 3.59×10^−14^), 6p21.33 (rs143334143, P = 1.28×10^−10^), 7p11.2 (rs622568, P = 2.57×10^−8^), 9q34.2 (rs505922, P = 2.24×10^−9^), 12q24.13 (rs2269899, P = 3.24×10^−8^), 19p13.3 (rs2109069, P = 6.4×10^−13^), and 21q22.11 (rs13050728, P =1.91×10^−11^) (Figure 2A, Supplemental Table S3, Figure S6, and Materials S2). Among these eight loci, three loci, 1p22.2, 6p21.33 and 7p11.2, were newly identified. It should be noted that there were two independent genetic association signals (Index SNPs: rs35081325 and rs33998492) in the 3p21.31 locus for severe COVID-19 (Figure 2B and Supplemental Figure S7A-C). Using the Variant2Gene (V2G) algorithm [48], we prioritized *CXCR6* as a candidate causal gene for rs35081325 and causal gene *CCR1* for rs33998492 (Supplemental Method S1).

**Figure 2.**
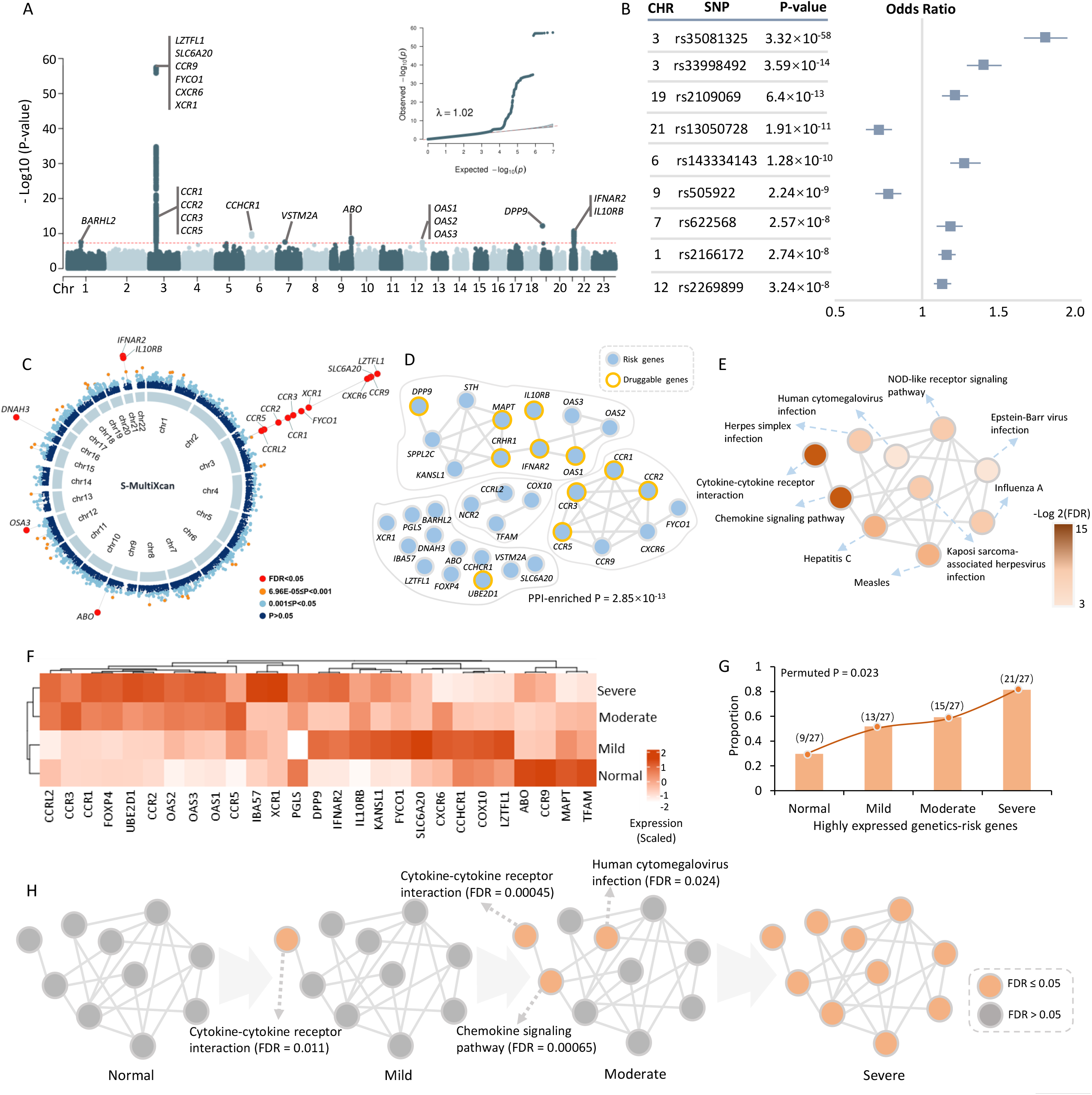
Risk genes and pathways associated with hospitalized COVID-19 from meta-GWAS summary data. A) Manhattan plot and quantile-quantile (QQ) plot of meta-GWAS analysis highlighting eight risk genetic loci for hospitalized COVID-19. The red horizontal line represents the genome-wide significance threshold of P < 5×10^−8^. The genomic inflation factor λ = 1.02. B) Nine index SNPs within eight genomic loci associated with hospitalized COVID-19. Left panel shows the P value of each index SNP, and right panel shows the odds ratio with 95% confidence interval. C) Circus plot showing the results of S-MultiXcan-based analysis. The inner ring demonstrates the 22 autosomal chromosomes (Chr1-22). In the outer ring, a circular symbol represents a specific gene and color marks the statistical significance of the gene for hospitalized COVID-19 (Red marks FDR < 0.05, orange indicates 6.96×10^−5^ ≤ P < 0.001, light blue marks 0.001 ≤ P ≤ 0.05, and dark blue indicates P > 0.0). D) PPI network of these 34 identified risk genes based on the STRING database (v11.0, https://string-db.org/). Orange ring represents druggable genes targeted by at least one known drug. E) Network module constructed by using the Jaccard distance showing the connectivity of 10 significant pathways enriched by 34 risk genes. F) Heatmap showing the results of hierarchical clustering analysis of 27 risk genes on COVID-19 severity. Seven risk genes did not expressed in the dataset #1, and the expression level of each gene was scaled. G) The proportion of highly-expressed genes among 27 risk genes in normal controls and in the three phases of COVID-19 (mild, moderate, and severe patients). Using 10,000 times of permutation analysis to calculate the significance of the observation (permuted P = 0.023). H) Plot showing an increase of the significantly enriched pathways in the network module with elevated COVID-19 severities. Orange color represents a significant enriched pathway (FDR ≤ 0.05) and gray color represents a non-significant enriched pathway (FDR > 0.05).

Furthermore, the index SNP of rs505922 (P = 2.24×10^−9^) in the 9q34.2 locus is highly LD with the reported SNP of rs657152 (R^2^ = 0.874) [27] and rs8176719 (R^2^ = 0.876) [25]. Based on the top-ranked V2G score for rs505922, we prioritized *ABO* as a potential causal gene contributing susceptibility to severe COVID-19. By performing a MAGMA gene-level association analysis, we observed that 25 genes including *CXCR6, CCR1, IFNAR2, IL10RB*, and *OAS1* were significantly associated with severe COVID-19 (FDR < 0.05, Supplemental Figure S8 and Table S4). GWAS-based pathway enrichment analysis revealed that 19 biological pathways, including cytokine-cytokine receptor interaction, influenza A, and TNF signaling, were significantly associated with hospitalized COVID-19 (Supplemental Figure S9 and Table S5).

### Integrative analysis of GWAS on severe COVID-19 with GTEx eQTL data

To obtain combined signals from multiple tissues [49], we leveraged S-MultiXcan to meta-analyze the tissue-specific associations from 49 tissues in GTEx (see Methods), which showed that the genetically predicted expressions of 16 genes were significantly associated with severe COVID-19 (FDR < 0.05, Figure 2C and Supplemental Table S6). Of note, 14 of 16 genes (87.5%) were identified to be significant in MAGMA analysis (Supplemental Figure S10A-B). Through conducting S-PrediXcan analysis of blood and lung tissues that were linked with SARS-CoV-2 infection, we found eight genes whose genetically-regulated expression were significantly associated with severe COVID-19 (FDR < 0.05, Supplemental Table S7). Using *in silico* permutation analysis, we further observed that there existed a high consistence among results from MAGMA, S-PrediXcan, and S-MultiXcan analyses (P < 1.0×10^−5^, Supplemental Figure S11A-C). The aforementioned multiple genomic analyses identified 34 risk genes that showed supportive evidence of involvement in the etiology of COVID-19 (Supplemental Figure S12A-B).

### Functional characterization of 34 risk genes for severe COVID-19

The result of a network-based enrichment analysis suggested that 22 of 34 risk genes were significantly enriched in a PPI subnetwork (P = 2.85×10^−13^, Figure 2D), which is consistent with the consensus that disease-related genes are more densely connected [50, 51]. To functionally characterize the drug targets of these genes, we conducted a drug-gene interaction analysis and identified 11 genes including *CCR1, IFNAR2, IL10RB*, and *OAS1* were targeted by at least one known drug (Figure 2D and Supplemental Figure S14), of which some genes including *CCR1, IFNAR2*, and *IL10RB* have been reported to be drug targets for treating severe COVID-19 patients [25, 26]. Furthermore, these 34 genes were significantly enriched in a functional module consisting of 10 biological pathways (Figure 2E, Supplemental Table S8 and Figure S13), among which two top-ranked ones being cytokine-cytokine receptor interaction and chemokine signaling pathway (FDR < 0.05). Most of these enriched pathways have been reported to be implicated in COVID-19 [25, 52, 53].

Based on the expression profile of dataset #1, we conducted a hierarchical clustering analysis of these identified risk genes on COVID-19 severity, and found that these risk genes predisposed be highly-expressed in severe patients compared to normal group (Permuted P = 0.023, Figure 2F-G). Consistently, the number of significant enriched pathways were elevated with increased severities (Figure 2H). Genes in both cytokine-cytokine receptor interaction and chemokine signaling pathways showed significantly high expressions in the early phase of SARS-CoV-2 infection (Figure 2H), suggesting that these two pathways could play critical roles in the initiation of COVID-19.

### Genetics-influenced peripheral immune cell types for severe COVID-19

To identify genome-wide genetics-influenced immune cells for severe COVID-19, we first leveraged a regression-based polygenic model to integrate GWAS summary data on severe COVID-19 with single-cell transcriptomic profiles (dataset #1) according to different COVID-19 severities (See methods). We found that CD16+monocytes were significantly associated with three phases of COVID-19, mature B cells showed remarkable associations with mild COVID-19, megakaryocytes were significantly associated with moderate and severe COVID-19, and memory CD8+T cells showed significant associations with severe COVID-19 (simulated P < 0.05, Figure 3A). Further, we used a generalized linear regression model to validate these severe COVID-19-associated cell types by conditioning on the 10% most specific genes for each type, and consistently found that CD16+monocytes and megakaryocytes showed notable associations with severe COVID-19 (P < 0.05, Supplemental Method S2). These results indicated that CD16+monocytes, megakaryocytes, and memory CD8+T cells were more vulnerable to the influence of genetic components on severe-stage patients.

**Figure 3.**
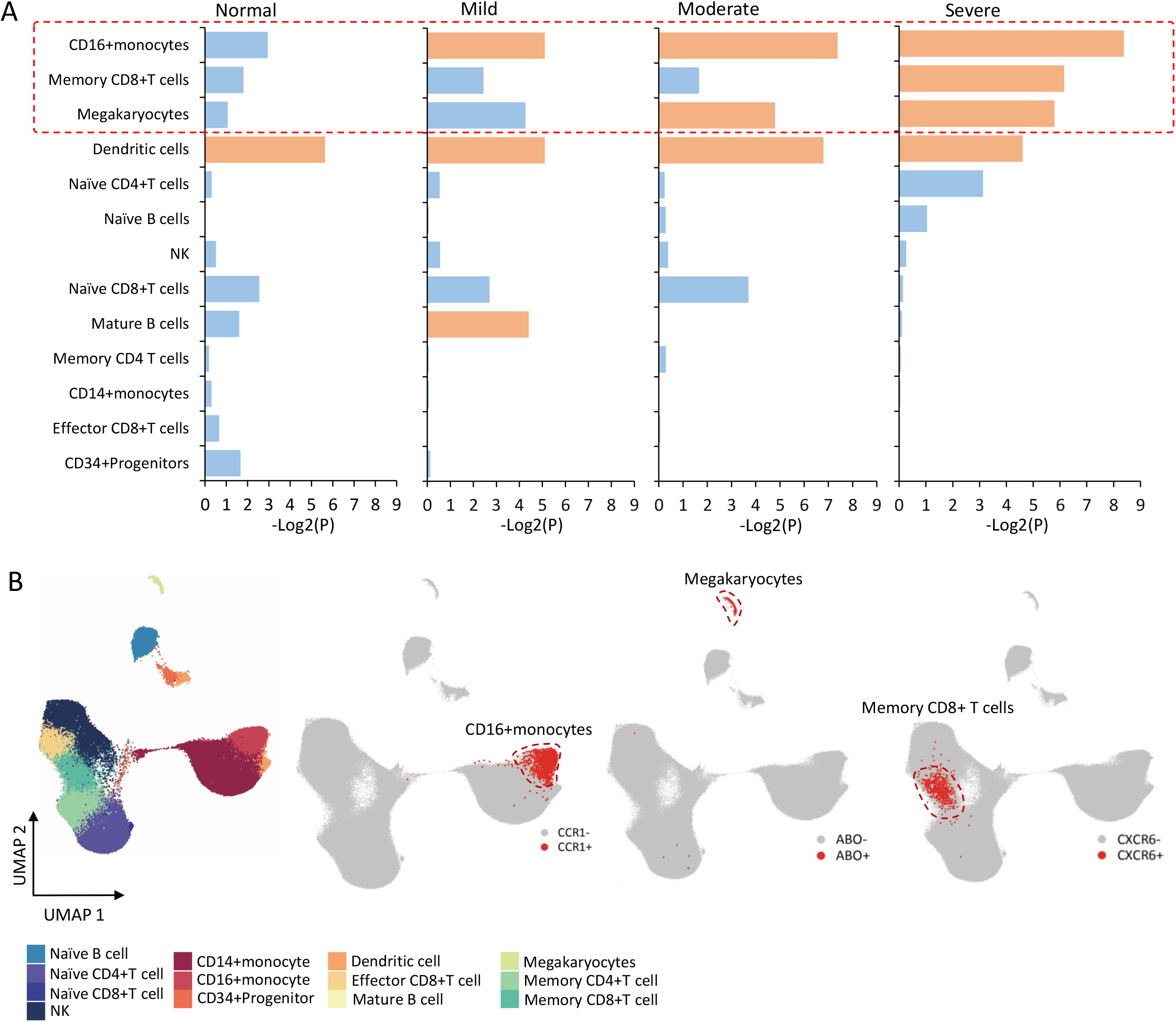
Integrative analysis identifies genetic associations between peripheral immune cells and severe COVID-19. A) Bar graph showing the results of the combination of scRNA-seq data and GWAS summary statistics on severe COVID-19 based on the RolyPoly among normal controls and patients with different severities (i.e., mild, moderate, and severe). The y-axis shows the 13 cell types, and x-axis shows mean negative log-transformation P value (-Log2(P)). Orange color indicates a cell type showing a significant association, and light blue represents there is no significant association. B) UMAP projections of peripheral immune cells colored by annotated cell types. The plot showing the region of CD16+monocytes, megakaryocytes, and memory CD8+T cells. Red dot represents positive gene expressions of *CCR1*^+^, *ABO*^+^, and *CXCR6*^+^, and gray stands for negative cells.

Based on the specificity algorithm used in MAGMA, we noticed that the top specific cell type of *CCR1* was CD16+monocytes, *CXCR6* was most specifically expressed in memory CD8+T cells, and *ABO* was specific to megakaryocytes (Supplemental Figure S15A), recalling that *CXCR6, CCR1* and *ABO* were prioritized to be candidate causal genes for severe COVID-19 based on the V2G score in above genetics-based analysis. Compared with other cell types, *CCR1* was primarily expressed in CD16+monocytes (24.77%), *CXCR6* was mainly expressed in memory CD8+T cells (40.29%), and the *ABO*-expressed cells were highly specific to megakaryocytes (54.63%) (Supplemental Figure S15B and Table S9). To gather additional empirical support, we analyzed the combined dataset of both datasets #2 and #3 as a validation and found *CCR1, CXCR6*, and *ABO* showed a consistent specificity in the three cell types (Supplemental Figure S16).

Given that the primary goal of current study was to characterize genetics-influenced peripheral immune cell types for severe COVID-19, the majority of our subsequent detailed analyses would be concentrated on three immune cell subpopulations: *CCR1*^+^ CD16+monocytes, *ABO*^+^ megakaryocytes, and *CXCR6*^+^ memory CD8+T cells (Figure 3B).

### CCR1^+^ CD16+monocytes and ABO^+^ megakaryocytes contributing higher risk to cytokine storm

The accumulating lines of evidence [29, 54] have suggested that subsets of monocytes and megakaryocytes might be the major resources of inflammatory storm. We sought to examine whether *CCR1*^***+***^ CD16+monocytes and *ABO*^***+***^ megakaryocytes play more important roles in cytokine storm among severe patients. As for *CCR1*^***+***^ CD16+monocytes, we found that the inflammatory cytokine score was significantly higher than that of *CCR1*^***-***^ CD16+monocytes (P = 2.5×10^−7^, Figure 4A). Consistently, the combined score of both cytokine-cytokine receptor interaction and chemokine signaling pathway was prominently higher in *CCR1*^***+***^ CD16+monocytes (P < 2.2×10^−16^, Supplemental Figure 17A). Compared with *CCR1*^***-***^ CD16+monocytes, there were 351 significantly highly-expressed genes in *CCR1*^***+***^ CD16+monocytes, such as inflammatory and cytokine genes of *IL1B, IL27, CXCL10, CXCL8, CD14*, and *OSM* (FDR < 0.05, Figure 4B and Supplemental Table S11), which have been documented to be associated with the inflammatory response and chemotaxis of immune cells among COVID-19 patients [10, 15, 55, 56]. Functionally, 19 KEGG pathways were significantly overrepresented by the 351 highly-expressed genes (FDR < 0.05, Figure 4C and Supplemental Table S12), including cytokine-cytokine receptor interaction and chemokine signaling pathway, reminiscing that most of them were identified in above genetics-based pathway analysis. Additionally, these highly-expressed genes among *CCR1*^***+***^ CD16+monocytes have a remarkably higher proportion of druggable genes and COVID-19-associated druggable genes (P ≤ 0.01, Supplemental Figure S17 and Table S13).

**Figure 4.**
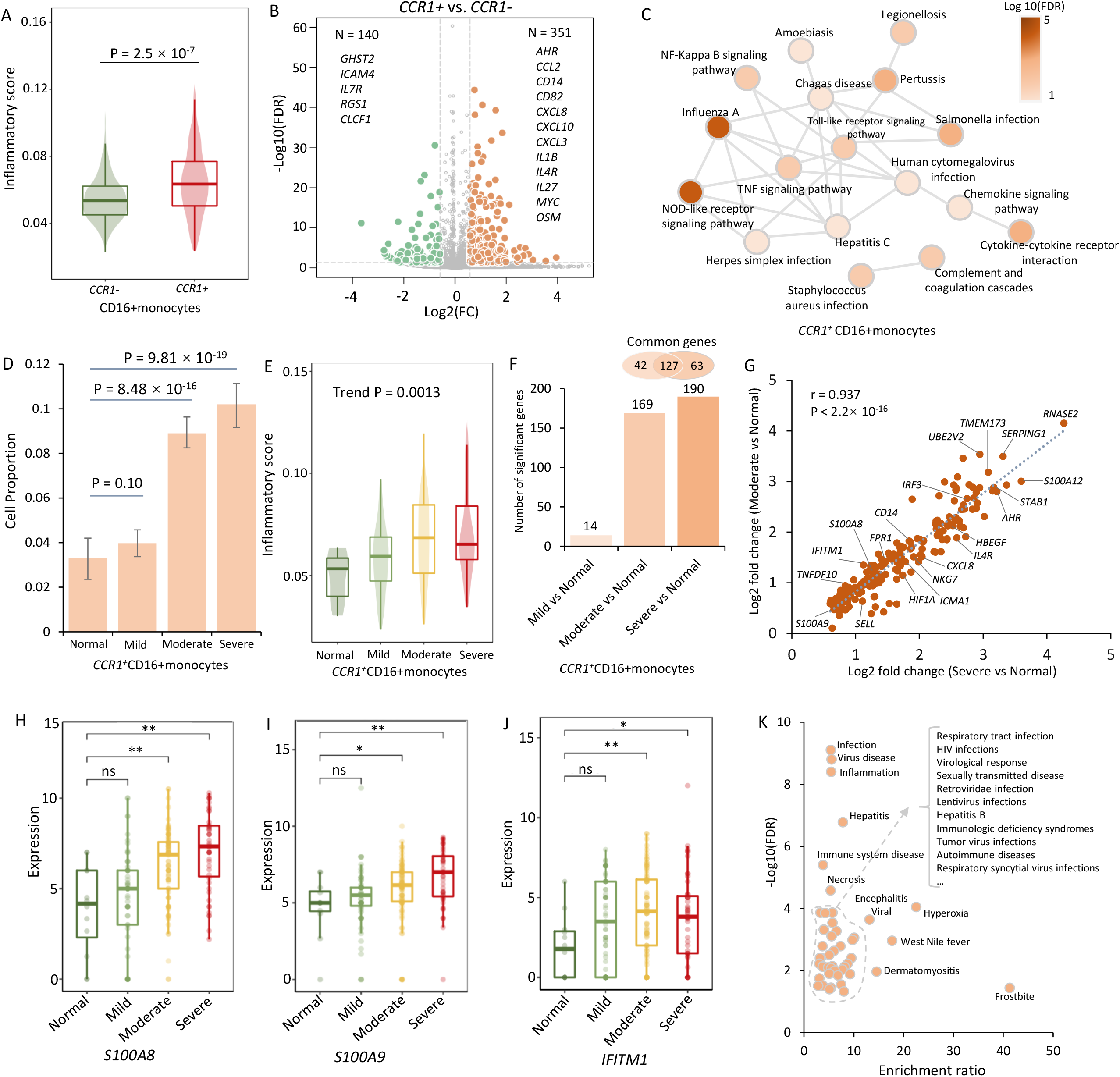
*CCR1*^+^ CD16+momocytes contributes higher risk to cytokine storms among severe COVID-19 patients. A) Boxplot showing the difference in inflammatory cytokine score between *CCR1*^+^ and *CCR1*^-^ CD16+ monocytes. Two-side Wilcoxon sum-rank test was used. B) Volcano plot showing differentially expressed genes between *CCR1*^+^ and *CCR1*^-^ CD16+ monocytes. C) Significantly enriched pathways by 351 highly-expressed genes among *CCR1*^+^ CD16+ monocytes. Color legend represents the log transformed FDR value (-Log10(FDR)). D) Bar graph showing the proportion of *CCR1*^+^ CD16+ monocytes among normal, mild, moderate, and severe groups. E) Boxplot showing the inflammatory cytokine score of *CCR1*^+^ CD16+ monocytes among normal, mild, moderate, and severe groups. The Mann-Kendall trend analysis was used. F) Bar graph showing the differentially up-DEGs among different COVID-19 patients compared with normal controls. Namely, mild COVID-19 vs. normal, moderate COVID-19 vs. normal, and severe COVID-19 vs. normal. Venn plot on top of bar showing the overlapped up-DEGs between moderate and severe patients. G) The correlation of up-DEGs between moderate and severe patients. Pearson correlation analysis was used to calculate the correlation coefficient and P value. H)-J) Representative up-DEGs among *CCR1*^+^ CD16+ monocytes showing significantly elevated expressions with increased COVID-19 severities. H) *S100A8*, I) *S100A9*, and J) *IFITM1*. K) Disease-terms enrichment analysis on 190 up-DEGs based on the GLAD4U database. The y-axis shows -Log10(FDR), and x-axis shows the enrichment ratio.

The cell percentage of *CCR1*^***+***^ CD16+monocytes showed a notable elevation among moderate and severe patients compared with normal controls (P < 0.001), with no significant difference between mild patients and normal controls (P = 0.1, Figure 4D). Furthermore, the inflammatory cytokine scores among *CCR1*^***+***^ CD16+monocytes were significantly elevated with increased severities (Trend P = 0.0013, Figure 4E). In comparison with normal controls, mild, moderate, and severe patients displayed significantly up-regulated expressions (up-DEGs) with 14, 169, and 190 genes respectively (FDR < 0.05, Figure 4F and Supplemental Figure S17D). Notably, there existed a high correlation between up-DEGs of moderate and severe patients (r = 0.937, P < 2.2×10^−16^; Figure 4G), such as *S100A8, S100A9*, and *IFITM1* (Figure 4H-4J), indicating a similar expression pattern between moderate and severe patients. Accumulating release of massive amounts of calprotectin (*S100A8/S100A9*) in monocytes contributes to inflammatory response among severe COVID-19 patients [10, 16, 29].

Furthermore, these 190 up-DEGs were significantly enriched in disease-terms associated with viral infection and inflammation and 17 functional GO-terms (FDR < 0.05, Figure 4K, Supplemental Figure S17E and Tables S14-S15), including interferon alpha/beta signaling and interferon gamma signaling. These interferon-related genes including *IRF3, IRF2, IFI6, IFITM1, ISG15*, and *ICAM1* may induce autoinflammatory and autoimmune conditions contributing to the innate immune cells against SARS-CoV-2 infection [57, 58]. Of note, a high proportion of 63.68% among 190 up-DEGs such as *CXCL8, IFITM1, S100A8*, and *S100A9* were annotated into 15 potential druggable gene categories (Supplemental Figure S17F-L and Table S16). These results indicated that interferon-related genes among *CCR1*^***+***^ CD16+monocytes have instrumental effects in exacerbating inflammation among severe patients.

In addition, we found that *ABO*^***+***^ megakaryocytes had a significantly higher inflammatory cytokine score than that in *ABO*^***-***^ cells (P < 0.001, Supplemental Figure S18A-B). Compared with *ABO*^***-***^ megakaryocytes, 424 genes were significantly highly-expressed in *ABO*^*+*^ megakaryocytes (FDR < 0.05, Supplemental Figure S18C and Table S17). These 424 highly-expressed genes were significantly enriched in systemic lupus erythematosus, alcoholism, and platelet activation (FDR < 0.05, Supplemental Figure S18D and Table S18). Similar to *CCR1*^***+***^ CD16+monocytes, the cell percentage of *ABO*^***+***^ megakaryocytes was significantly elevated among moderate and severe patients (P < 0.01, Supplemental Figure S18E). Among *ABO*^*+*^ megakaryocytes, 20 and 35 up-DEGs were notably associated with moderate and severe patients, respectively (FDR < 0.05, Supplemental Figure S18F-G). There was a highly overlapped rate of these up-DEGs between moderate and severe COVID-19 groups, including *ACP1, S100A8*, and *A100A9* (18/20 = 90%, Supplemental Figure S18F-N). These 35 up-DEGs were annotated to 12 druggable gene categories and significantly enriched in several disease terms (Supplemental Figure S18H and Tables S19-S20), such as shock and thrombocytopenia, which were reported to be associated with COVID-19 [59]. Overall, these results suggest that both *CCR1*^***+***^ CD16+monocytes and *ABO*^***+***^ megakaryocytes contribute higher risk to inflammatory storm among severe patients.

### CXCR6^+^ memory CD8+T cells convey risk to severe COVID-19

Earlier studies [10, 60] have indicated that polyfunctional T cells play important roles in dominating the anti-viral infection immune response and can release a substantially higher amount of multiple distinct cytokines and chemokines in comparison to other T cells. It is plausible to infer that there exist subsets of memory CD8+T cells predisposing to be multi-functional for against SARS-CoV-2 infection. We calculated several immunological features to evaluate whether *CXCR6*^***+***^ memory CD8+T cells have a higher polyfunctionality than *CXCR6*^***-***^ memory CD8+T cells. Compared with *CXCR6*^***-***^ memory CD8+T cells, we found that scores of cytokine, chemokine, IFN-ɑ/β response, T cell activation, proliferation, and migration were significantly higher among *CXCR6*^***+***^ memory CD8+T cells (P < 0.05, Figure 5A-D and Supplemental Figure S19A-C). There were 158 highly-expressed genes among *CXCR6*^***+***^ memory CD8+T cells in comparison with *CXCR6*^***-***^ cells (FDR < 0.05, Figure 5E). These highly-expressed genes were significantly enriched in two biological pathways of cytokine-cytokine receptor interaction and inflammatory bowel (FDR < 0.05, Supplemental Figure S19D and Table S21). The chemokine signaling pathway showed a suggestive enrichment (P < 0.05). These highly-expressed genes contained numerous pro-inflammatory cytokine and chemokine genes, such as *CCR1, CCR2, CCR5, CCR6, CCL3L1, IFNGR1, IL18R1, IL23R, MYC*, and *TNFSF14*, which may be associated with the activation of memory CD8+T cells.

**Figure 5.**
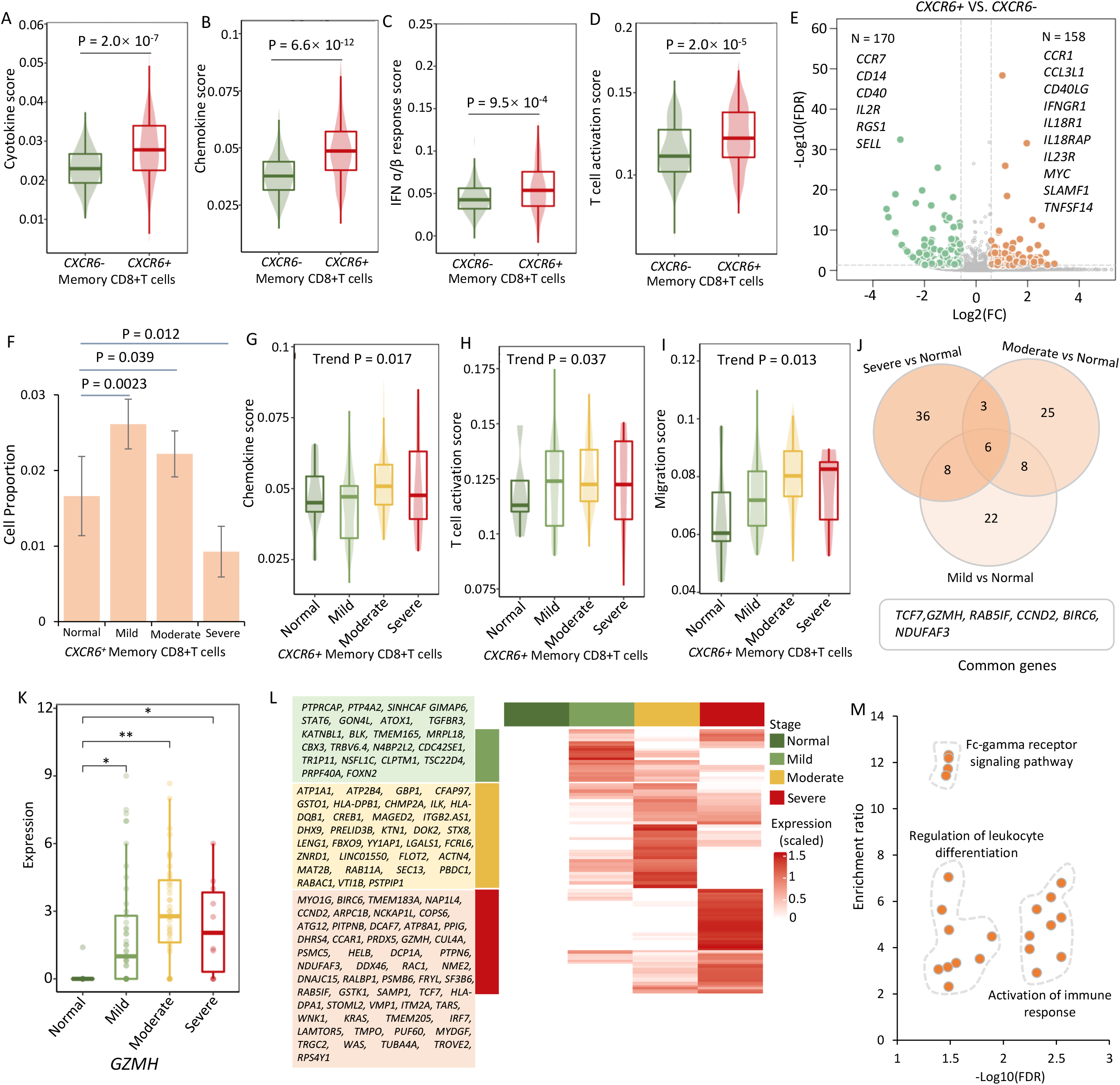
Multi-functionality of *CXCR6*^+^ memory CD8+T cells for severe COVID-19. A)-D) Boxplots showing the difference in (A) cytokine score, (B) chemokine score, (C) IFN-ɑ/β response score, and (D) T cell activation score between *CXCR6*^+^ and *CXCR6*^-^ memory CD8+T cells. Two-side Wilcoxon sum-rank test was used. E) Volcano plot showing differentially expressed genes between *CXCR6*^+^ and *CXCR6*^-^ memory CD8+T cells. F) Bar graph showing the proportion of *CXCR6*^+^ memory CD8+T cells among normal, mild, moderate, and severe groups. G)-I) Boxplots showing the (G) chemokine score, (H) T cell activation score, and (I) migration score of *CXCR6*^+^ memory CD8+T cells among normal, mild, moderate, and severe groups. The Mann-Kendall trend analysis was used. J) Venn plot showing the overlapped up-DEGs between pairwise comparisons: mild vs. normal, moderate vs. normal, and severe vs. normal. K) Representative gene of *GZMH* among *CXCR6*^+^ memory CD8+T cells showing significantly elevated expressions with increased COVID-19 severities. L) Heatmap showing up-DEGs in *CXCR6*^+^ memory CD8+T cells from pairwise comparisons: mild vs. normal, moderate vs. normal, severe vs. normal. The up-DEGs listed in the green panel were from mild vs. normal, yellow panel were from moderate vs. normal, and orange panel were from severe vs. normal. M) Scatter plot showing the enriched GO biological processes by 108 up-DEGs among *CXCR6*^+^ memory CD8+T cells. The x-axis shows -Log10(FDR), and y-axis shows the enrichment ratio.

Furthermore, the cell proportion of *CXCR6*^***+***^ memory CD8+T cells was significantly higher among both mild and moderate COVID-19 than that among normal group (P < 0.05), whereas the cell proportion of *CXCR6*^***+***^ memory CD8+T cells among severe COVID-19 was remarkably lower than that among normal group (P = 0.012, Figure 5F). Consistently, we found that the scores of chemokine, T cell activation, and migration were increased with the increasing patient severities among *CXCR6*^***+***^ memory CD8+T cells (Trend P < 0.05, Figure 5G-I), and that lower cytotoxicity score and exhaustion score were observed among moderate-to-severe patients (Trend P < 0.05, Supplemental Figure S19E-F). Additionally, we found 44, 42, and 53 up-DEGs that were notably associated with mild, moderate, and severe COVID-19, and there were six significant common genes across three phases of COVID-19, including *TCF7, GZMH, RAB5IF, CCND2, BIRC6*, and *NDUFAF3* (Figure 5J-K and Supplemental Figure S19G-N). The gene of *TCF7* was an essential factor in memory CD8+T cell differentiation [61], and *GZMH* was reported to mediate antiviral activity through direct cleavage of viral substrates [62]. These 108 up-DEGs were found to be significantly enriched in 22 functional GO-terms, including Fc-gamma receptor signaling pathway, regulation of leukocyte differentiation, and activation of immune response (Figure 5L-M and Supplemental Table S22). Overall, these results indicated that *CXCR6*^***+***^ memory CD8+T cells have an enhanced propensity to be multi-functional and activated T cells involved in severe COVID-19.

### Elevated cellular interactions may enhance the resident to lung airway for COVID-19

To gain refined insights into *CCR1*^+^ CD16+monocytes and *CXCR6*^+^ memory CD8+T cells, we examined the cellular interactions among cell populations in PBMCs and BALFs according to the COVID-19 disease status using the CellChat algorithm [47]. For *CCR1*^+^ CD16+monocytes in PBMCs, we found a notable increase in cell-to-cell interactions with other immune cells among severe patients than that in normal controls (P < 0.05, Figure 6A and Supplemental Figure S20). There was no statistical difference in cellular communications of *CCR1*^-^ CD16+monocytes with other cells between normal and COVID-19 patients (P > 0.05, Figure 6B). Compared with normal controls, *CCR1*^+^ CD16+monocytes showed elevated interactions with megakaryocytes, memory CD8+T cells, NK, effector CD8+T cells, and CD14+monocytes among severe patients (Supplemental Figure S20). There were 14 ligand-receptor interactions observed to be remarkably dominated among severe patients (Figure 6C), including *ANXA1-FPR1, ITGB2-ICAM2/CD226, LGALS9-CD44, SELPLG-SELL/SELP, APP-CD74*, and *THBS1-CD36/CD47*.

**Figure 6.**
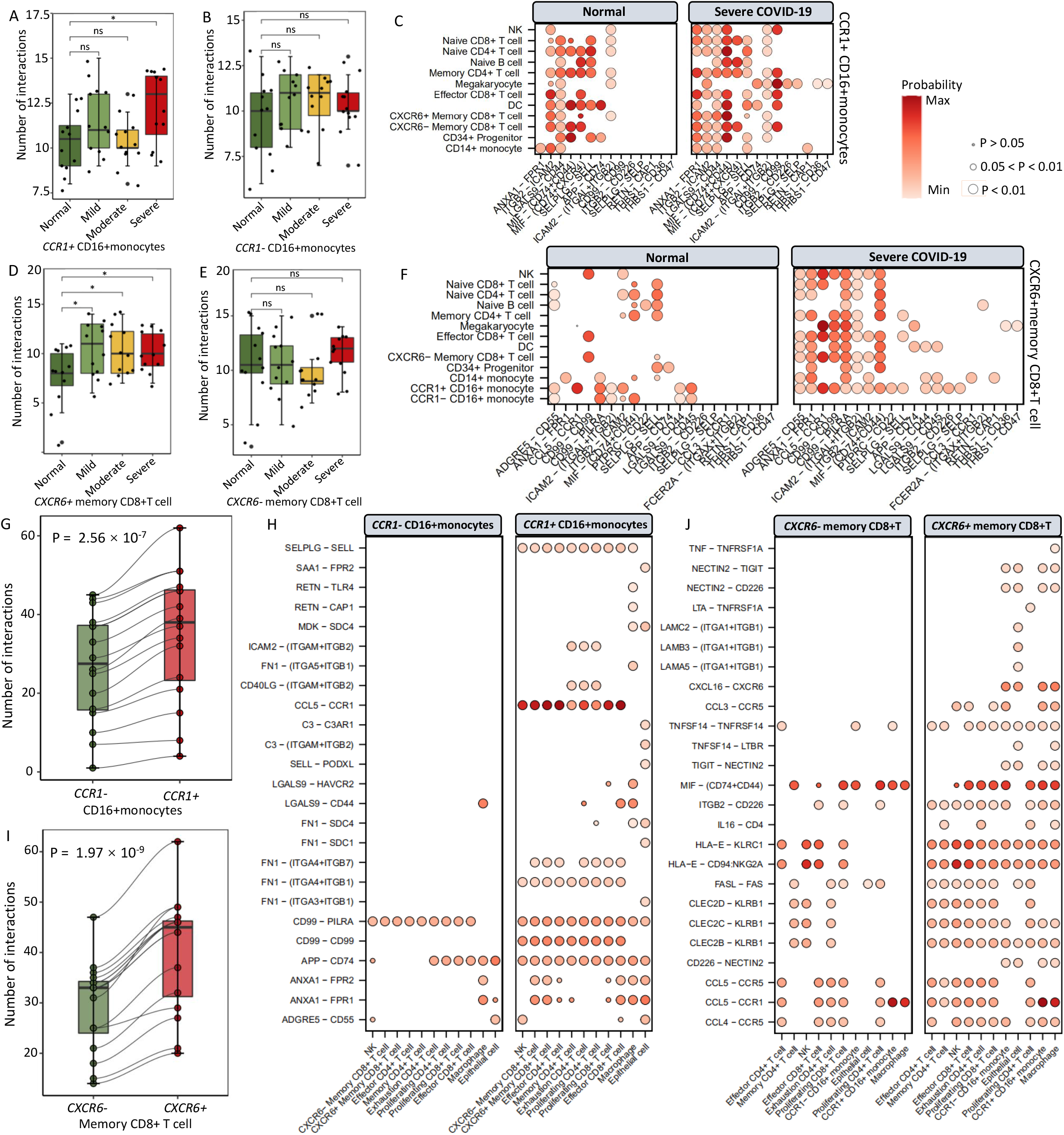
Cell-to-cell interactions of *CCR1*^+^ CD16+momocytes and *CXCR6*^+^ memory CD8+T cells with other cells in PBMC and BALF. A)-B) Boxplot showing the number of cellular interactions of (A) *CCR1*^+^ CD16+ monocytes and (B) *CCR1*^-^ CD16+ monocytes with other immune cells in PBMC between normal controls and patients with increased COVID-19 severities. C) Predicted cellular interactions of *CCR1*^+^ CD16+ monocytes with other immune cells in PBMC, comparing severe COVID-19 vs. normal control. D)-E) Boxplot showing the number of cellular interactions of (D) *CXCR6*^+^ memory CD8+T cells and (E) *CXCR6*^-^ memory CD8+T cells with other immune cells in PBMC between normal controls and patients with increased COVID-19 severities. F) Predicted cellular interactions of *CXCR6*^+^ memory CD8+T cells with other immune cells in PBMC, comparing severe COVID-19 vs. normal control. G) Boxplot showing an increase in cellular interactions with other cells in BALF for *CCR1*^+^ CD16+ monocytes than *CCR1*^-^ CD16+ monocytes. H) Predicted cellular interactions with other cells in BALF, comparing *CCR1*^+^ CD16+ monocytes with *CCR1*^-^ CD16+ monocytes. I) Boxplot showing an increase in cellular interactions with other cells in BALF for *CXCR6*^+^ memory CD8+T cells than *CXCR6*^-^ memory CD8+T cells. J) Predicted cellular interactions with other cells in BALF, comparing *CXCR6*^+^ memory CD8+T cells with *CXCR6*^-^ memory CD8+T cells. The circular size represents the significance of each ligand-receptor axis, and color represents the communication probability.

With regard to *CXCR6*^+^ memory CD8+T cells in PBMCs, the predicted cell-to-cell interactions showed a prominent elevation with increased severities of COVID-19 (P < 0.05, Figure 6D). Similar to *CCR1*^-^ CD16+monocytes, we observed no obvious difference of cellular interactions between normal controls and COVID-19 patients among *CXCR6*^-^ memory CD8+T cells (P > 0.05, Figure 6E). Compared with healthy individuals, *CXCR6*^+^ memory CD8+T cells demonstrated higher cellular communications with CD14+monocytes, CD34+progenitors, dendritic cells, effector CD8+T cells, naïve CD8+T cells, memory CD4+T cell, naïve CD4+T cells, NK, and megakaryocytes among severe patients (Supplemental Figure S20). There were 20 elevated cellular interactions of *CXCR6*^+^ memory CD8+T cells with other immune cells among severe patients, including *ADGRE5-CD55, ANXA1-FPR1, CCL3/CCL5-CCR1, CD99-CD99/PILRA, ICAM2-* (*ITGAL+ITGB2*), and *ITGB2-ICAM2/CD226* (Figure 6F). These cell adhesion molecules (*ANXA1* and *ICMA2*), cytokine binding and receptor activity genes (*CD44, CD36, CD74, CXCR4*, and *THBS1*), and inflammatory genes (*FPR1* and *SELL*) have been reported to be associated with COVID-19 [16, 55, 63, 64].

Among BALF cells, we also observed a remarkable increase in cellular interactions of *CCR1*^+^ CD16+monocytes and *CXCR6*^+^ memory CD8+T cells comparing to their corresponding negative cells (P < 0.001, Figure 6G-J and Supplemental Figure S21A). For example, enhanced ligand-receptor axes of *SELPLG-SELL, CCL5-CCR1, FN1-(ITGA4+ITGB1), CD99-CD99*, and *APP-CD74* among *CCR1*^+^ CD16+monocytes (Figure 6H), as well as *CXCL16-CXCR6, TNFSF14-TNFRSF14, ITGB2-CD226, CLEC2B/CLEC2C-KLRB1*, and *CCL3/CCL4-CCR5* among *CXCR6*^+^ memory CD8+T cells (Figure 6J). Notably, there was a 60% increase in cellular interactions between *CCR1*^+^ CD16+monocytes and epithelial cells compared with that of *CCR1*^-^ CD16+monocytes (Supplemental Figure S21B). We also found a 33.33% increase in the interactions between *CXCR6*^+^ memory CD8+T cells and epithelial cells compared with that of *CXCR6*^-^ memory CD8+T cells (Supplemental Figure S21C), such as enhanced ligand-receptor interactions including *TNF-TNFRSF1A, CXCL16-CXCR6*, and *CCL3-CCR5*. Previous studies [65, 66] have reported that the *CXCL16-CXCR6* axis modulates the localization of tissue-resident memory CD8+T cells to the lung airway. Overall, these results suggest that the increased cellular interactions with epithelial cells probably enhance the resident to the lung airway for against SARS-CoV-2 infection.

## Discussion

By using large-scale genetics data, we identified eight genomic loci including three novel loci (e.g., 1p22.2, 6p21.33, and 7p11.2) that were significantly associated with severe COVID-19. Other five loci including 3p21.31, 9q34.2, 12q24.13, 19p13.3, and 21q22.11 have been reported to be involved in COVID-19 risk in previous studies [23-28]. Notably, we prioritized 34 risk genes, including potential causal genes of *CXCR6, CCR1*, and *ABO*, to be associated with severe COVID-19. The CXC motif chemokine receptor 6 (CXCR6), which is a G protein-coupled receptor with seven transmembrane domains, regulates the partitioning of resident memory T cells by recruiting lung tissue-resident memory CD8+T cells to airways [65]. *CCR1* gene encodes the CC motif chemokine receptor 1 (CCR1) belonging to a member of the beta chemokine receptor family. Several previous GWASs have reported genetic variants in *CCR1* are associated with COVID-19 susceptibility at a genome-wide significant level [25, 27]. For the *ABO* gene, it encodes protein relevant to the ABO blood group system. Both genetic and non-genetic studies [25, 27, 67] have showed the involvement of *ABO* gene in COVID-19 susceptibility, while the *ABO* gene encodes protein that is relevant to the ABO blood group system, and it was also notably associated with several thrombotic and coagulation-related traits including deep vein thrombosis and pulmonary heart disease, which have been reported to be risk factors and sequalae to severe COVID-19 [68, 69].

Understanding the immune responses of monocytes and memory T cells is fundamental to the rational design of innovative and effective strategies to develop better vaccines [70, 71], and contributes to reveal the pathogenesis of severe COVID-19 [29]. Our current analyses reveal that host genetic determinants have a prominent influence on the immune responses of CD16+monocytes, megakaryocytes, and memory CD8+T cells to severe COVID-19. Previous studies [11, 29, 54] showed that the influence caused by monocytes and megakaryocytes in inflammatory storms is noteworthy among severe COVID-19 patients. We found that *CCR1*^*+*^ CD16+monocytes and *ABO*^+^ megakaryocytes showed a significantly increased propensity to cause inflammatory storms among severe patients. The observations suggest highly-expressed interferon-related genes, including *S100A8, S100A9, S100A12, CD14, CXCL8, IGSF6, IRF3, IFI6, IFITM1*, and *IFITM3* among the two cell subsets contribute to exacerbate inflammation among severe patients. The inflammatory mediator of EN-RAGE encoded by *S100A12* was significantly correlated with COVID-19 [21], and *S100A8, S100A9, IRF3, IFI6, IFITM1*, and *IFITM3* have been reported to elicit autoinflammatory and autoimmune conditions in response to SARS-CoV-2 infection [10, 16, 29, 57, 58]. Double positive CD14+CD16+monocytes reported as tissue-infiltrative cells have a higher potency of antigen presentation and highly-expressed proinflammatory cytokines [72, 73]. Additionally, interferons are the mediators in several canonical host antiviral signaling to activate the expression of numerous required molecules of the early response to viral infection [74], and impaired type I interferon activity play important roles in severe COVID-19 [58]. Our findings described above suggest that *CCR1*^***+***^ CD16+monocytes and *ABO*^+^ megakaryocytes as a functional subset of myeloid cells convey higher risks to severe COVID-19.

Memory CD8+T cells could elicit improved immunological features that are critical in host protection from viral infectious [71]. After influenza virus infections, memory CD8+T cells reside in the lung for a couple of months and these resident memory T cells are necessary for effective immunity against secondary infection [75]. Among severe COVID-19 patients, we found that *CXCR6*^*+*^ memory CD8+T cells undertook several improved immunological features, including higher scores of cytokine, chemokine, T cell activation, proliferation, and migration, which suggests *CXCR6*^*+*^ memory CD8+T cells potentially contribute to the protection of SARS-CoV-2 infection. Among these positive *CXCR6*^+^ cells, numerous highly-expressed cytokine and chemokine genes, including *CCR1, CCR2, IFNGR1*, and *MYC*, may work on activating memory T cells. Earlier evidence indicated that MYC was rapidly but temporally induced during the early stage of T cell activation [76]. The *CCR1* plays a pivotal role in the recruitment of effector immune cells to the site of inflammation, and the pharmacologic inhibition of this gene may suppress immune hyper-activation in severe COVID-19 [15]. Memory CD8+T cells obtained the capability of transforming to effector cells by sensing inflammation from monocytes [71]. Thus, inflammatory *CCR1*^*+*^ CD16+monocytes among severe COVID-19 patients potentially accelerate the activation of memory CD8+T cells.

Additionally, we observed a prominent decrease of the cell proportion of *CXCR6*^*+*^ memory CD8+T cells among severe patients. This decrease in peripheral blood among severe patients is probably due to efflux to the site of viral infected lung tissue in answer to ongoing tissue damage. Earlier studies [29, 77] have reported that functional CD8+T cell subsets manifest a notable decrease in the peripheral blood of severe COVID-19 patients. Epithelium is the most vulnerable tissue to be attacked by viral or microbial infection, thus the presence of resident memory CD8+T cells are imperative for defending the debilitating infections for hosts [75]. In the current study, we found an obvious increase in cellular interactions of *CXCR6*^+^ memory CD8+T cells with epitheliums. Enhanced ligand-receptor interactions including *TNF-TNFSFRSF1A, CXCL16-CXCR6*, and *CCL3-CCR5* may contribute to the lung-residence of memory CD8+T cells. Previous evidence demonstrated a major role for *CXCL16-CXCR6* interactions in regulating the resident of virus-specific memory CD8+T cells [65, 66]. An earlier study showed a stronger interactions between epithelial and immune cells among severe COVID-19 cases than that among moderate cases [15]. We demonstrated that *CXCR6*^+^ memory CD8+T cells mounted highly effective immune responses to against COVID-19, highlighting the remarkable biological plasticity in subsets of memory CD8+T cells differentiation.

The power of this study is limited by the lack of matched genetic data and scRNA-seq data in each sample for uncovering the genetic effects on immune cells for severe COVID-19. To reduce the influence of this limitation, we adopted a widely-used approach by integrating a large-scale GWAS summary statistics with enormous amount of single cell sequencing data, as referenced in previous studies [45, 78]. Based on our findings suggesting that host genetic components exert regulatory effects on immunological dysregulations for SRAS-CoV-2 infection, more studies are warranted for exploring the genetic modification of peripheral T cells to defend against lethal severe COVID-19.

## Conclusions

In sum, we provide comprehensive insights that host genetic determinants are fundamental in influencing the peripheral immune responses to severe COVID-19. Both *CCR1*^*+*^ CD16+monocytes and *ABO*^*+*^ megakaryocytes contribute higher risk to the inflammatory storms among severe patients. *CXCR6*^+^ memory CD8+T cells exhibit a notable polyfunctionality of several improved immunologic features implicated in the etiology of severe COVID-19. Further experiments to parse the molecular mechanism of these three cell subpopulations on severe COVID-19 patients are crucial for promoting personalized protective immunity.

## Data Availability

All the GWAS summary statistics used in this study can be accessed in the official websites (www.covid19hg.org/results). The GTEx eQTL data (version 8) were downloaded from Zenodo repository (https://zenodo.org/record/3518299#.Xv6Z6igzbgl). Three scRNA-seq datasets were downloaded from the GEO database (https://www.ncbi.nlm.nih.gov/gds/?term=GSE149689 and https://www.ncbi.nlm.nih.gov/gds/?term=GSE150861) and the ArrayExpress database (https://www.ebi.ac.uk/arrayexpress/experiments/E-MTAB-9357). All analysis code in the Methods is available in an online GitHub repository at https://github.com/mayunlong89/COVID19_scRNA.

## Abbreviations

COVID-19: coronavirus disease 2019
SARS-CoV-2: severe acute respiratory syndrome coronavirus 2
GWAS: genome-wide association study
scRNA-seq: single cell RNA sequencing
PBMCs: peripheral blood mononuclear cells
BALF: bronchoalveolar lavage fluid
eQTL: expression quantitative trait loci
GEO: the Gene Expression Omnibus database
WHO: the World Health Organization
SNP: single nucleotide polymorphism
OR: odds ratio
MAF: minor allele frequency
QQ: quantile-quantile
MAGMA: Multi-marker Analysis of GenoMic Annotation
LD: linkage disequilibrium
FDR: false discovery rate
KEGG: the Kyoto Encyclopedia of Genes and Genomes
PPC: the Pearson correlation coefficient
MDS: multidimensional scaling
OTG: the Open Target Genetics
PPI: protein-protein interaction
up-DEG: significantly up-regulated expression gene associated with COVID-19

## Declarations

## Acknowledgments

We appreciate Prof. Yang Jian from Westlake University for providing helpful suggestions, and our appreciation also goes to all the authors from the COVID-19 Host Genetic Consortium who have deposited and shared GWAS summary data on public databases and goes to the authors who publicly released the scRNA-seq datasets on PBMC and BALF with distinct COVID-19 severities.

## Authors’ contributions

J.S., and Y.M. conceived and designed the study. Y.M., F.Q., C.D., J.L., Y.K.H., Y.R.Z., Y.X., Y.G.Z., and Y.H.Y. contributed to management of data collection. Y.M., F.Q., C.D., Y.K.H., and J.L. conducted bioinformatics analysis and data interpretation. Y.M., J.S., Z.W. and J.Q. wrote the manuscripts. All authors reviewed and approved the final manuscript.

## Ethics approval and consent to participate

Not applicable

## Consent for publication

Not applicable

