## Supplemental Figures for "Single cell sequencing analysis uncovers genetics-influenced CD16+monocytes and memory CD8+T cells involved in severe COVID-19"

### 1. *Supplemental Methods*

**Supplemental Method S1. Prioritization of candidate causal genes for eight identified genomic loci**

To further explore the causal effects of nine index SNPs in eight genomic loci on severe COVID-19, we performed a prioritization analysis by leveraging the Open Target Genetics (OTG, https://genetics.opentargets.org/) tool (*1*). The OTG tool is a web-access integrative resource that aggregates human GWAS summary statistics, including UK Biobank data and NHGRI-EBI GWAS catalog, and functional genomic data, including protein quantitative trait loci data (pQTLs), expression QTL (eQTL) from eQTL Catalogue, eQTLGen and GTEx, and promoter capture hiC (PCHI-C) from 27 different cell types, and DNase I hypersensitive site (DHS)-gene promoter correlation and other datasets from a wide range of cell types and tissues. Based on the Variant2Gene (V2G) algorithm, for the index SNP rs2166172 in 1p22.2, we prioritized *BARHL2* as a candidate causal gene for severe COVID-19. With the highest V2G score, *CCHCR1* was highlighted as candidate causal gene for the index SNP rs143334143 in 6p21.33, *VSTM2A* as candidate causal gene for rs622568 in 7p11.2, *OAS1* as candidate causal gene for rs2269899 in 12q24.13, *DPP9* as candidate causal gene for rs2109069 in 19p13.3, and *IFNAR2* as candidate causal gene for rs13050728 in 21q22.11. The index SNP of rs13050728 ($P = 1.91\times10^{-11}$) in 21q22.11 is highly LD with the reported SNP of rs9976829 ($R^2 = 1$) (*2*) and rs2236757 ($R^2 = 0.8266$) (*3*). The index SNP of rs2269899 in 12q24.13 ($P = 3.24\times10^{-8}$) shows high LD with the reported SNP of rs10735079 ($R^2 = 0.9282$) (*3*).

**Supplemental Method S2. Combination of GWAS summary statistics with scRNA-seq data (dataset #1) by using the MAGMA algorithm**

As an independent technical approach to validate genetics-related peripheral immune cells in PBMCs implicated in severe COVID-19 identified from the RolyPoly algorithm, we applied a generalized linear regression (GLR) model in the MAGMA (v1.06, https://ctg.cncr.nl/software/magma) (*4*). Conditioning on the 10% most specific genes from the single cell data for each cell type, we examined gene-level genetic associations of severe COVID-19-associated immune cell types by adding these variables as covariates for the linear regression model. The specificity of each gene for each cell type was calculated by using the mean gene expression of cell type divided the sum of mean gene expression across all cell types. A window of 50 kb upstream to 50 kb downstream for a given gene coordinates was set to calculate gene-level association statistics. The 1,000 Genome Project European Phase 3 panel (*5*) was used as the reference panel for both methods. We restricted the GLR analysis to SNPs in the autosomes, and any SNPs with minor allele frequency < 5% were excluded. The major histocompatibility complex region (Chr6: 25-35 Mbp) was also excluded due to the extensive linkage disequilibrium (LD) in this region.

#### *2. Supplemental Figures*

**Supplemental Figure S1. UMP projections of cells in PBMCs from normal controls, mild, moderate, and severe COVID-19 patients by using the Seurat R package (dataset #1).** Dark green represents normal controls, light green represents mild patients, orange represents moderate patients, and red represents severe patients.

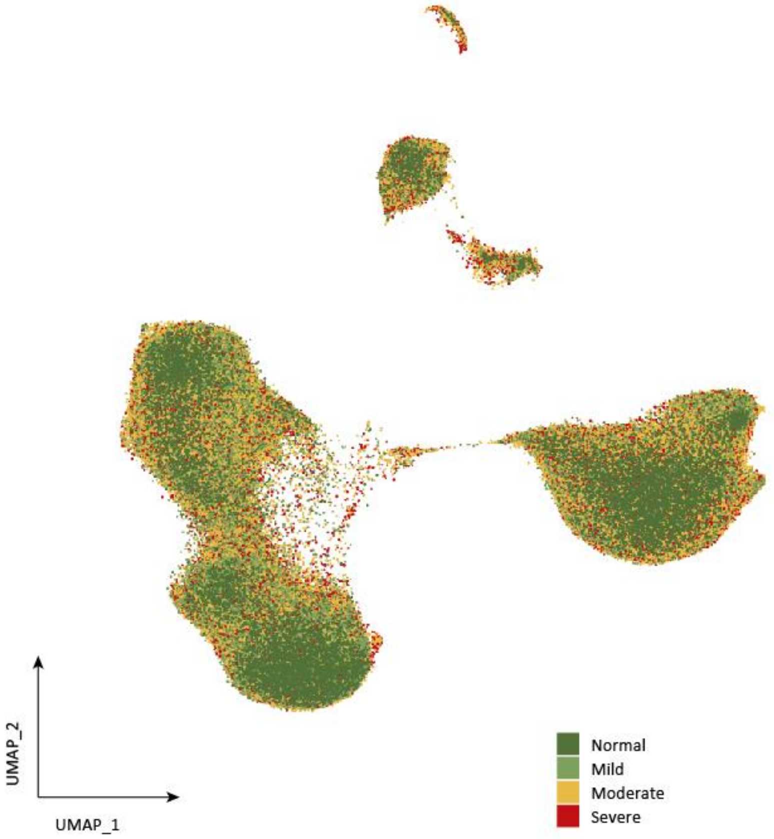

**Supplemental Figure S2. Heatmap showing levels of well-known marker genes specific for each cell type in PBMCs.** Color legend represents the expression level of each marker gene.

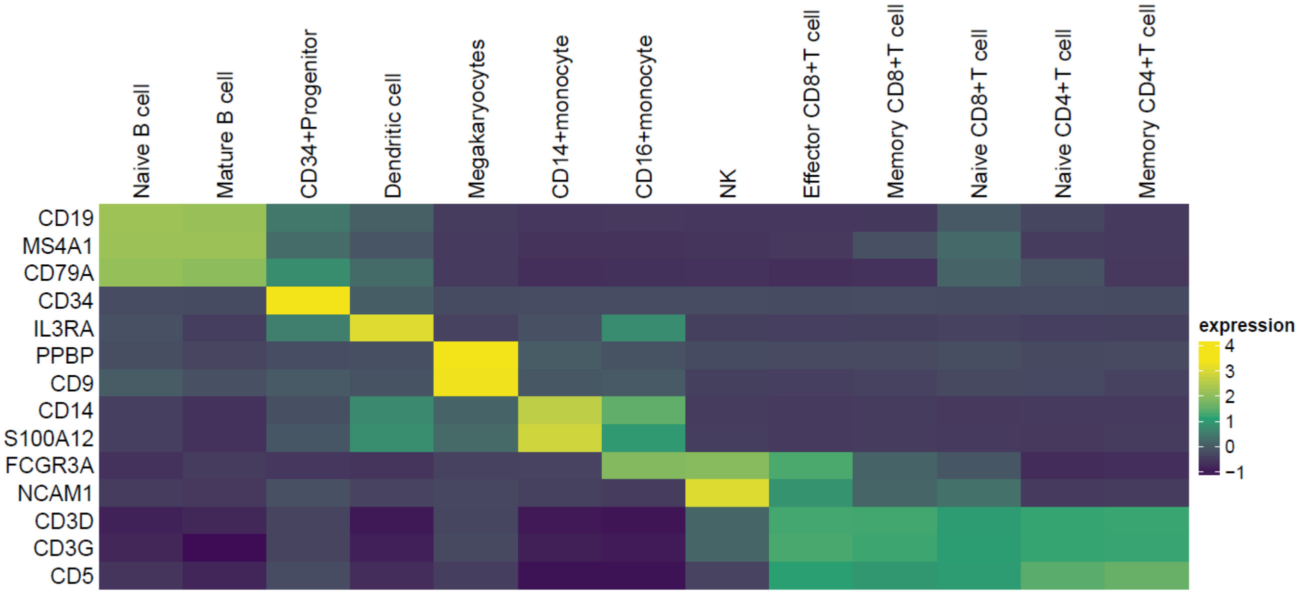

**Supplemental Figure S3. Single-cell transcriptomes of PBMCs from normal controls, mild, moderate, and severe COVID-19 patients.** A) UMAP projections of 514,400 cells from normal controls, mild, moderate, and severe patients colored by annotated cell types. There was 13 clusters with 13 distinct cell types annotated by using well-known markers. B) Normalized expression of marker genes on a UMAP plot. For example, the well-known markers of CD14, NKG7, PRF1, GNLY, GZMB, LEF1, CD4, CD8A, GZMK, FCGR3A, and NR4A1. C) Proportion of cell types among normal controls, mild, moderate, and severe patients. D) Violin plots of selected markers (lower row) for 13 cell subpopulations. The left column shows the cell types annotated by combinations of markers.

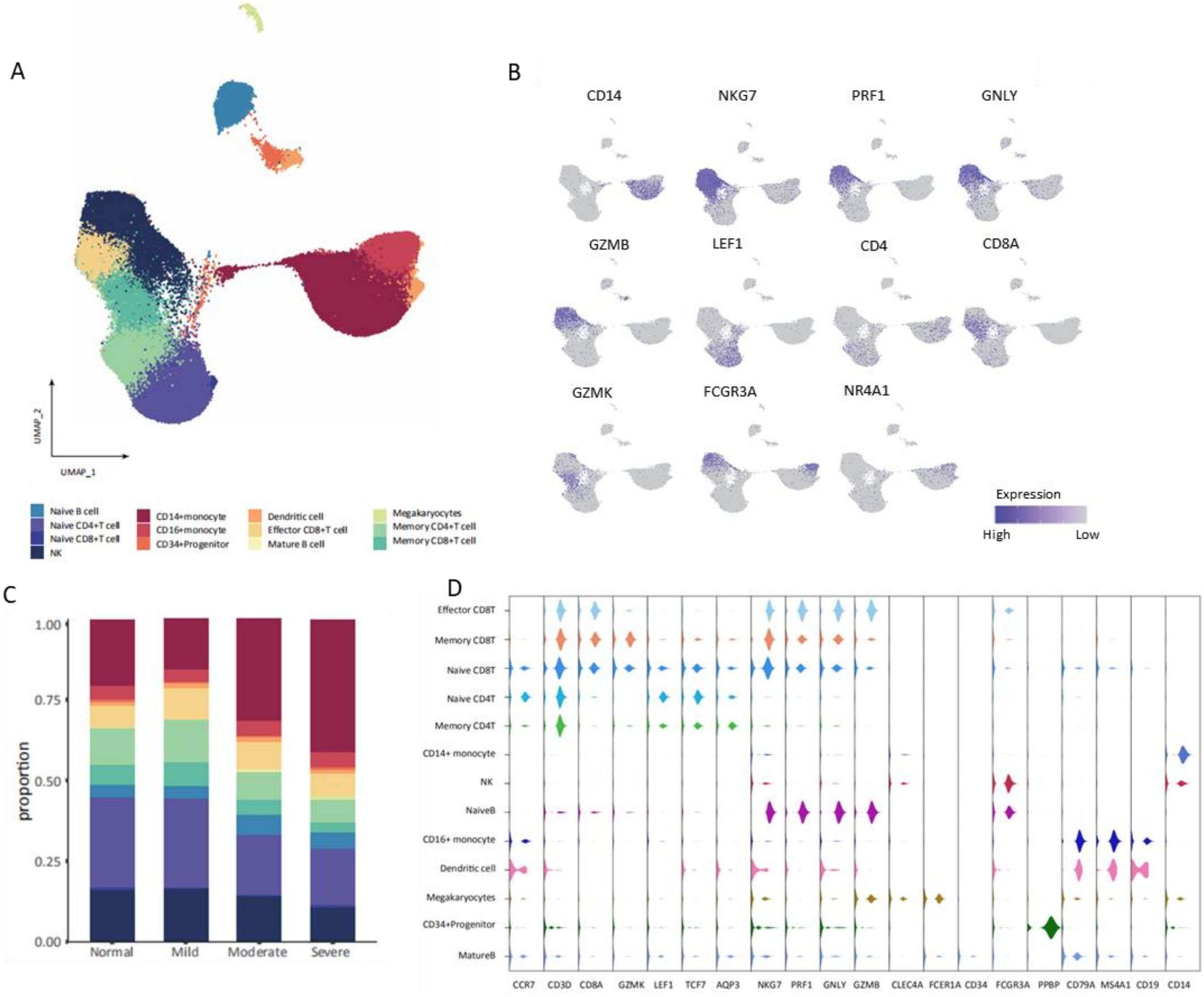

**Supplemental Figure S4. Hierarchical clustering using the PCC of a normalized transcriptome between diseases (normal and patients with different COVID-19 severities) in cell type resolution (n = 13).** The color intensity of the heatmap represents the PPC values. The color bars above the heatmap shows the disease group and cell type. This analysis was based on the largest scRNA-seq dataset of dataset #1 (accession number: E-MTAB-9357).

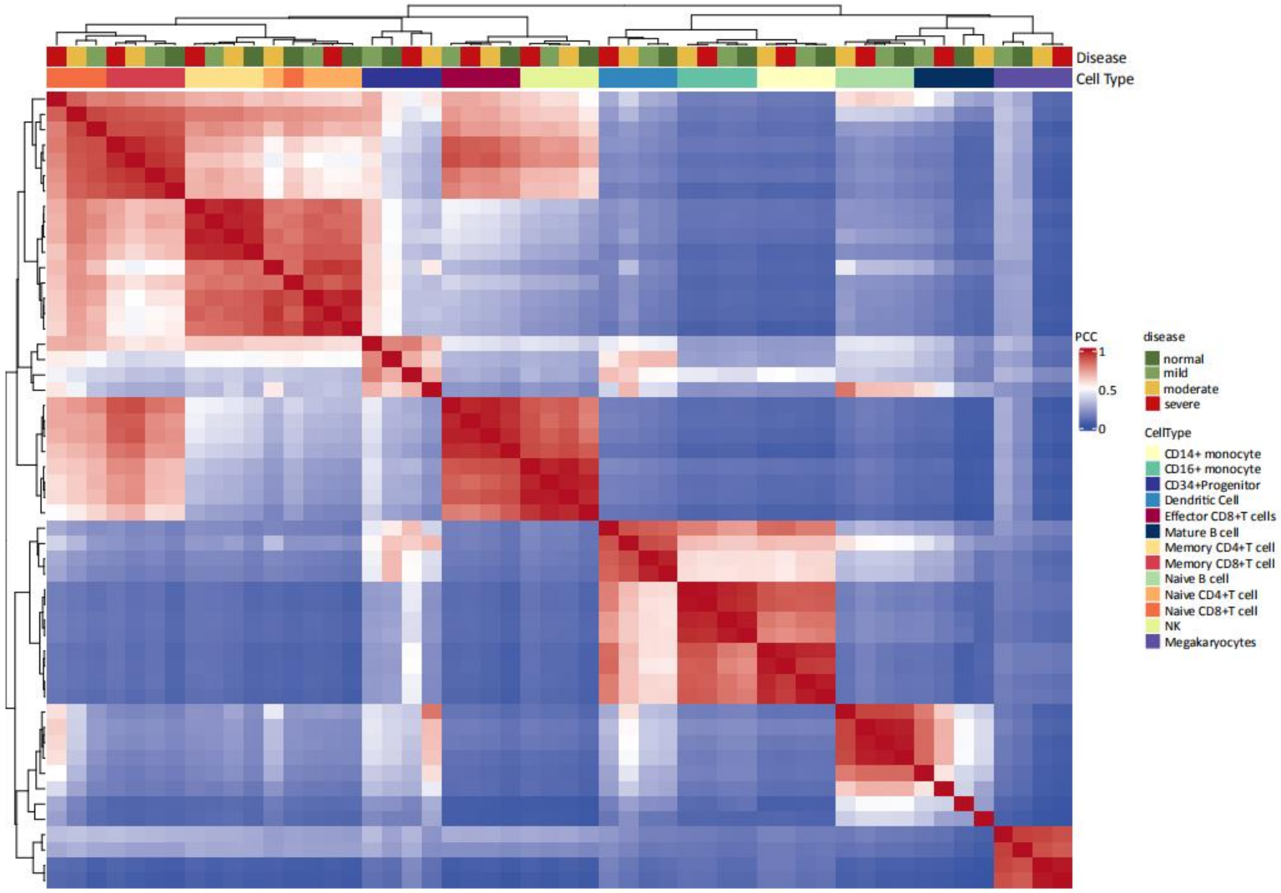

**Supplemental Figure S5. Boxplots showing percentages of each cell type for PBMCs in donors from healthy control and COVID-19 patients with different severities based on the combined dataset.** The statistical analysis for calculating difference among different groups was based on the Wilcoxon sum-rank test.

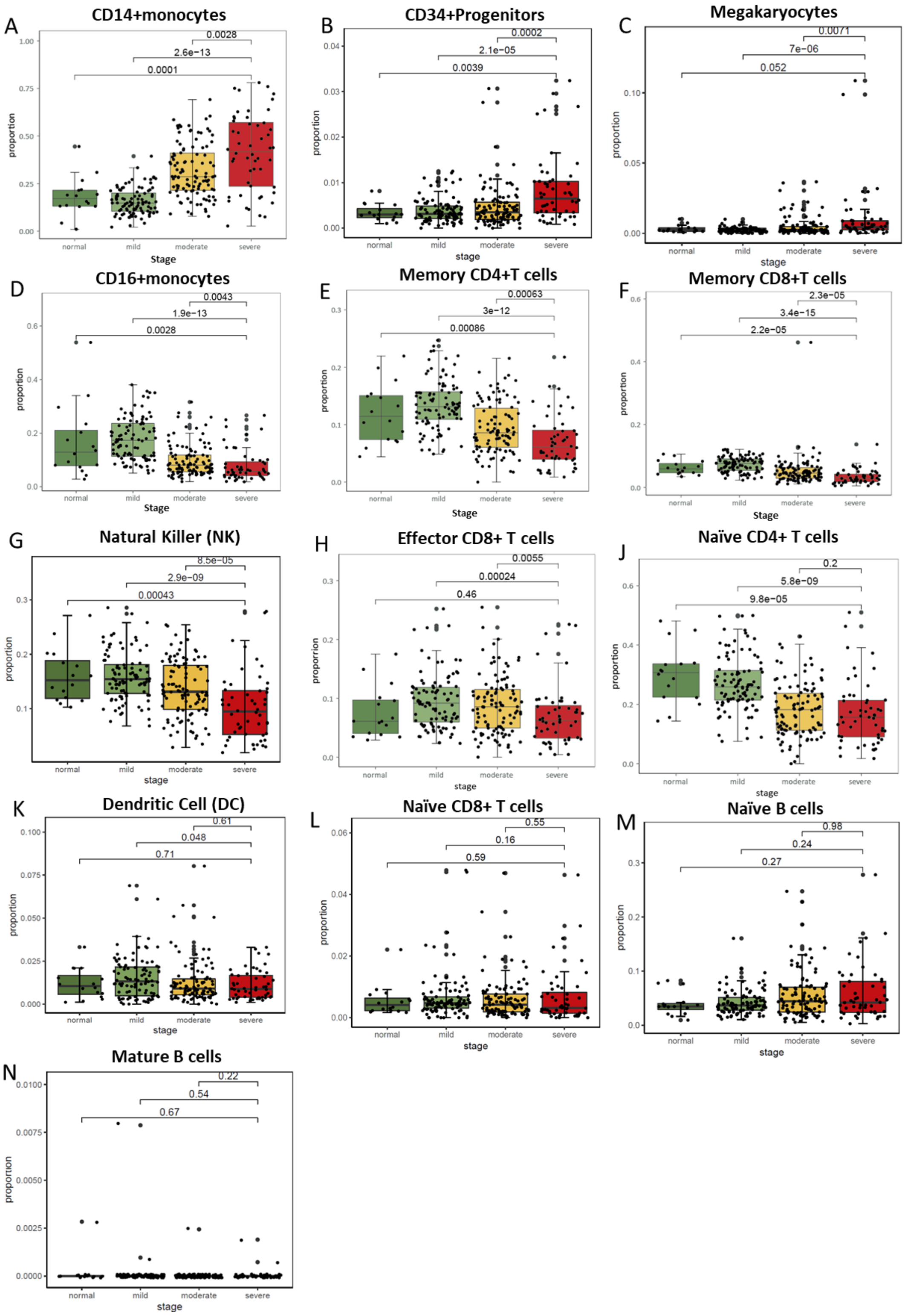

**Supplemental Figure S6. Regional association plots for severe COVID-19-associated genetic loci based on meta-GWAS summary data.** A-F) 1p22.2, 6p21.33, 7p11.2, 12q24.13, 19p13.3 and 21q22.11. The purple diamond marks the most strongly associated SNP in each locus with severe COVID-19. The color illustrates LD information with the given SNP, as shown in the color legend. The detailed information is shown in Supplemental Table S3.

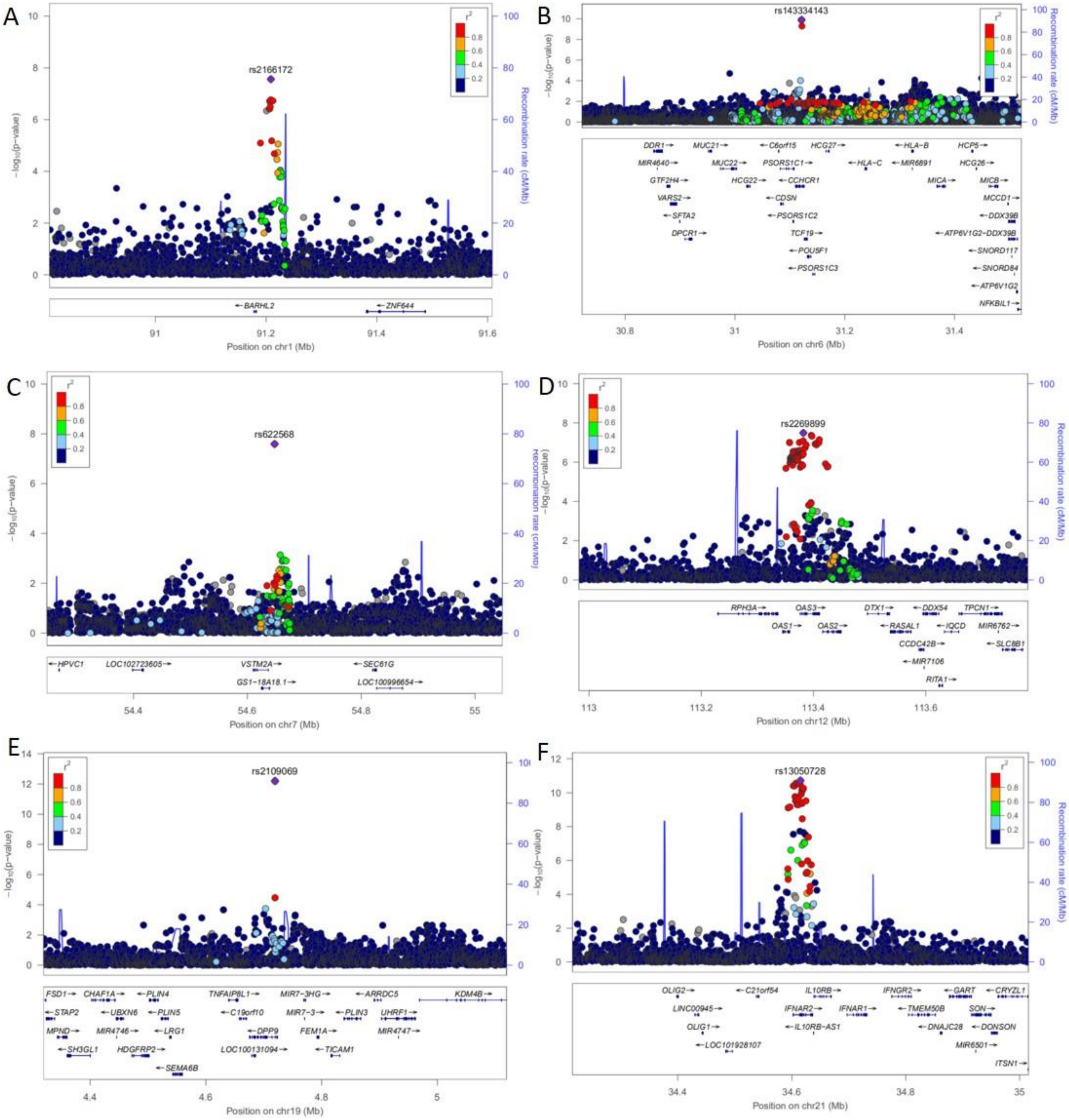

**Supplemental Figure S7. Regional association plots for severe COVID-19-associated genetic loci based on meta-GWAS summary data.** A-B) Two independent genetic association signals in the 3p21.31 loci (rs35081325, P = $3.32 \times 10^{-58}$, and rs33998492, P = $3.59 \times 10^{-14}$, respectively). The purple diamond marks the most strongly associated SNP in each locus with severe COVID-19. The color illustrates LD information with the given SNP, as shown in the color legend. C) The results of calculating the LD information between rs35081325 and rs33998492 using the LDpair Tool based on the European population (EUR+TSI). D) Regional association plot for the 9q34.2 locus associated with severe COVID-19.

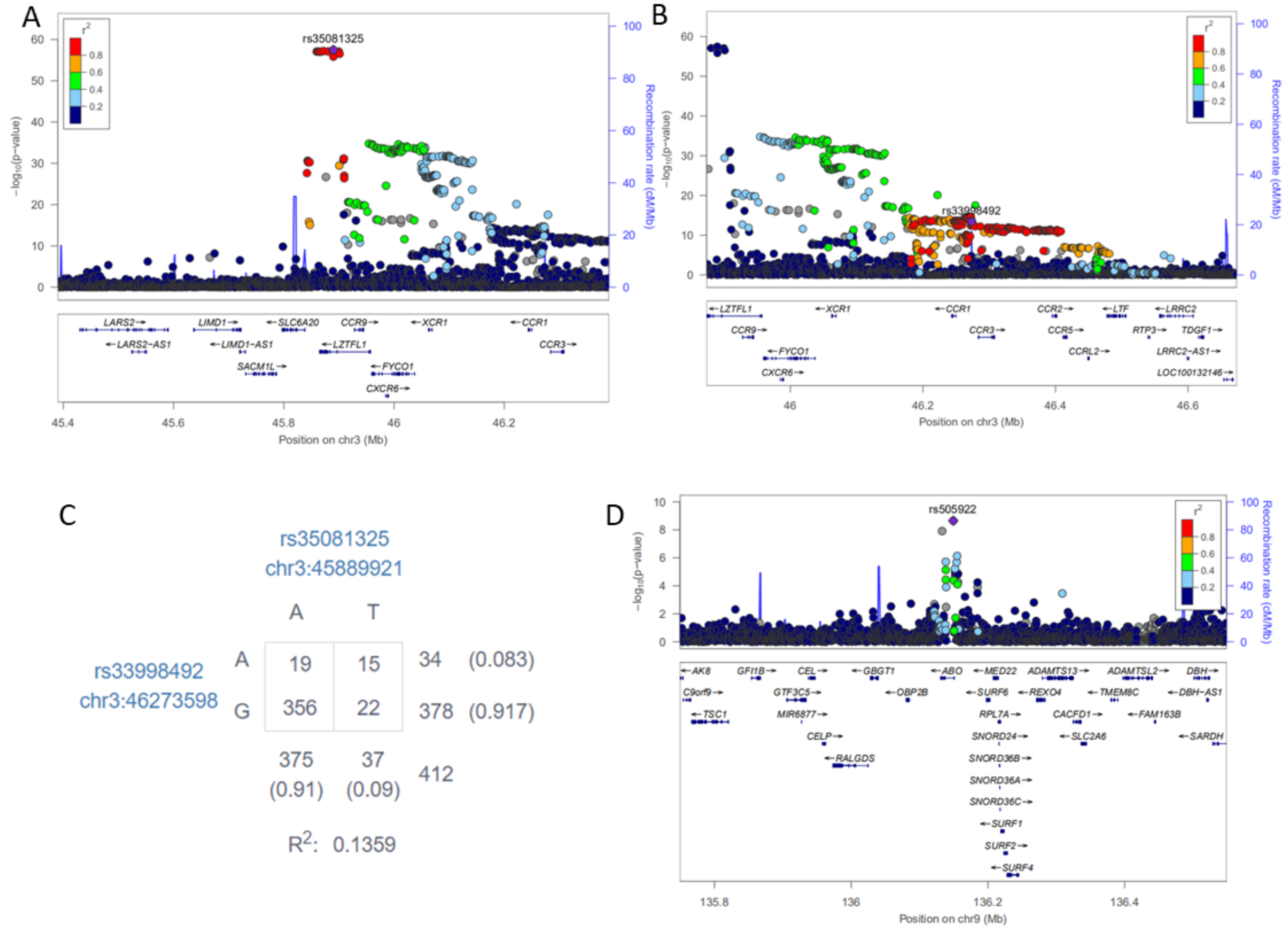

**Supplemental Figure S8. Circus plot showing the results of MAGMA-based gene-level association analysis.** A for lung tissue, and B for blood. The inner ring demonstrates the 22 autosomal chromosomes (Chr1-22) and X chromosome. In the outer ring, a circular symbol stands for a specific gene, and color marks the significant level of the gene. Red color marks genes significantly associated with severe COVID-19 with FDR < 0.05, orange color indicates genes suggestively associated with severe COVID-19 with $6.96\times10^{-5} \leq P < 0.001$, light blue color marks suggestive genes with $0.001 \leq P < 0.05$, and dark blue indicates genes have non-significant associations ($P > 0.05$).

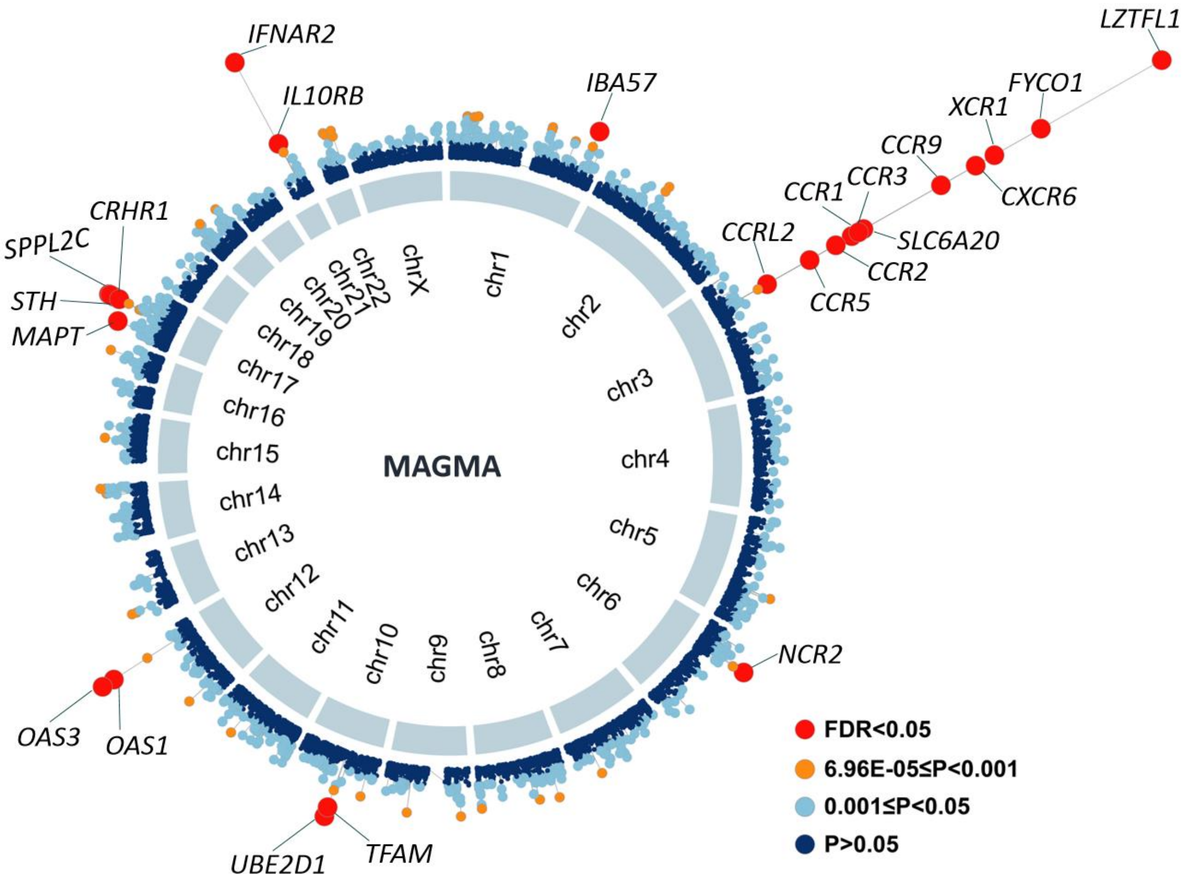

**Supplemental Figure S9. The 19 biological pathways enriched from the MAGMA-based pathway enrichment analysis.** A) Barplot showing the 19 biological pathways based on the KEGG pathway. B) Multidimensional scaling plot for clustering the 19 biological pathways based on their Jaccard distance (see the Methods). Color represents gene number in each pathway, and circular ring size indicates the Z score of enrichment for each pathway. The detailed information is shown in Supplemental Table S5.

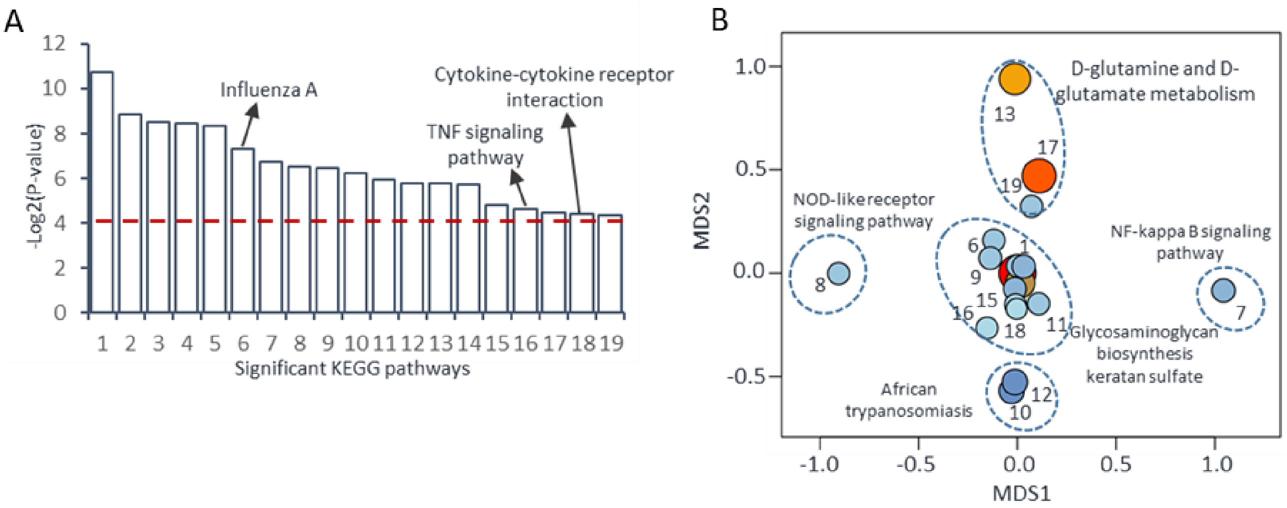

**Supplemental Figure S10. High consistence results between MAGMA and S-MultiXcan analysis.** A) Venn diagram exhibiting the overlapped significant genes between MAGMA and S-MultiXcan analysis. B) Correlation of significant risk genes identified frim MAGMA and S-MultiXcan analysis.

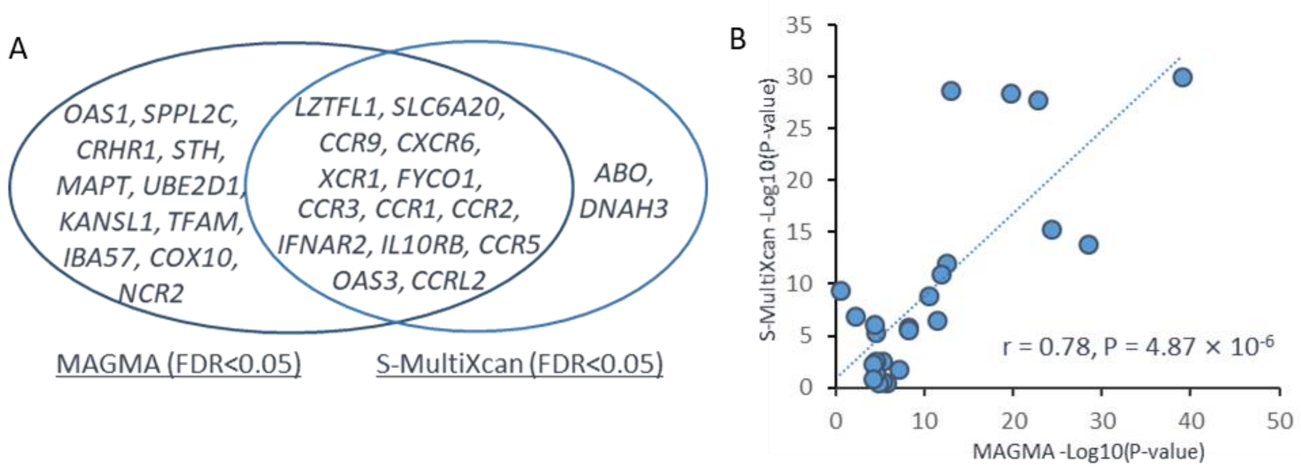

**Supplemental Figure S11.** ***In silico*** **permutation analysis of 100,000 times of random selections.** A) The comparison of the top-ranked genes from MAGMA gene-based analysis ($P < 0.05$) with that from S-MultiXcan-based analysis ($P < 0.05$). B) The comparison of the top-ranked genes from MAGMA gene-based analysis ($P < 0.05$) with that from S-PrediXcan-based analysis based on lung tissue ($P < 0.05$). C) The comparison of the top-ranked genes from MAGMA gene-based analysis ($P < 0.05$) with that from S-PrediXcan-based analysis based on blood tissue ($P < 0.05$).

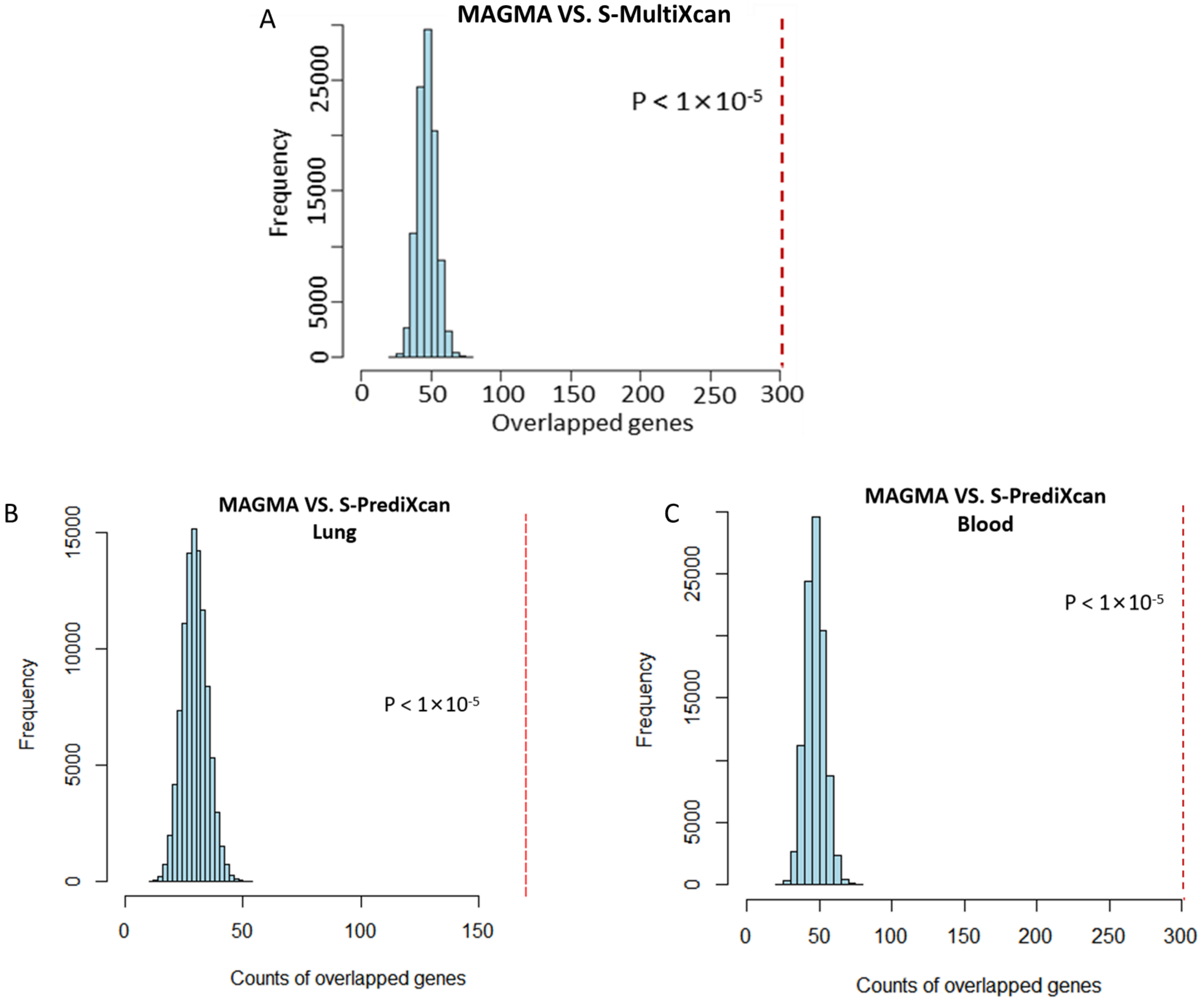

**Supplemental Figure S12. Multiple independent approaches identify genetics-relevant risk genes associated with severe COVID-19.** A) Venn diagram showing the overlapped genes among SNP-level association analysis, MAGMA gene-based analysis, S-MultiXcan analysis, and S-PrediXcan analysis based on lung and blood. B) Summary of total 34 genetically risk genes associated with severe COVID-19.

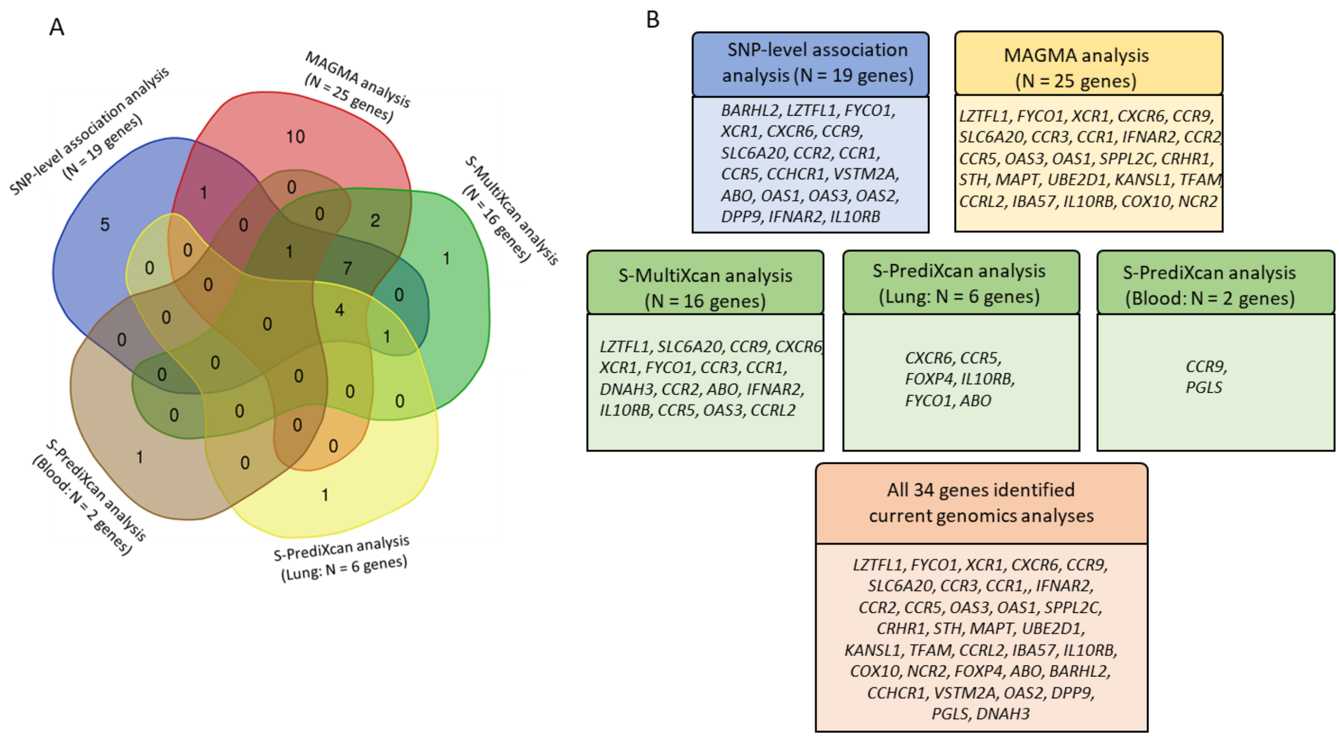

**Supplemental Figure S13. The 10 biological pathways significantly enriched by 34 risk genes based on the KEGG database.** A) Barplot showing these 10 biological pathways associated with severe COVID-19. The Arabic numeral in each bar represents the ID of each pathway ordered by the significant level (Supplemental Table S10). B) Multidimensional scaling plot for clustering these 10 pathways based on the Jaccard distance (see Methods). Color represents the significance of each pathway (red color marks the significant pathways with lowest P values).

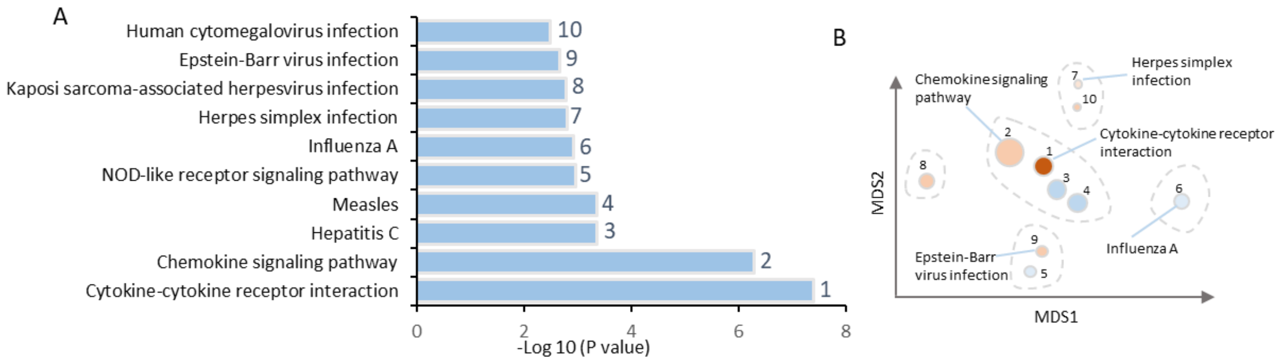

**Supplemental Figure S14. Plot of gene-drug interaction analysis for 34 risk genes.** The gene-drug information was downloaded from the STITCH database (http://stitch.embl.de/). The orange node represents the risk gene, and the light green color stands for the targeted drugs. For more detail information, see the method for gene-drug interaction analysis.

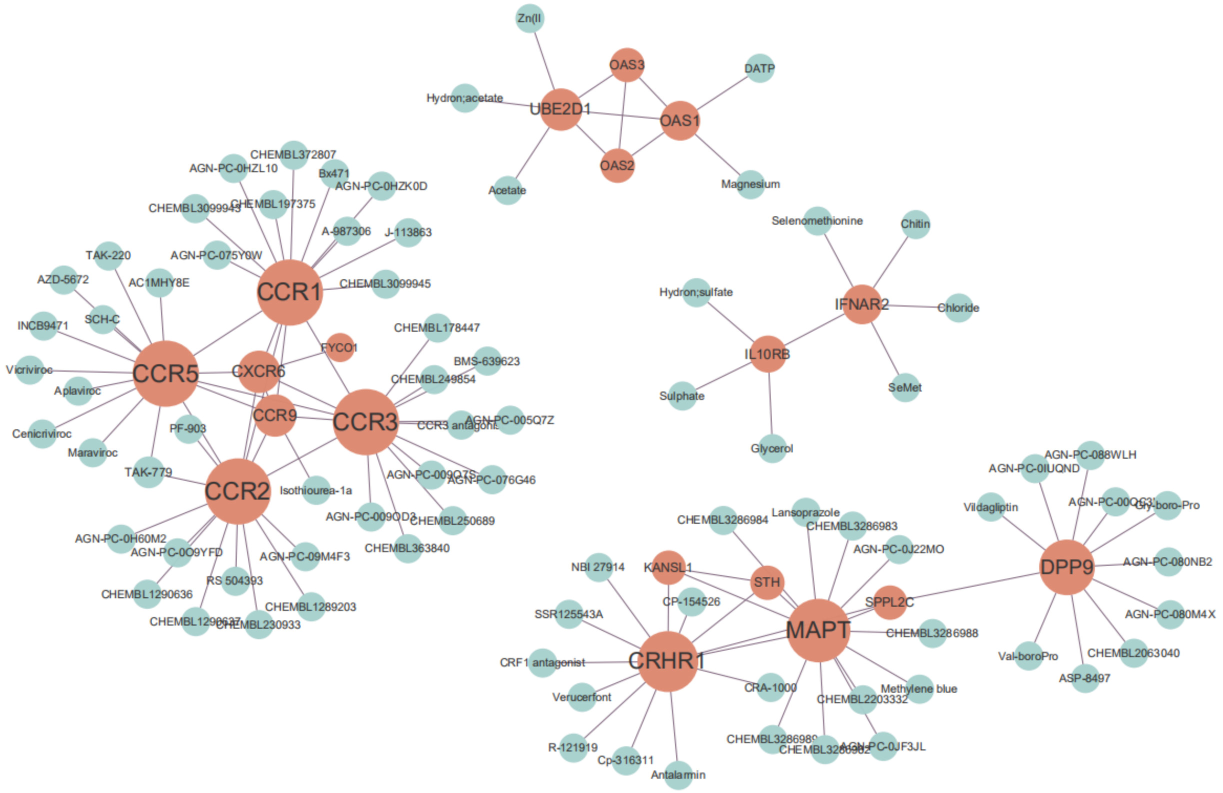

**Supplemental Figure S15. Genetics-risk genes influenced three immune cell subsets for severe COVID-19.** A) Plot showing the specific genes expressed in three identified immune cell subsets for severe COVID-19. The most specificity gene for each cell type is *CCR1* for CD16+monocytes, *CXCR6* for memory CD8+T cells, and *ABO* for megakaryocytes. B) Dot plot showing the expressed percent of three risk genes of *CXCR6*, *CCR1*, and *ABO* in each peripheral cell type in PBMCs among severe patients based on the scRNA-seq dataset #1 (E-MTAB-9357). Dot size represents fraction of cells within cell type expressing a given gene, and color intensity represents binned count-based expression amounts (log(scaled UMI +1)) among expressing cells.

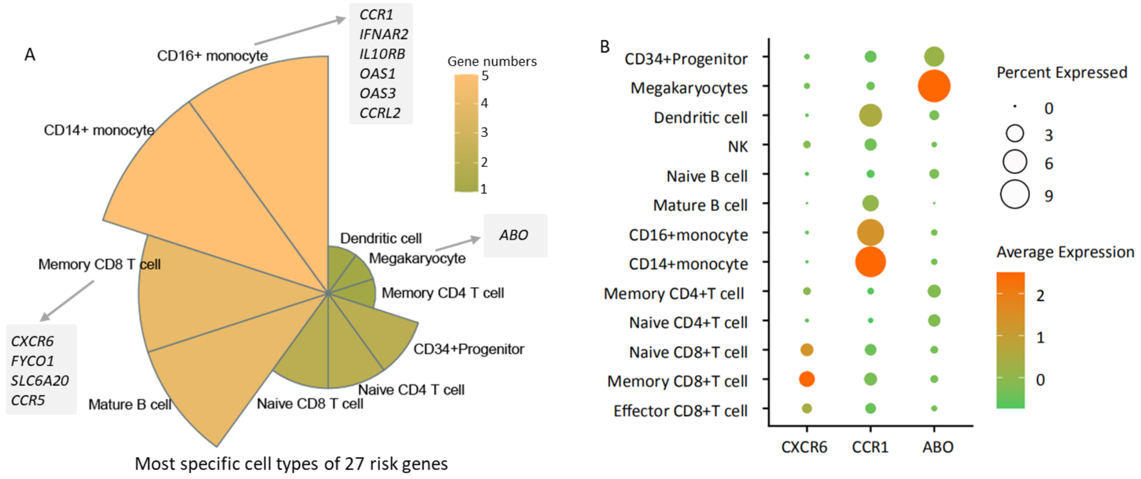

**Supplemental Figure S16. Dot plot showing the expressed percent of three risk genes of *CXCR6*, *CCR1*, and *ABO* in each peripheral cell type in PBMCs among severe patients based on two scRNA-seq dataset of #2 (GSE149689) and #3 (GSE150861)**. Dot size represents fraction of cells within cell type expressing a given gene, and color intensity represents binned count-based expression amounts (log(scaled UMI +1)) among expressing cells.

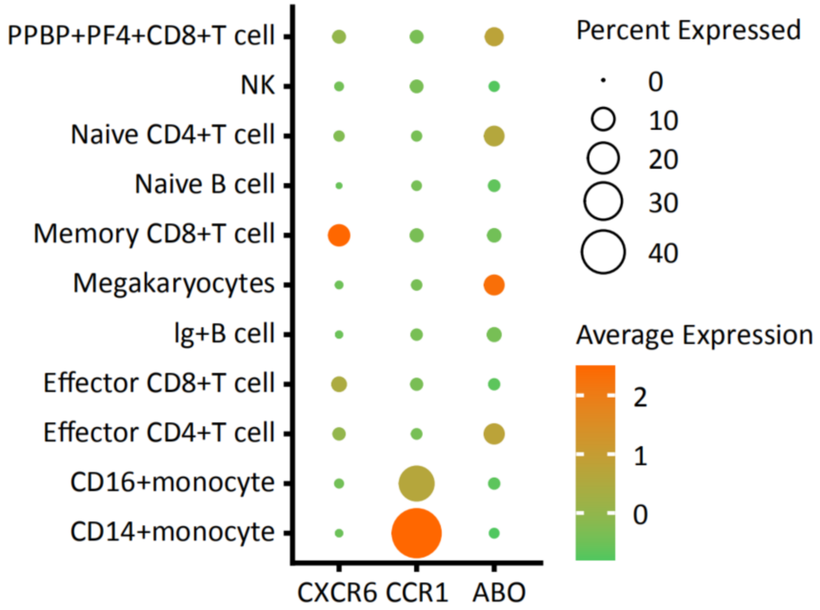

**Supplemental Figure S17. *CCR1*$^{+}$ CD16+monocytes showing higher risk to cytokine storms among COVID-19 patients.** A) Boxplot showing the difference of pathway activity score of both cytokine-cytokine receptor interaction and chemokine signaling pathway between *CCR1*$^{+}$ and

*CCR1*⁻ CD16+monocytes. Two-side Wilcoxon sum-rank test was used to calculate the significance. B)-C) Barplot showing the proportion of significant druggable proteins (B) and significant druggable proteins associated with COVID-19 (C) between *CCR1*$^{+}$ and *CCR1*$^{-}$ CD16+monocytes. The hypergeometric test was applied to calculate the significance. D) Venn plot exhibiting the overlapped up-DEGs between pairwise comparisons of mild vs. normal, moderate vs. normal, and severe vs. normal. E) Functional enrichment analysis based on GO biological process for 190 up-DEGs. F) Gene-drug interaction analysis for 190 up-DEGs. G)-L) Representative up-DEGs among *CCR1*$^{+}$ CD16+monocytes showing significantly elevated expressions with increased COVID-19 severities. G) *IRF3*, H) *CXCL8*, I) *CD14*, J) *S100A12*, K) *IFI6*, and L) *IGSF6*.

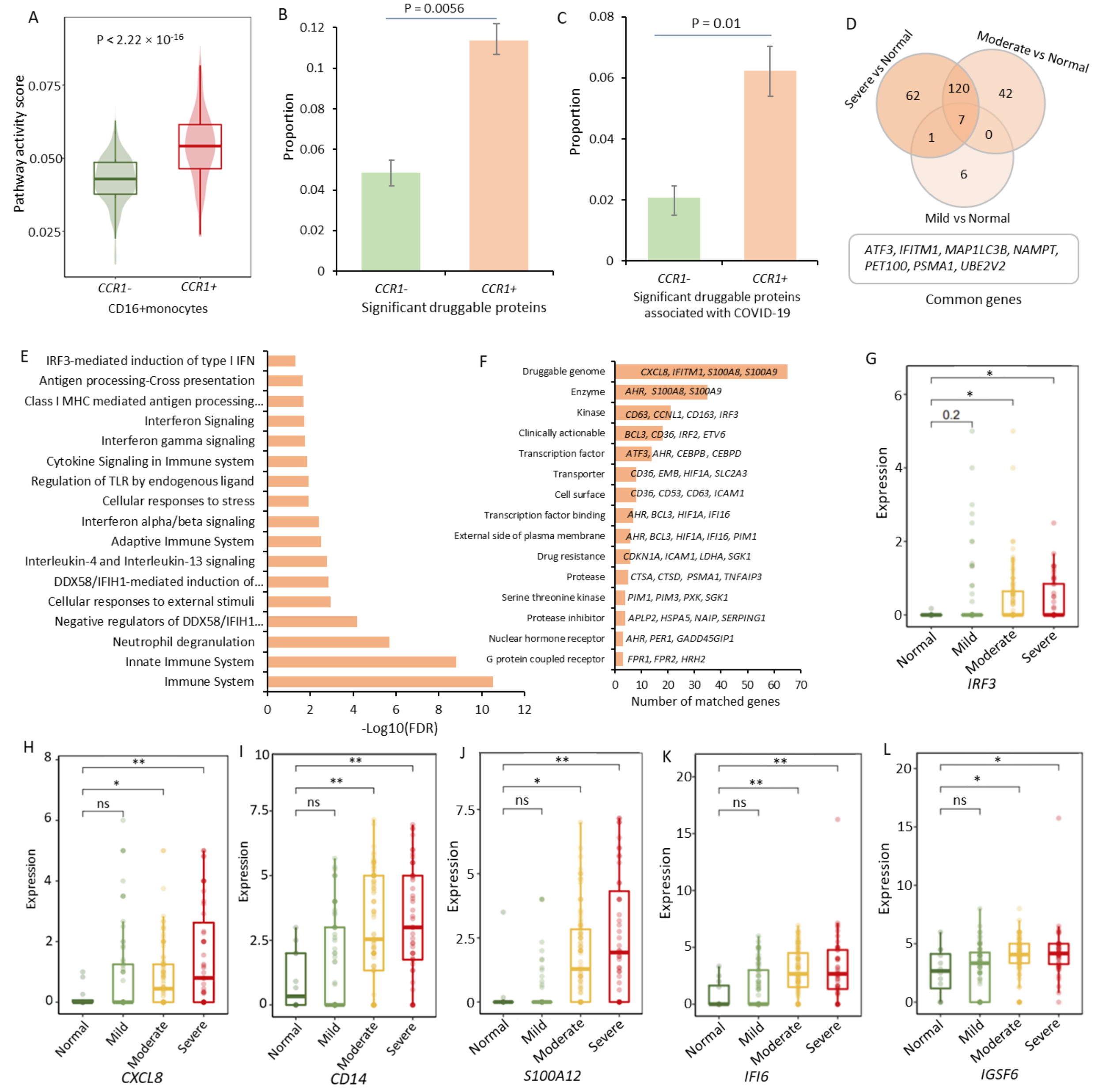

**Supplemental Figure S18. *ABO*$^{+}$ megakaryocytes contribute higher risk to cytokine storms among severe COVID-19 patients.** A)-B) Boxplot showing the difference of inflammatory cytokine score (A) and pathway active score (B) between *ABO*$^{+}$ and *ABO*$^{-}$ megakaryocytes. There were two pathways of cytokine-cytokine receptor interaction and chemokine signaling pathway used. Two-side Wilcoxon test was applied. C) Volcano plot showing differentially expressed genes between *ABO*$^+$ and *ABO*$^-$ megakaryocytes. There were 424 highly-expressed genes among *ABO*$^+$ megakaryocytes compared with *ABO*$^-$ cells. D) Pathway enrichment analysis of 424 highly-expressed genes based on the KEGG resource. E) Barplot exhibiting the proportion of *ABO*$^+$ megakaryocytes among normal controls, mild, moderate, and severe COVID-19 patients. F) Barplot showing the differentially up-DEGs from the pairwise comparisons of normal controls with different COVID-19 severities. Venn plot on the top of bar showing the overlapped up-DEGs between moderate and severe patients. G) Heatmap showing the up-DEGs from the pairwise comparisons. F) Gene-drug interaction analysis for 35 up-DEGs. I)-N) Representative up-DEGs among *ABO*$^+$ megakaryocytes showing significantly elevated expressions with increased COVID-19 severities.

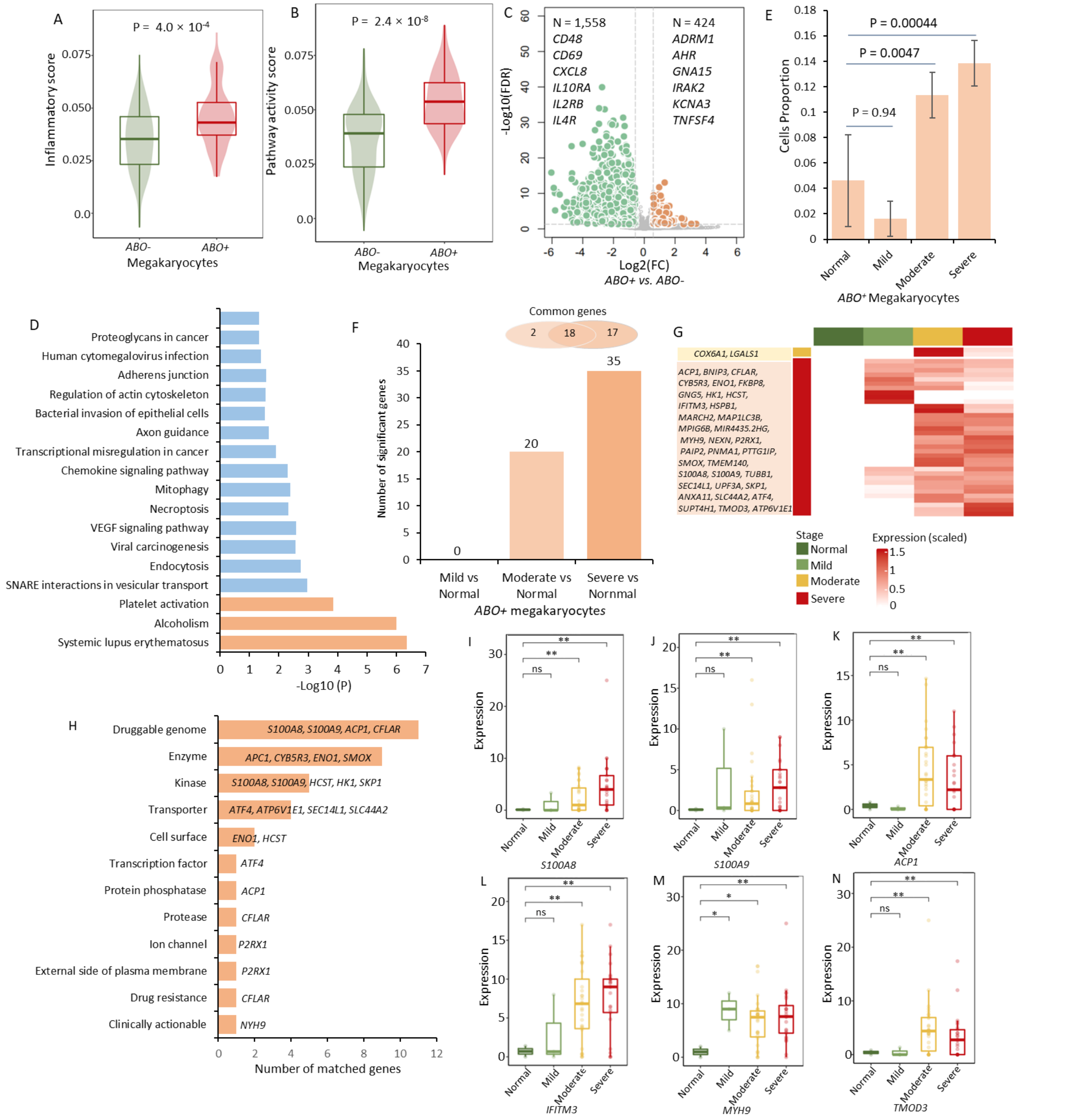

**Supplemental Figure S19. Evidence showing the multi-functionality of *CXCR6*$^+$ memory CD8+T cells for severe COVID-19.** A)-C) Boxplots showing the difference of inflammatory score (A), proliferation score (B), and migration score (C) between *CXCR6*$^+$ and *CXCR6*$^-$ memory CD8+T cells.

Two-side Wilcoxon sum-rank test was used. D) Pathway enrichment analysis of 158 highly-expressed genes based on the KEGG resource. E)-F) Boxplots showing the cytotoxicity score (E) and exhaustion score (F) of among normal, mild, moderate, and severe groups. G)-N) Representative up-DEGs among *CXCR6*[+] memory CD8+T cells showing significantly elevated expressions with increased COVID-19 severities. G) *TCF7*, H) *HELB*, I) *COPS6*, J) *PSMB6*, K) *PUF60*, L) *RAB5IF*, M) *NDUFAF3*, and N) *HLA-DPB1*.

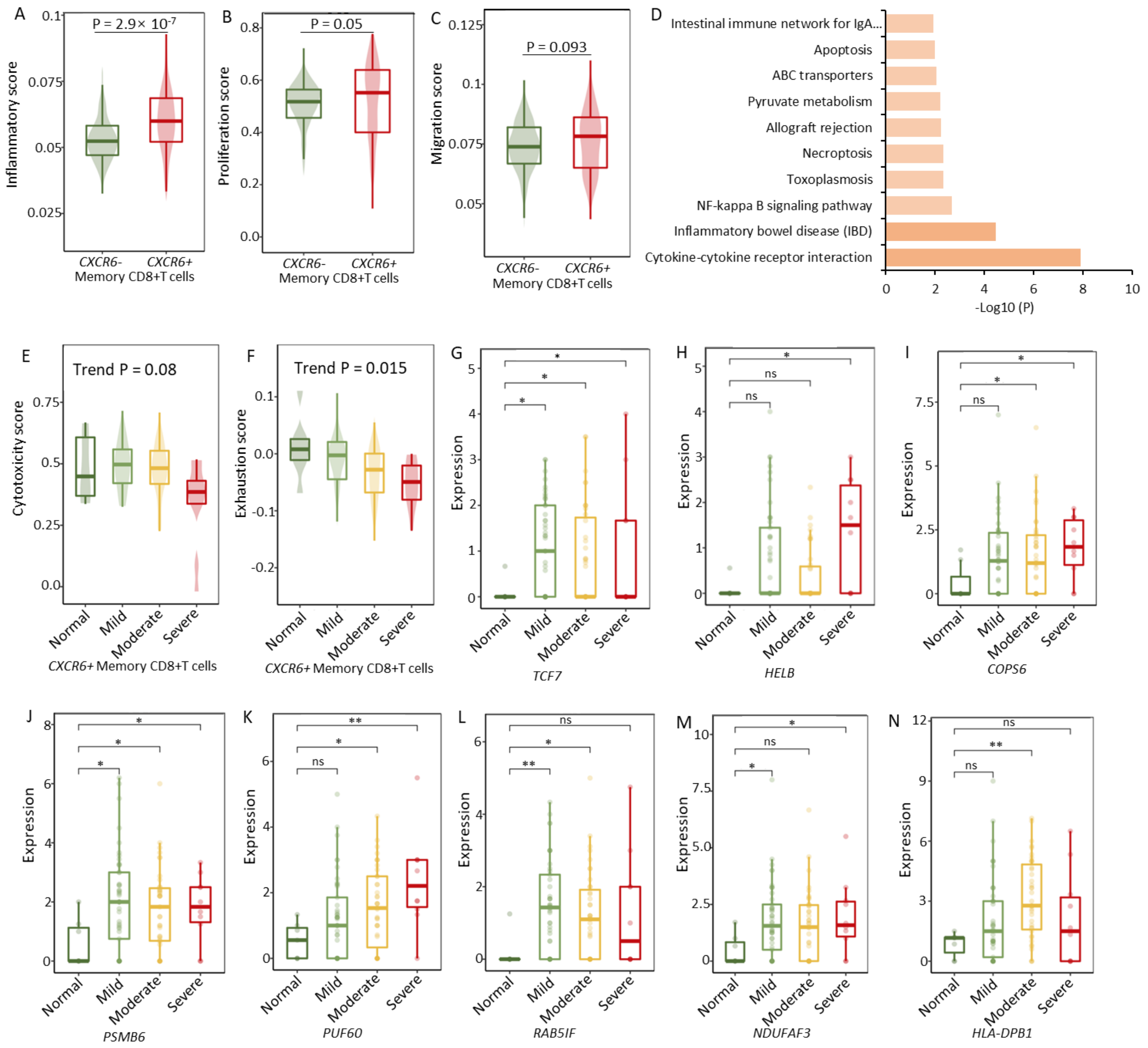

**Supplemental Figure S20. Differences in the number of predicated cell-to-cell interactions in PBMCs, comparing severe COVID-19 patients with normal controls.** Color legend represents the differential number of cellular interactions. Red rectangle marks the elevated interactions of both *CCR1*$^{+}$ CD16+monocytes and *CXCR6*$^{+}$ memory CD8+T cells with other immune cells in PBMCs.

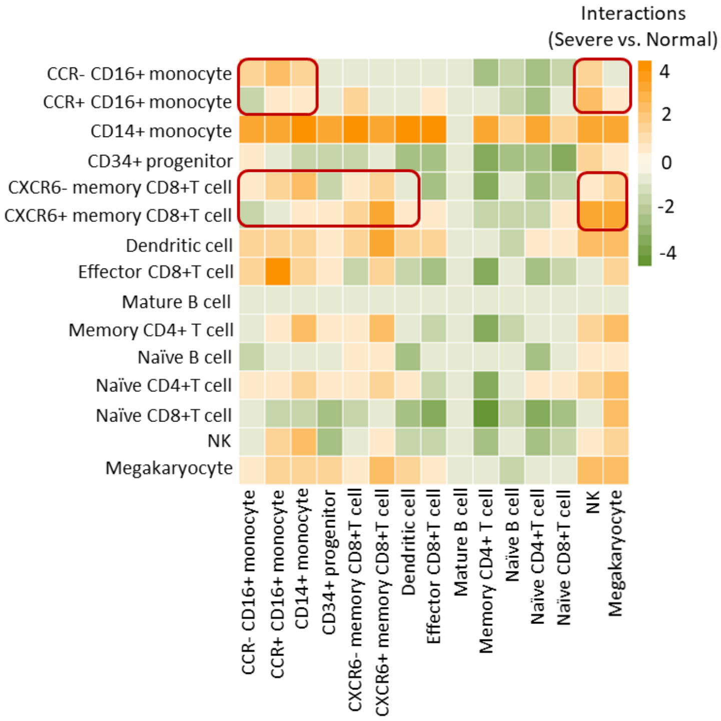

**Supplemental Figure S21. Prediction of cell-to-cell interactions of cells in BALFs.** A) The number of predicated cellular interactions of cells in BALFs. Color legend represents the number of predicted cellular interactions. The Arabic numerals in the heatmap represent the specific number of cellular interactions between pairwise cell types in BALFs. B) The increased percent of cellular interactions of *CCR1*$^{+}$ CD16+monocytes with epithelial cells compared with *CCR1*$^{-}$ CD16+monocytes. C) The increased percent of cellular interactions of *CXCR6*$^{+}$ memory CD8+T cells with epithelial cells compared with *CXCR6*$^{-}$ memory CD8+T cells.

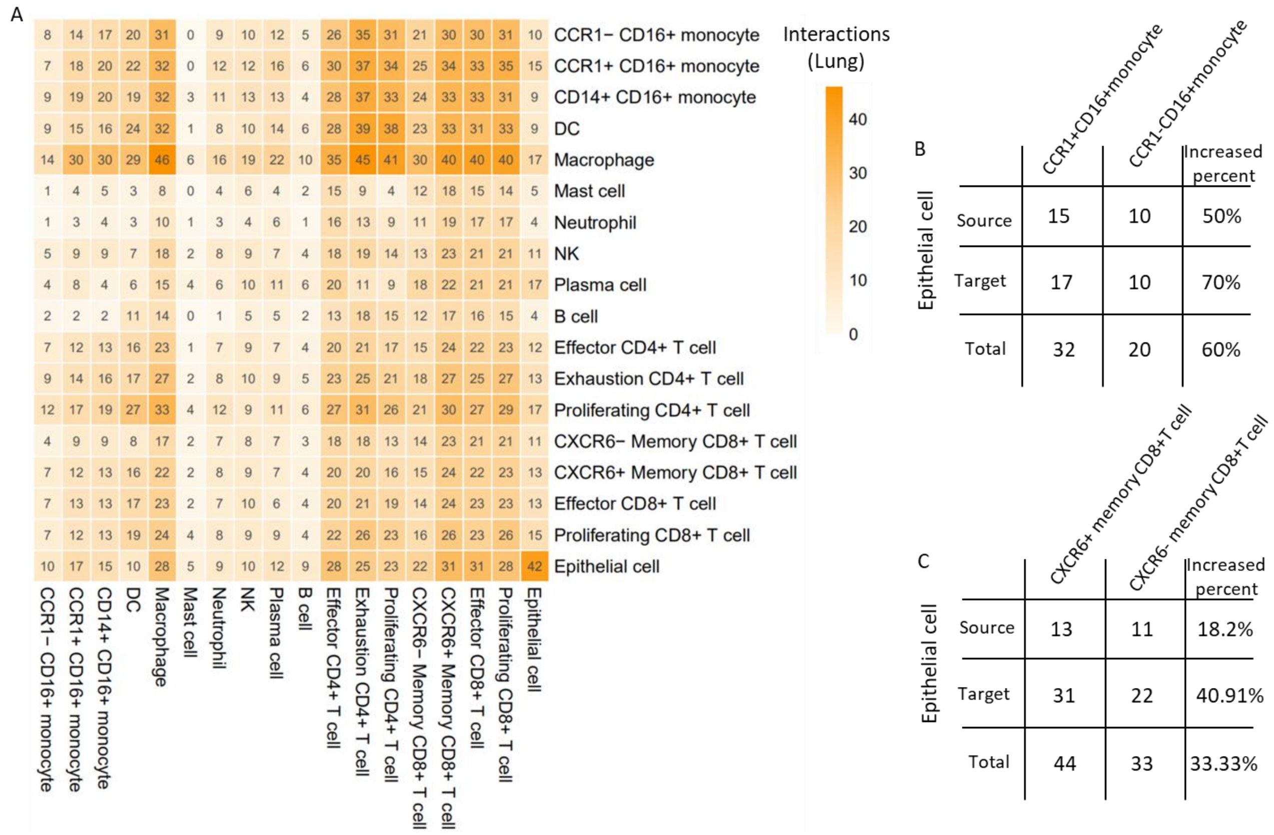

#### 3. *Supplemental Tables*

**Supplemental Table S1. Samples collected from four independent scRNA-seq datasets on COVID-19**

| **Characteristics** | **Dataset #1 (E-MTAB-9357)** | **Dataset #2 (GSE149689)** | **Dataset #3 (GSE150861)** | **Dataset #4 (GSE158055)** | **Total samples** |
|---|---|---|---|---|---|
| Normal controls | 16 | 4 | 0 | 0 | 20 |
| Mild COVID-19 | 96 | 5 | 0 | 0 | 101 |
| Moderate COVID-19 | 106 | 0 | 0 | 3 | 109 |
| Severe COVID-19 | 52 | 6 | 3 | 9 | 70 |
| Remission | 0 | 0 | 4 | 0 | 4 |

**Supplemental Table S2. Selected well-known markers used to define cell types in PBMCs**

| **Cell types** | **Markers** |
|---|---|
| CD14+ monocytes | *CD14, CD4, NR4A1* |
| CD16+ monocytes | *FCGR3A, CX3CR1, CD16, CD4, NR4A1* |
| Na ïve CD4+T cells | *LEF1, CD197, TCF7, CD3D, CD3E, CD4* |
| Na ïve CD8+T cells | *LEF1, CD197, TCF7, CD3D, CD3E, CD8A* |
| Effector CD4+T cells | *PRDM1, PRF1, GZMB, GNLY, CD3D, CD3E, CD4* |
| Effector CD8+T cells | *PRDM1, PRF1, GZMB, GNLY, CD3D, CD3E, CD8A* |
| Memory CD4+T cells | *GZMK, CD69, AQP3, CD3D, CD3E, CD4* |
| Memory CD8+T cells | *GZMK, CD69, AQP3, CD3D, CD3E, CD8A* |
| Exhaustion CD4+T cells | *LAG3, TIGIT, CD279,PDCD1, TIM3, CD3D, CD3E, CD4* |
| Exhaustion CD8+T cells | *LAG3, TIGIT, CD279, PDCD1, TIM3, CD3D, CD3E, CD8A* |
| Proliferating CD4+T cells | *MK167, TYMS, CD3D, CD3E, CD4* |
| Proliferating CD8+T cells | *MK167, TYMS, CD3D, CD3E, CD8A* |
| Regulatory CD4+T cells | *FOXP3, CD3D, CD3E, CD4* |
| Regulatory CD8+T cells | *FOXP3, CD3D, CD3E, CD8A* |
| NK | *FCGR3A, CD16, TRGC1, NKG7,NCAM1,CD38,CD62L,GZMB,CD56* |
| B | *CD79A, MS4A1, CD19* |
| Plasma B | *CD79A, CD38* |
| Na ïve B | *IGHD, FCER2* |
| mDC | *CD14, CD1C* |
| pDC | *CLEC4A, CD123* |
| Megakaryocyte | *PPBP* |
| Progenitor | *CD34, CD38* |
| DC | *FCER1A, IL3RA, CD1C, CD141* |

**Supplemental Table S3. Significant SNPs associated with severe COVID-19 identified by meta-GWAS analysis**

| SNP | CHR | POS | Loci | ATL | OR (95% CI) | Meta P-value |
|---|---|---|---|---|---|---|
| rs35081325 | 3 | 45489921 | 3p21.31 | T | 1.81 (1.788-1.853) | 3.32E-58 |
| rs2109069 | 19 | 4319443 | 19p13.3 | A | 1.183 (1.175-1.191) | 6.40E-13 |
| rs13050728 | 21 | 34215210 | 21q22.11 | C | 0.858 (0.851-0.865) | 1.914E-11 |
| rs143334143 | 6 | 30721426 | 6p21.33 | A | 1.273 (1.241-1.273) | 1.283E-10 |
| rs505922 | 9 | 135749229 | 9q34.2 | T | 0.88 (0.875-0.886) | 2.24E-09 |
| rs622568 | 7 | 54247894 | 7p11.2 | C | 1.167 (1.158-1.175) | 2.57E-08 |
| rs2166172 | 1 | 90808514 | 1p22.2 | C | 1.132 (1.126-1.137) | 2.74E-08 |
| rs2269899 | 12 | 112981956 | 12q24.13 | T | 1.132 (1.127-1.138) | 3.24E-08 |

**Note:** SNP =single nucleotide polymorphism, CHR = chromosome, POS = position, ATL = altered allele, OR = odds ratio, 95% CI = 95% confidence interval, Meta P-value is generated from the Metal tool by meta-analyzing 969,689 samples from 21 independent contributing studies.

**Supplemental Table S4. Significant genes associated with severe COVID-19 identified by MAGMA gene-based association analysis**

| Gene name | CHR | Start POS | Stop POS | Number of SNPs | Z score | P value | FDR |
|---|---|---|---|---|---|---|---|
| *LZTFL1* | 3 | 45844808 | 45977216 | 314 | 13.14 | 1.01E-39 | 1.93E-35 |
| *FYCO1* | 3 | 45939391 | 46057316 | 331 | 11.15 | 3.67E-29 | 3.51E-25 |
| *XCR1* | 3 | 46042291 | 46088979 | 165 | 10.28 | 4.36E-25 | 2.78E-21 |
| *CXCR6* | 3 | 45964973 | 46009845 | 120 | 9.91 | 1.86E-23 | 8.90E-20 |
| *CCR9* | 3 | 45907996 | 45964667 | 152 | 9.19 | 2.01E-20 | 7.69E-17 |
| *SLC6A20* | 3 | 45776941 | 45858039 | 208 | 7.33 | 1.19E-13 | 3.80E-10 |
| *CCR3* | 3 | 46263872 | 46328197 | 214 | 7.19 | 3.35E-13 | 9.16E-10 |
| *CCR1* | 3 | 46223200 | 46269832 | 177 | 7.00 | 1.30E-12 | 3.11E-09 |
| *IFNAR2* | 21 | 34582231 | 34656831 | 221 | 6.83 | 4.21E-12 | 8.95E-09 |
| *CCR2* | 3 | 46375235 | 46422413 | 136 | 6.53 | 3.26E-11 | 6.24E-08 |
| *CCR5* | 3 | 46391633 | 46437697 | 133 | 5.69 | 6.28E-09 | 1.01E-05 |
| *OAS3* | 12 | 113356249 | 113431056 | 265 | 5.69 | 6.35E-09 | 1.01E-05 |
| *OAS1* | 12 | 113324739 | 113377712 | 157 | 5.28 | 6.57E-08 | 9.67E-05 |
| *SPPL2C* | 17 | 43902256 | 43944438 | 281 | 4.63 | 1.81E-06 | 2.47E-03 |

| | | | | | | | |
|---|---|---|---|---|---|---|---|
| *CRHR1* | 17 | 43677710 | 43933194 | 1203 | 4.59 | 2.24E-06 | 2.86E-03 |
| *STH* | 17 | 44056616 | 44097060 | 225 | 4.57 | 2.43E-06 | 2.91E-03 |
| *MAPT* | 17 | 43951702 | 44125700 | 901 | 4.41 | 5.09E-06 | 5.73E-03 |
| *UBE2D1* | 10 | 60074739 | 60150513 | 188 | 4.40 | 5.44E-06 | 5.78E-03 |
| *KANSL1* | 17 | 44087282 | 44322740 | 941 | 4.20 | 1.35E-05 | 1.36E-02 |
| *TFAM* | 10 | 60124903 | 60178990 | 145 | 4.02 | 2.94E-05 | 2.81E-02 |
| *CCRL2* | 3 | 46428721 | 46474488 | 195 | 3.99 | 3.33E-05 | 3.03E-02 |
| *IBA57* | 1 | 228333429 | 228389958 | 118 | 3.98 | 3.52E-05 | 3.06E-02 |
| *IL10RB* | 21 | 34618665 | 34689539 | 230 | 3.92 | 4.36E-05 | 3.63E-02 |
| *COX10* | 17 | 13952719 | 14131996 | 549 | 3.87 | 5.37E-05 | 4.28E-02 |
| *NCR2* | 6 | 41283528 | 41338625 | 328 | 3.83 | 6.32E-05 | 4.84E-02 |

**Note:** SNP =single nucleotide polymorphism, CHR = chromosome, POS = position, FDR = False discovery rate.

**Supplemental Table S5. Significant enriched pathways associated with severe COVID-19 identified from MAGMA-based pathway enrichment analysis**

| ID | KEGG ID | Pathway names | Number of genes | Beta | SE | P value |
|---|---|---|---|---|---|---|
| 1 | hsa00533 | Glycosaminoglycan biosynthesis - keratan sulfate | 14 | 0.66 | 0.20 | 5.84E-04 |
| 2 | hsa05168 | Herpes simplex virus 1 infection | 183 | 0.16 | 0.06 | 2.13E-03 |
| 3 | hsa00601 | Glycosphingolipid biosynthesis - lacto and neolacto series | 27 | 0.43 | 0.15 | 2.69E-03 |
| 4 | hsa04390 | Hippo signaling pathway | 153 | 0.17 | 0.06 | 2.87E-03 |
| 5 | hsa05162 | Measles | 131 | 0.19 | 0.07 | 3.13E-03 |
| 6 | hsa05164 | Influenza A | 167 | 0.15 | 0.06 | 6.36E-03 |
| 7 | hsa04064 | NF-kappa B signaling pathway | 92 | 0.19 | 0.08 | 9.44E-03 |
| 8 | hsa04621 | NOD-like receptor signaling pathway | 167 | 0.13 | 0.06 | 1.11E-02 |
| 9 | hsa04625 | C-type lectin receptor signaling pathway | 104 | 0.16 | 0.07 | 1.13E-02 |
| 10 | hsa05143 | African trypanosomiasis | 34 | 0.28 | 0.12 | 1.33E-02 |
| 11 | hsa05160 | Hepatitis C | 131 | 0.14 | 0.07 | 1.63E-02 |
| 12 | hsa04392 | Hippo signaling pathway - multiple species | 28 | 0.28 | 0.13 | 1.80E-02 |
| 13 | hsa00603 | Glycosphingolipid biosynthesis - globo and isoglobo series | 15 | 0.48 | 0.23 | 1.81E-02 |
| 14 | hsa04137 | Mitophagy - animal | 64 | 0.17 | 0.08 | 1.87E-02 |
| 15 | hsa04630 | JAK-STAT signaling pathway | 158 | 0.12 | 0.07 | 3.50E-02 |
| 16 | hsa04668 | TNF signaling pathway | 110 | 0.13 | 0.07 | 3.95E-02 |

| | | | | | | |
|---|---|---|---|---|---|---|
| 17 | hsa00471 | D-Glutamine and D-glutamate metabolism | 5 | 0.57 | 0.34 | 4.47E-02 |
| 18 | hsa04060 | Cytokine-cytokine receptor interaction | 286 | 0.09 | 0.05 | 4.67E-02 |
| 19 | hsa05132 | Salmonella infection | 81 | 0.14 | 0.09 | 4.92E-02 |

**Supplemental Table S6. The 16 significant genes associated with severe COVID-19identified by S-MultiXcan analysis based on 49 tissues from GTEx consortium**

| Gene name | CHR | START | STOP | P value | FDR |
|---|---|---|---|---|---|
| *LZTFL1* | chr3 | 45864808 | 45957216 | 1.08E-30 | 2.41E-26 |
| *SLC6A20* | chr3 | 45796941 | 45838039 | 2.19E-29 | 2.44E-25 |
| *CCR9* | chr3 | 45927996 | 45944667 | 4.16E-29 | 3.10E-25 |
| *CXCR6* | chr3 | 45984973 | 45989845 | 1.56E-28 | 8.71E-25 |
| *XCR1* | chr3 | 46062291 | 46068979 | 4.20E-16 | 1.88E-12 |
| *FYCO1* | chr3 | 45959391 | 46037316 | 1.53E-14 | 5.69E-11 |
| *CCR3* | chr3 | 46283872 | 46308197 | 8.15E-13 | 2.60E-09 |
| *CCR1* | chr3 | 46243200 | 46249832 | 1.13E-11 | 3.15E-08 |
| *DNAH3* | chr16 | 20944476 | 21170762 | 4.44E-10 | 1.10E-06 |
| *CCR2* | chr3 | 46395235 | 46402413 | 1.16E-09 | 2.59E-06 |
| *ABO* | chr9 | 136130563 | 136150630 | 1.36E-07 | 2.76E-04 |
| *IFNAR2* | chr21 | 34602231 | 34636831 | 3.28E-07 | 6.10E-04 |
| *IL10RB* | chr21 | 34638665 | 34669539 | 8.28E-07 | 1.42E-03 |
| *CCR5* | chr3 | 46411633 | 46417697 | 1.22E-06 | 1.95E-03 |
| *OAS3* | chr12 | 113376249 | 113411056 | 2.83E-06 | 4.21E-03 |
| *CCRL2* | chr3 | 46448721 | 46454488 | 4.61E-06 | 6.43E-03 |

**Note:** CHR = chromosome, START = start position on chromosome, STOP = stop position on chromosome, FDR = False discovery rate.

**Supplemental Table S7. The eight significant genes associated with severe COVID-19 identified by S-PrediXcan analysis based on lung and blood tissues**

| Gene name | CHR | START | STOP | Z score | P value | FDR | Tissue |
|---|---|---|---|---|---|---|---|
| *CCR9* | chr3 | 45927996 | 45944667 | 11.69 | 1.47E-31 | 1.79E-27 | Blood |
| *PGLS* | chr19 | 17622278 | 17632097 | -4.64 | 3.43E-06 | 2.09E-02 | Blood |
| *CXCR6* | chr3 | 45984973 | 45989845 | -10.55 | 5.10E-26 | 7.41E-22 | Lung |

| | | | | | | | |
|---|---|---|---|---|---|---|---|
| *CCR5* | chr3 | 46411633 | 46417697 | -5.46 | 4.85E-08 | 3.52E-04 | Lung |
| *FOXP4* | chr6 | 41514164 | 41570122 | 4.88 | 1.08E-06 | 5.23E-03 | Lung |
| *IL10RB* | chr21 | 34638665 | 34669539 | 4.74 | 2.19E-06 | 7.95E-03 | Lung |
| *FYCO1* | chr3 | 45959391 | 46037316 | 4.65 | 3.35E-06 | 8.45E-03 | Lung |
| *ABO* | chr9 | 136130563 | 136150630 | 4.64 | 3.49E-06 | 8.45E-03 | Lung |

**Note:** CHR = chromosome, START = start position on chromosome, STOP = stop position on chromosome, FDR = False discovery rate.

**Supplemental Table S8. The biological pathways enriched by 34 risk genes associated with severe COVID-19**

| ID | KEGG ID | Pathway names | Number of genes | Ratio | P Value | FDR |
|---|---|---|---|---|---|---|
| 1 | hsa04060 | Cytokine-cytokine receptor interaction | 294 | 10.89 | 3.89E-08 | 1.27E-05 |
| 2 | hsa04062 | Chemokine signaling pathway | 189 | 13.17 | 5.12E-07 | 8.35E-05 |
| 3 | hsa05160 | Hepatitis C | 131 | 10.86 | 4.29E-04 | 3.60E-02 |
| 4 | hsa05162 | Measles | 132 | 10.78 | 4.42E-04 | 3.60E-02 |
| 5 | hsa04621 | NOD-like receptor signaling pathway | 168 | 8.47 | 1.10E-03 | 6.36E-02 |
| 6 | hsa05164 | Influenza A | 171 | 8.32 | 1.17E-03 | 6.36E-02 |
| 7 | hsa05168 | Herpes simplex infection | 185 | 7.69 | 1.57E-03 | 6.51E-02 |
| 8 | hsa05167 | Kaposi sarcoma-associated herpesvirus infection | 186 | 7.65 | 1.60E-03 | 6.51E-02 |
| 9 | hsa05169 | Epstein-Barr virus infection | 201 | 7.08 | 2.13E-03 | 0.077 |
| 10 | hsa05163 | Human cytomegalovirus infection | 225 | 6.32 | 3.20E-03 | 0.104 |

**Supplemental Table S9. The percentage of three severe COVID-19-risk genes expressed in all 13 distinct cell types in PBMCs**

| Cell types | *CXCR6* | *CCR1* | *ABO* |
|---|---|---|---|
| Effector CD8+T cells | 0.007 | 0.008 | 0.002 |

| | | | |
|---|---|---|---|
| Memory CD8+T cells | 0.021 | 0.014 | 0.004 |
| Naive CD8+T cells | 0.013 | 0.01 | 0.003 |
| Naive CD4+T cells | 3.08E-04 | 0.001 | 0.012 |
| Memory CD4+T cells | 0.004 | 0.002 | 0.014 |
| CD14+ monocytes | 8.84E-05 | 0.099 | 0.002 |
| CD16+ monocytes | 2.42E-04 | 0.079 | 0.002 |
| NK | 3.19E-03 | 0.012 | 0.001 |
| Naive B cells | 3.27E-04 | 0.004 | 0.007 |
| Dendritic cells | 1.63E-04 | 0.051 | 0.007 |
| Megakaryocytes | 1.64E-03 | 0.004 | 0.112 |
| CD34+Progenitors | 1.17E-03 | 0.011 | 0.039 |
| Mature B cells | 0 | 0.024 | 0 |

**Supplemental Table S10. Summary of inflammatory and cytokine-related genes and genes in two identified KEGG pathways**

| Categories | Gene lists | Resources |
|---|---|---|
| Inflammatory-related genes | *ABCA1, ABI1, ACVR1B, ACVR2A, ADGRE1, ADM, ADORA2B, ADRM1, AHR, APLNR, AQP9, ATP2A2, ATP2B1, ATP2C1, AXL, BDKRB1, BEST1, BST2, BTG2, C3AR1, C5AR1, CALCRL, CCL17, CCL2, CCL20, CCL22, CCL24, CCL5,CCL7, CCR7, CCRL2, CD14, CD40, CD48, CD55, CD69, CD70,CD82, CDKN1A, CHST2, CLEC5A, CMKLR1, CSF1, CSF3, CSF3R, CX3CL1, CXCL10, CXCL11, CXCL6, CXCL8, CXCL9, CXCR6, CYBB, DCBLD2, EBI3, EDN1, EIF2AK2, EMP3, EREG, F3, FFAR2, FPR1, FZD5, GABBR1, GCH1, GNA15, GNAI3, GP1BA, GPC3, GPR132, GPR183, HAS2, HBEGF, HIF1A, HPN, HRH1, ICAM1, ICAM4, ICOSLG, IFITM1, IFNAR1, IFNGR2, IL10, IL10RA, IL12B, IL15, IL15RA, IL18, IL18R1, IL18RAP, IL1A, IL1B, IL1R1, IL2RB, IL4R, IL6, IL7R, INHBA, IRAK2, IRF1, IRF7, ITGA5, ITGB3, ITGB8, KCNA3, KCNJ2, KCNMB2, KIF1B, KLF6, LAMP3, LCK, LCP2, LDLR, LIF, LPAR1, LTA, LY6E, LYN, MARCO, MEFV, MEP1A, MET, MMP14, MSR1, MXD1, MYC, NAMPT, NDP, NFKB1, NFKBIA, NLRP3, NMI, NMUR1, NOD2, NPFFR2, OLR1, OPRK1, OSM, OSMR, P2RX4, P2RX7, P2RY2, PCDH7, PDE4B, PDPN, PIK3R5, PLAUR, PROK2, PSEN1, PTAFR, PTGER2, PTGER4, PTGIR, PTPRE, PVR, RAF1, RASGRP1, RELA, RGS1, RGS16, RHOG, RIPK2, RNF144B, ROS1, RTP4, SCARF1, SCN1B, SELE, SELENOS, SELL, SEMA4D, SERPINE1, SGMS2, SLAMF1, SLC11A2, SLC1A2, SLC28A2, SLC31A1, SLC31A2, SLC4A4, SLC7A1, SLC7A2, SPHK1, SRI, STAB1, TACR1,* | PMID: 33657410 |

| | | |
|---|---|---|
| | *TACR3, TAPBP, TIMP1, TLR1, TLR2, TLR3, TNFAIP6, TNFRSF1B, TNFRSF9, TNFSF10, TNFSF15, TNFSF9, TPBG, VIP* | |
| Cytokine-related genes | *IL2, IL7, CSF3, CXCL10, CCL2, CCL3, TNF, TFTN1, IL6* | PMID: 32192578 |
| Cytokine-related genes | *CXCL10, CCL7, IL1RN, CSF1, IFNG, IL6, IL2RA, IL10, IL18, HGF, CXCL9, CSF3, CCL3, CCL27* | Exuberant elevation of IP-10, MCP-3 and IL-1ra during SARS-CoV-2 2 infection is associated with disease severity and fatal outcome, 2020.3, *MedRxiv* |
| Cytokine-related genes | *TGFB1* | PMID: 32346099 |
| Cytokine-related genes | *IL1B, LTA, IFNG, CSF1, CSF2, LTB, IL6, TNFSF13, IL18, IL2, IL4* | PMID: 32377375 |
| Cytokine-related genes | *IL1B, IL18, IL6, TNF, CCL2, CCL7, CCL12, CXCL8, CCL3, CXCL9, CXCL10, CXCL11* | PMID: 32505227 |
| Cytokine-related genes | *IL1B, IL6, TNF, CCL2, CCL3, CCL4, CCL7, CXCL9, CXCL10, CXCL11, CXCL1, CXCL2, CXCL3, CXCL8, CCL3L1, CCL8, CXCL16* | PMID: 32398875 |
| Cytokine-related genes | *IL6, CXCL8* | PMID: 32434211 |
| Cytokine-related genes | *CSF2, IL6, IFNG* | Pathogenic T-cells and inflammatory monocytes incite inflammatory storms in severe COVID-19 patients, 2020.6, National Science Review |
| Cytokine-related genes | *IL6, IL10, CXCL8, CXCL10, IFNG, IFNA1* | A consensus Covid-19 immune signature combines immuno-protection with discrete sepsis-like traits associated with poor prognosis., 2020.6, medRxiv |
| Cytokine-related genes | *CXCL10, CXCL9, CCL2, IL1RN, CCL5, CCL11, TNF, HGF, IFNA2* | PMID: 32669297 |
| Cytokine-related genes | *TNF, IL1B, IL18* | PMID: 32651212 |
| Cytokine-related genes | *TNF, CCL3, CCL4, CCL20, IL1B, IL6, IL10, CXCL2, CXCL3, CXCL8, CXCL9, CCL3L1, CCL4L2* | PMID: 32764665 |
| Cytokine-related genes | *IL6, CXCL10, CCL7, HGF, OSM, TNFSF14, IFNA1, SA100A12, FGF19, CXCL5, CCL4, CCL8, CCL19, CXCL11, CCL3, IL18R1, CSF1, TNF, CCL20, TGFA, IFNB1* | PMID: 32788292 |

| | | |
|---|---|---|
| Cytokine-related genes | *TNF* | PMID: 32877699 |
| Cytokine-related genes | *IL6, IL8, TNF, IL1B* | PMID: 32839624 |
| Cytokine-related genes | *IL17C, TNFSF10, FGF7, XCL1, FGF13, LIF, TGFB3, INHBE, CERS1, TXLNA, IFNW1, IL22, XCL2, CCL25, CCL16, CD40LG, IL20, FASLG, TPO, SCYL3, PF4V1, TNFSF8, GDF15, IL1A, VEGFA, GDF7, BMP6, PDGFA, IL21, ABCD-1, ABCD-2, PDGFB, TNFSF4, FAM19A1, HBEGF, PDGFD, IL12RB2, GH1, VEGFB, MIP3B, IL27, PF4, BMP8B, TNFSF12, IL15, SCYL2, SCYL1, TSLP, GDF11, SDF1B, INHBA, PPBP, FGF11, IFNG-AS1, FGF22, VEGFC, CCL18, TNFSF11, IL12A, EBI3, AMH, IL26, IL32, PDGFC, FGF23, IGF1, IL1F11, CCL28, CLCF1, TNFSF9, BMP3, IL24, GDF10, CXCL6, GDF9, IL23A, IL16, CD70, IL5, FGF9, IFNL1, TSC1, FGF2, IL23R, IL1G, SPP1, IL12RB1, BMP4, IL13, TPAR1, TGFB2, FAM19A2, AGIF3, EDA, MIF, TNFSF13B, BMP7, FGF18, CCL23* | KEGG database (PMID: 10592173) |
| Genes in the KEGG pathway of cytokine-cytokine receptor interaction (hsa04060) | *GDF11, CCL26, CXCL13, CXCR6, TNFSF13B, CCR9, CCL27, EDAR, IL24, IL17F, TNFRSF13C, CCR1, CCR3, CCR4, CCR5, CCR6, CCR7, CCR8, CNTF, CNTFR, ACVR1C, IL17RE, IL31RA, CSF1, CSF1R, CSF2, CSF2RB, CSF3, CSF3R, CSH1, CSH2, IL34, CTF1, IL23R, GDF7, CX3CR1, IFNLR1, EDA, EPO, EPOR, TNFRSF13B, CLCF1, IL17RA, IL27, IL36RN, GDF1, GDF2, MSTN, GDF9, GDF10, AMH, GH1, GH2, AMHR2, GHR, IL36B, IL37, IL36A, IL17C, IL17B, TNFRSF21, BMP10, CCR10, IFNL2, IFNL3, IFNL1, XCR1, CXCR3, CXCL17, CXCL1, CXCL2, CXCL3, IL19, IFNE, IFNA1, IFNA2, IFNA4, IFNA5, IFNA6, IFNA7, IFNA8, IFNA10, IFNA13, IFNA14, IFNA16, IFNA17, IFNA21, IFNAR1, IFNAR2, IFNB1, IFNG, IFNGR1, IFNGR2, IFNW1, BMP8A, FAS, IL1A, IL1B, IL1R1, IL1RAP, IL1RN, IL2, IL2RA, FASLG, IL2RB, IL2RG, IL3, IL4, IL4R, IL5, IL5RA, IL6, IL6R, IL6ST, IL7, IL7R, CXCL8, CXCR1, IL9, CXCR2, IL10, IL10RA, IL10RB, IL11, IL11RA, IL12A, IL12B, IL12RB1, IL12RB2, IL13, IL13RA1, IL13RA2, IL15, IL15RA, IL16, TNFRSF9, IL17A, IL18, INHA, INHBA, INHBB, INHBC, CXCL10, IL31, CCL4L1, GDF6, LEP, LEPR, LIF, LIFR, LTA, LTB, LTBR, CCL3L3, CXCL9, MPL, NGF, NGFR, NODAL, TNFRSF11B, OSM, IL20, IL21R, IL22, TNFRSF12A, ACKR4, IL23A, PF4, PF4V1, IL17D, IL20RA, IL20RB, PPBP, TNFRSF19, IL17RB, IL26, PRL, PRLR, IL36G, CCL28, IFNK, ACKR3, CXCL16, IL22RA1, IL21, EDA2R, TNFRSF17, CCL1, CCL2, CCL3, CCL3L1, CCL4, CCL5, CCL7, CCL8, CCL11, CCL13, CCL14, CCL15, CCL16, CCL17, CCL18, CCL19, CCL20, CCL21, CCL22, CCL23, CCL24, CCL25, CXCL6, CXCL11, CXCL5, XCL1, CX3CL1, CXCL12, CXCR5, IL25, BMP2, BMP3, BMP4, BMP5, BMP6, BMP7, BMP8B, BMPR1A, BMPR1B, BMPR2, XCL2, TGFB1, TGFB2, TGFB3, TGFBR1, TGFBR2, THPO, TNF, TNFRSF1A, TNFRSF1B, TNFSF4, CCR2, TNFRSF4, IL1R2, CXCR4, GDF5, INHBE, IL1F10, IL17RC, RELT, TSLP, TNFSF11, TNFRSF25, TNFSF14, TNFSF13, TNFSF12, TNFSF10, TNFSF9, TNFRSF14, TNFRSF6B, TNFRSF18, TNFRSF11A, TNFRSF10D, TNFRSF10C, TNFRSF10B, TNFRSF10A, IL18RAP, IL1RL2, IL18R1, TNFSF18, ACVR1, IL33, ACVR1B, IL1RL1, OSMR, ACVR2A, CD4, BMP15, IL32, ACVR2B, CD27, ACVRL1, TNFRSF8, TNFSF8, IL27RA, GDF15, CXCL14, CCL4L2, GDF3, CD40, CD40LG, CD70, TNFSF15* | KEGG database (PMID: 10592173) |
| Genes in the KEGG pathway of chemokine signaling pathway (hsa04062) | *AKT3, RASGRP2, CCL26, VAV3, CXCL13, CXCR6, GNB5, ADCY1, ADCY2, CCR9, CCL27, ADCY3, ADCY5, ADCY6, ADCY7, ADCY8, CHUK, ADCY9, CCR1, CCR3, CCR4, CCR5, CCR6, CCR7, CCR8, GRK7, CRK, CRKL, PIK3R6, CX3CR1, ADRBK1, ADRBK2, DOCK2, ADCY4, AKT1, AKT2, PTK2B, FGR, FOXO3, PLCB1, PIK3R5, SHC2, GNAI1, GNAI2, GNAI3, GNB1, GNB2, GNB3, GNG3, GNG4, GNG5, GNG7, GNG10, GNG11, GNGT1, GNGT2, CCR10, XCR1, CXCR3, GRK4, GRK5, GRK6, GRB2, CXCL1, CXCL2, CXCL3, GSK3A, GSK3B, HCK, HRAS, IKBKB, CXCL8, CXCR1, CXCR2, CXCL10, ITK, JAK2, JAK3, KRAS, RHOA, CCL4L1, SHC4, LYN, ARRB1, ARRB2, CCL3L3, CXCL9, NFKB1, NFKBIA, NFKBIB, NRAS, PAK1, GNG13, PF4, PF4V1, PIK3CA, PIK3CB, PIK3CD, PIK3CG, PIK3R1, PIK3R2, PLCB2, PLCB3, PLCB4, SHC3, GNG2, PPBP, PRKACA, PRKACB, PRKACG, PRKCB, PRKCD, PRKCZ, MAPK1, MAPK3, GNG12, MAP2K1, PARD3, CCL28, BAD, PTK2, PREX1, CXCL16, PXN, RAC1, RAC2, RAF1, RAP1A, RAP1B, GNB4, RELA, ROCK1, CCL1, CCL2, CCL3, CCL3L1, CCL4, CCL5, CCL7, CCL8, CCL11, CCL13, CCL14, CCL15, CCL16, CCL17, CCL18, CCL19, CCL20, CCL21, CCL22, CCL23, CCL24, CCL25, CXCL6, CXCL11, CXCL5, XCL1, CX3CL1, CXCL12, CXCR5, SHC1, NCF1, SOS1, SOS2, SRC, BRAF, STAT1, STAT2, STAT3, STAT5B, XCL2, TIAM1, CCR2, VAV1, VAV2, WAS, CXCR4, PIK3R3, IKBKG, WASL, GNG8, ROCK2, CXCL14, CCL4L2, BCAR1, ELMO1, CDC42* | KEGG database (PMID: 10592173) |

#### Supplemental Table S11. Highly-expressed inflammatory and cytokine genes among *CCR1*+ CD16+monocytes

| Gene | T score | Fold change | P value | FDR |
|---|---|---|---|---|
| *ADM* | 6.26 | 1.72 | 5.17E-10 | 2.97E-08 |
| *AHR* | 5.16 | 1.71 | 2.83E-07 | 1.07E-05 |
| *AQP9* | 5.70 | 1.96 | 1.47E-08 | 7.07E-07 |
| *C3AR1* | 6.30 | 1.56 | 3.99E-10 | 2.31E-08 |
| *CCL3* | 6.16 | 1.55 | 9.43E-10 | 5.26E-08 |
| *CCL3L1* | 5.57 | 1.71 | 3.01E-08 | 1.37E-06 |
| *CCL4L2* | 3.06 | 1.92 | 2.28E-03 | 2.49E-02 |
| *CD14* | 18.55 | 2.14 | 2.02E-68 | 1.57E-64 |
| *CD82* | 3.98 | 1.59 | 7.16E-05 | 1.43E-03 |
| *CSF3R* | 7.18 | 1.56 | 1.16E-12 | 9.94E-11 |
| *CXCL10* | 4.17 | 3.36 | 3.29E-05 | 7.14E-04 |
| *CXCL2* | 7.12 | 2.72 | 1.83E-12 | 1.53E-10 |
| *CXCL3* | 3.33 | 3.33 | 9.03E-04 | 1.19E-02 |
| *CXCL8* | 5.56 | 1.52 | 3.29E-08 | 1.48E-06 |
| *EIF2AK2* | 7.87 | 1.95 | 7.19E-15 | 7.88E-13 |
| *EREG* | 6.05 | 2.02 | 1.92E-09 | 1.02E-07 |
| *FFAR2* | 5.56 | 1.92 | 3.29E-08 | 1.48E-06 |
| *FPR1* | 10.14 | 1.53 | 2.60E-23 | 5.69E-21 |
| *GPR183* | 2.99 | 1.69 | 2.85E-03 | 2.99E-02 |
| *HBEGF* | 4.55 | 1.70 | 5.84E-06 | 1.57E-04 |
| *IL1B* | 6.66 | 1.58 | 3.89E-11 | 2.65E-09 |
| *IL1RN* | 8.87 | 2.68 | 2.54E-18 | 3.63E-16 |
| *IL27* | 2.90 | 2.73 | 3.75E-03 | 3.67E-02 |
| *IL4R* | 3.29 | 1.58 | 1.04E-03 | 1.33E-02 |
| *ITGA5* | 2.85 | 1.53 | 4.39E-03 | 4.17E-02 |
| *LDLR* | 4.06 | 1.72 | 5.11E-05 | 1.05E-03 |
| *MARCO* | 7.60 | 1.66 | 5.50E-14 | 5.44E-12 |

| | | | | |
|---|---|---|---|---|
| *MYC* | 3.12 | 1.90 | 1.83E-03 | 2.11E-02 |
| *NLRP3* | 4.92 | 1.61 | 9.51E-07 | 3.16E-05 |
| *OSM* | 2.92 | 1.82 | 3.53E-03 | 3.51E-02 |
| *PTAFR* | 6.99 | 1.85 | 4.44E-12 | 3.42E-10 |
| *RTP4* | 3.35 | 1.80 | 8.32E-04 | 1.11E-02 |
| *SELL* | 9.16 | 1.94 | 1.92E-19 | 3.18E-17 |
| *SPHK1* | 4.81 | 2.25 | 1.71E-06 | 5.32E-05 |
| *STAB1* | 7.78 | 2.40 | 1.56E-14 | 1.65E-12 |
| *TNFAIP6* | 3.44 | 3.33 | 5.94E-04 | 8.43E-03 |
| *CCL2* | 3.36 | 5.74 | 8.08E-04 | 1.08E-02 |

**Supplemental Table S12. Pathway enrichment analysis of 351 highly-expressed genes among *CCR1*+ CD16+monocytes.**

| Pathway name | Gene size | Enrichment ratio | P Value | FDR |
|---|---|---|---|---|
| NOD-like receptor signaling pathway | 168 | 4.25 | 4.32E-07 | 9.07E-05 |
| Influenza A | 171 | 4.17 | 5.56E-07 | 9.07E-05 |
| Cytokine-cytokine receptor interaction | 294 | 3.14 | 1.61E-06 | 1.75E-04 |
| Hematopoietic cell lineage | 97 | 5.19 | 2.83E-06 | 2.31E-04 |
| Pertussis | 76 | 5.52 | 1.12E-05 | 7.32E-04 |
| Salmonella infection | 86 | 4.88 | 3.39E-05 | 1.84E-03 |
| Legionellosis | 55 | 6.10 | 4.16E-05 | 1.94E-03 |
| Staphylococcus aureus infection | 56 | 5.99 | 4.75E-05 | 1.94E-03 |
| Complement and coagulation cascades | 79 | 4.78 | 9.93E-05 | 3.60E-03 |
| Toll-like receptor signaling pathway | 104 | 4.03 | 1.73E-04 | 5.63E-03 |
| TNF signaling pathway | 110 | 3.81 | 2.74E-04 | 8.13E-03 |
| NF-kappa B signaling pathway | 95 | 3.98 | 4.09E-04 | 1.11E-02 |

| | | | | |
|---|---|---|---|---|
| Malaria | 49 | 5.14 | 1.01E-03 | 2.52E-02 |
| Hepatitis C | 131 | 3.20 | 1.10E-03 | 2.56E-02 |
| Herpes simplex infection | 185 | 2.72 | 1.50E-03 | 3.26E-02 |
| Chemokine signaling pathway | 189 | 2.66 | 1.80E-03 | 3.67E-02 |
| Amoebiasis | 96 | 3.50 | 1.98E-03 | 3.79E-02 |
| Human cytomegalovirus infection | 225 | 2.42 | 2.71E-03 | 4.91E-02 |
| Chagas disease (American trypanosomiasis) | 102 | 3.29 | 2.90E-03 | 4.97E-02 |
| Phagosome | 152 | 2.76 | 3.32E-03 | 5.42E-02 |
| Cytosolic DNA-sensing pathway | 63 | 4.00 | 3.71E-03 | 5.77E-02 |
| Measles | 132 | 2.86 | 4.16E-03 | 6.17E-02 |
| Transcriptional misregulation in cancer | 186 | 2.48 | 4.78E-03 | 6.77E-02 |
| IL-17 signaling pathway | 93 | 3.16 | 6.57E-03 | 8.92E-02 |
| Glycosaminoglycan degradation | 19 | 6.63 | 9.74E-03 | 1.27E-01 |
| JAK-STAT signaling pathway | 162 | 2.33 | 1.51E-02 | 1.90E-01 |
| Bladder cancer | 41 | 4.09 | 1.59E-02 | 1.92E-01 |
| Acute myeloid leukemia | 66 | 3.18 | 2.01E-02 | 2.19E-01 |
| Inflammatory bowel disease (IBD) | 65 | 3.23 | 1.90E-02 | 2.19E-01 |
| Rheumatoid arthritis | 90 | 2.80 | 2.01E-02 | 2.19E-01 |
| RIG-I-like receptor signaling pathway | 70 | 3.00 | 2.53E-02 | 2.66E-01 |
| Tuberculosis | 179 | 2.11 | 2.70E-02 | 2.75E-01 |
| Nicotinate and nicotinamide metabolism | 30 | 4.20 | 3.37E-02 | 2.97E-01 |
| PPAR signaling pathway | 74 | 2.84 | 3.12E-02 | 2.97E-01 |
| Osteoclast differentiation | 128 | 2.29 | 3.29E-02 | 2.97E-01 |
| Leishmaniasis | 74 | 2.84 | 3.12E-02 | 2.97E-01 |
| Kaposi sarcoma-associated herpesvirus infection | 186 | 2.03 | 3.34E-02 | 2.97E-01 |

| | | | | |
|---|---|---|---|---|
| Antifolate resistance | 31 | 4.06 | 3.67E-02 | 3.15E-01 |
| Th17 cell differentiation | 107 | 2.35 | 4.22E-02 | 3.52E-01 |
| Glycosphingolipid biosynthesis | 15 | 5.59 | 4.84E-02 | 3.85E-01 |

**Supplemental Table S13. Druggble proteins collected from the ChEMBL database**

| Categories | Protein lists | Resources |
|---|---|---|
| 1,263 human druggable proteins | *HGNC_SYMBOL, HTR3E, PSMD12, PLK4, PIK3R2, PSMD14, MSTN, PSMA7, XPO1, DCLK1, PLA2G10, PDPK1, CYP26A1, MLNR, MGAM, HCRTR2, CACNA1F, BRD4, NR1I2, GMNN, STK16, TRPA1, DGAT1, PAK3, HTR3B, BMP10, ABCA1, ABHD16A, F8, F9, TNF, IFNA1, IFNG, IGHE, FN1, TTR, CXCL10, CSF2, MAS1, CD74, CD3D, SHBG, TRAV29DV5, IGF1, CYP17A1, IL5, SERPINE1, CYP1A2, IL6, LCK, CDK1, CD2, CEACAM5, CALCA, LIPF, TUBB, CHRM4, SERPINA6, MMP2, PTPRC, MYL3, HCK, CYP3A4, CYP21A2, HTR1A, CD3G, ALOX5, WEE2, CD28, RARB, THRA, MYL2, HSPA8, CHRM1, PIM1, TOP1, TOP2A, DMD, CDK4, IMPDH2, FCGR1A, SRC, MYH6, RARG, ADRB3, FOLR2, SI, INSRR, PKM, TYR, GABRA1, IFNGR1, FOLR1, CD19, ELN, CYP11B1, PNLIP, ENG, FLT1, ADRA2B, GABRB1, GABRG2, CYP11B2, RXRA, ERBB3, GART, CDH3, CA4, MMP8, PPIB, EDNRB, HRH2, BRD2, PSMA1, PSMA3, HSD3B2, CD27, PIK3R1, PSMB8, PSMB9, PSMB4, PSMB6, RXRB, HSD11B1, CTGF, CD40LG, AVPR2, CALCR, SRD5A2, SSTR3, CNR2, TRHR, PTGS2, OPRM1, SCN4A, PDE6B, IFNGR2, THPO, MPL, ADH7, CASR, GRM5, MC3R, AKR1C3, PIK3CA, CA7, NR4A2, KIR2DL1, KIR2DL2, KIR2DL3, PSMC4, TNNT2, MMP13, NOTCH1, AVPR1B, MTNR1A, CSNK1A1, PIK3CG, MAPKAPK2, PSMB3, CLK1, CLK2, GSK3A, AGTR2, CDK7, IRAK1, PSMD7, CCR4, CCR9, BMX, KIF11, EPHB3, LOXL3, PSMC1, RBX1, TNNC1, TUBA4A, TUBB4B, HBB, HBA1, HBA2, PIP4K2B, HAMP, MUC5AC, CDK6, CDK5, CDK17, NFKB2, SCN7A, FKBP4, KMT2A, RELA, NOTCH2, ACVR1, GRIN1, BTK, LTB, PDE4B, PDE4D, MAP4K2, SLC10A2, GRIN2B, PDE3B, FKBP5, PRPF4B, HTR4, NCOA4, IKBKE, IL13RA2, MELK, NR1I3, EBP, SHH, MYLK, KCNJ8, ADRM1, NFE2L2, RPE65, DDB1, BCL2A1, IL17A, CYP2A13, AOC3, MGAM2, HLA-DRB1, MYLK3, SLC22A6, SLC44A4, TMEM97, TUBA1A, MARK2, SV2A, KCNK18, TPH2, PSMA8, SLC22A8, MAPK15, HDAC2, BCL2L2-PABPN1, GHSR, MAP4K1, APH1A, AURKB, SLC46A1, PVRL4, TAAR1, HBEGF, HSD17B10, TSSK1B, HDAC8, FGF23, IL22, HIPK3, CYP3A43, CD248, PDE7B, TNFRSF12A, KCNK9, DLL4, PSENEN, PIM2, HDAC6, GABBR1, GDF2, CA14, NOTCH3, GABRQ, SRPK3, AKT3, HCN4, ANGPTL3, HRH3, SCN10A, ICOS, TNFSF11, TNFSF14, LTA, CD4, MMP3, IL8, PDGFRA, TNFRSF9, DDR1, GLA, NCAM1, HGF, IL6ST, AKR1C1, MPO, IL12B, IL5RA, PSMB11, MYH7B, PSMD11, BMPR1B, CDC7, PIK3CD, STK25, FAAH, CACNA1A, GABRP, KCNK3, RIOK3, TERT, CHEK1, GABRD, FFAR1, IKBKB, AURKA, GAK, NPC1, ANGPT2, HDAC3, GRIN2D, TLR3, KCNN4, PRPF4, GPR39, PSMD3, MAP3K13, DAPK3, MAP3K7, RIPK2, CACNA1G, KCNQ3, KCNQ2, CA12, HCRTR1, PSCA, PDE6D, AKR1B10, NUAK1, GRIN3B, CCNT1, PDE8A, JAK2, ABCC9, RAMP1, CTSV, FZD7, ROCK2, GUCY1B2, NCOR1, ULK1, CA11, RPS6KA5, RPS6KA4, IDH1, TNFSF13, GABBR2, PDE5A, STK10, PRKD3, KCNK2, S1PR2, CACNA1H, PDE8B, TNKS, ESRRB, MAP4K4, GLP2R, S1PR4, PAK4, CHEK2, ADH1B, ADH1C, ALDH1A1, DHFR, GSR, PAH, SOD1, PNP, ABL1, CFD, F12, PLAT, CA1, CA2, SERPINC1, C5, PDGFB, NGF, IGF2, IL1A, IL1B, TRAC, LMNA, TNNC2, FGB, CHRNA1, AFP, GC, ESR1, KLKB1, TRGV3, HMGCR, RAF1, GBA, APOB, NR3C1, TK1, VWF, TUBB4A, TRBV7-9, ERBB2, NTRK1, TP53, AMY2A, CYP1A1, TYMS, ATP1A1, ATP1B1, APP, ALDH2, ITGB3, ITGB2, CYP11A1, IL4, PRKCG, FGF1, ITGB1, PRKCB, INSR, FYN, BCHE, PGR, EIF4E, NPM1, ITGAV, TH, TPO, KLK3, H1F0, ADH1A, FES, CSF1R, CHRNG, ADRB2, CD3E, HSP90AA1, YES1, LYN, IGF1R, CHRM2, ABCB1, NR3C2, HSP90AB1, ADH4, MME, INHBA, ITGA2B, MET, ADRB1, FCGR3A, ITGA5, VIM, STS, CHRM5, ADRA2A, ROS1, FGF2, HMGB1, TACSTD2, FGR, PARP1, LTA4H, ADORA3, CALM1, TUBA3C, CALCB, GAA, AR, RARA, ARAF, BCL2, TGFB3, CYP2D6, MAPT, KIT, THRB, CD37, UMPS, ITGAM, CHRNB1, BCR, FGFR1, VDR, ESRRA, CYP2A6, CYP19A1, CETP, CYP2C9, ADH5, MS4A1, ODC1, ACE, MYL4, MYH7, CCL2, CFTR, ITGA4,* | PMID: 33837377 |

| | |
|---|---|
| | *ATP1A3, EEF2, SLC5A1, HSD17B1, FDPS, DRD2, IL2RB, BRAF, GLUL, IL9, CSF2RA, PHKG2, MUC1, ALOX15, NPR1, CD44, CTLA4, EPCAM, DPEP1, TSHR, PDE6A, SELE, FER, PRKCA, ITGA2, PRKACA, TYRP1, TPH1, CHRNB2, CR1, PSMC3, ALOX12, ITGB5, SRD5A1, PDE6G, ITGB6, ADRA2C, SDC1, EPOR, TNNI3, AOC1, NFKB1, TYMP, CD22, ALOX5AP, CHRM3, PSMB1, ATP4A, ITGAL, DDC, CYP3A5, IMPDH1, MAG, MAOA, TACR2, S1PR1, CNR1, DRD1, C5AR1, TBXA2R, FGFR2, RYR1, DRD4, DRD5, COMT, IL10, ACHE, FGFR4, FGFR3, PRKACG, PRKACB, LHCGR, PTGS1, GLRA1, RPS6KB1, JAK1, TNFSF4, CPT2, RRM1, FSHR, CMA1, SLC6A2, GABRR1, CCND1, CYP3A7, TBXAS1, PRKCH, CCNE1, CDK2, CXCR1, CXCR2, GUCY2C, ADRA1D, EDNRA, TACR1, PTAFR, PSMA2, PSMA4, ITGB7, DNMT1, IL3RA, MAOB, MAPK3, MARK3, DPP4, PDE4A, PSMA5, PSMB5, HTR1D, HTR1B, HTR2A, ADH6, HTR2C, GABRB3, MAPK1, TNFRSF8, IMPA1, ADORA2A, ADORA2B, EPHA2, EPHA3, EPHA8, TACR3, LTK, CASP1, TYK2, CD6, CCND2, CCND3, WEE1, BDKRB2, SLC6A1, CHRNA5, TSPO, ADORA1, AGTR1, OXTR, SSTR1, SSTR2, CHRNB4, HTR1F, GNRHR, NTSR1, CPS1, RRM2, CD52, SSTR4, SLC5A2, GABRA5, SLC6A4, AKT1, AKT2, ATIC, CCKAR, CCKBR, MC4R, CCR1, CHRNA3, GRK4, HPD, ICAM3, CD70, MC5R, KIF5B, CYP2C19, GUCY1A2, ABCC1, CD80, TTK, GABRA3, EPHX2, HTR7, PTGER1, CRHR1, IL13, SSTR5, ADRA1A, HRH1, ADRA1B, PTGER4, DRD3, SCN1A, SOAT1, BSG, CHKA, PSMC2, MAP2K2, CHRNA7, FLT3, TGFBR1, SCNN1A, PPARG, FDFT1, AVPR1A, GGCX, GRIK1, PLA2G5, CD79B, PSMB10, IL15, PMEL, OPRD1, OPRK1, OPRL1, P2RY2, HNF4A, CSK, IARS, SLC19A1, HTR2B, CCR2, PRKCI, CD86, GRIA1, GRIA2, GRIA3, PIK3CB, MTOR, TEC, TXK, ABL2, FRK, HTT, PTGFR, PTGER3, PTGER2, PTGIR, MCAM, GLP1R, CTSK, GPD2, SYK, TNFRSF4, NAMPT, CHRNA4, TNNT3, MAPK8, MAPK9, RECQL, HTR3A, BDKRB1, GABRA2, GABRB2, GCGR, SCTR, HTR5A, XDH, GRIA4, SLC6A9, GABRA4, RXRG, IFNAR2, PSMD8, CSNK1D, IDH2, SLC9A3, TNNI2, MTNR1B, CDK8, FNTA, FNTB, CSNK1E, PSMB2, PGF, VEGFB, PSEN1, PSEN2, GSK3B, DIO1, NPY4R, HTR6, CDK9, ATP1A2, RAB9A, PDE6C, ATP4B, SCNN1B, SCNN1G, RORC, BLK, CYP2J2, ALDH5A1, CCL11, CCR5, KCNQ1, CLCN2, RPS6KA3, JAK3, PLK1, DAPK1, COL4A4, LIMK2, MAPK12, MAPK10, BLM, CACNA2D1, PRKAA2, ATP1B3, FXYD2, EPHA5, EPHB1, EPHA4, SLC12A3, PSMD4, NR1H2, MTTP, CASP9, GFER, P2RX3, HDAC4, KCNQ4, BACE1, CLDN18, SIK1, KCNK10, SHFM1, PSMA6, CXCR4, TGFB2, PSMC5, PSMC6, ESRRG, PPIA, FKBP1A, KCNJ2, FKBP1B, TUBA1B, CSNK2A1, GABRE, SRPK2, PRKDC, ADAM17, IL13RA1, MAP3K9, HSD11B2, ABAT, CDK16, CACNA1B, MDM2, MYL7, PMP22, MYT1, CACNA1D, MC2R, MC1R, SLC6A3, GUCY1A3, DHODH, GUCY1B3, PRKCE, RHD, GHRHR, MAP2K1, MAP3K10, TOP2B, PPARD, PTH1R, FOLH1, PRKCQ, CHRNE, MST1R, PTK2, PRKCZ, PRKCD, CHRNB3, SLC18A2, PPAT, AOX1, TYRO3, CHRND, BCL2L1, MCL1, PPARA, TNK2, CYP24A1, PPP3CA, PDE4C, PPID, ITK, DMPK, ABCC8, KCNMA1, KCNH2, MAP3K12, GRIN2A, DPYD, FAP, GRIK2, GRIK3, STK4, PLA2G7, PRKAA1, NR1H3, MAP2K5, PAK2, STK3, PSMD2, MAP3K1, PRKG2, PTGDR, GPR17, MSLN, IKZF1, ROCK1, FZD5, TNK1, MADCAM1, BIRC3, BIRC2, TUBB3, ATR, HDAC1, CAMK2G, CAMK2D, NAE1, CUL4A, SLC12A1, DYRK1A, TPBG, CACNA1S, ACVR2B, ATP1A4, BMPR2, PTK6, TUBB2A, CACNA1C, PDE7A, PDE6H, PRKG1, IL18, KEAP1, PTK2B, FZD2, GRM2, PDE3A, SCN5A, SQLE, KCNJ11, GRM3, GRIN2C, PSMD6, BRD3, PDCD1, PDK1, PDK2, PDK3, PRKD1, ERBB4, RPS6KA2, DHCR24, SF3B3, RYR3, RPS6KA1, TESK1, LTB4R, MAPK11, NPY5R, CHRNA2, CHRNA6, STK11, GRK1, SCN9A, GRIK4, CCDC6, GABRA6, GRIK5, PKN1, PKN2, MAPK14, MAP3K11, CALCRL, NTRK2, SMN2, PDK4, HIF1A, UGCG, CA9, PHKG1, DDR2, CYP51A1, TXNRD1, AAK1, TUBB8, PDCD4, MAP3K19, BRDT, TNNI3K, LRRK2, ANO1, CD276, TUBA3E, ULK3, NPSR1, HTR3D, TRPM8, DPP9, PIM3, MYLK4, HIPK1, IFNLR1, CAMK1D, MAP4K3, MAPKAPK5, ULK2, GABRG1, CA13, ABHD12, MINK1, DAGLB, HIPK4, TRPV1, TRPV3, UBA3, GRIN3A, STK35, HCAR2, GPBAR1, GRK7, CLEC4C, HDAC7, APH1B, HTR3C, RXFP2, MUC16, PIK3R5, CPT1B, NCSTN, PIK3R3, ESR2, RORB, BCL2L2, SLCO2A1, BHMT, TSLP, TOP1MT, HDAC10, HDAC11, LINGO1, SLC47A1, LOXL4, EGLN2, NR1H4, CAMKK2, SLC22A12, SRPK1, CRBN, CSF3R, SCN2A, PSMB7, PSMD1, S1PR3, GPER, P2RX7, PKMYT1, MAP3K5, SIGMAR1, PIP5K1A, MAP3K3, GPA33, SLC29A1, EPAS1, TSG101, SMO, GABRG3, SOST, VKORC1, TUBA1C, RIOK1, TUBB6, ABHD6, TUBB2B, RIOK2, EBPL, ACE2, STK33, DCLK3, EGLN1, CHRNA10, NUAK2, PARP12, SIK2, MYLK2, S1PR5, P2RY12, SLK, TNKS2, TAOK3, BHMT2, HIPK2, HRH4, SRMS, FZD8, TUBB1, EGLN3, SRD5A3, CLK4, TRPV4, RXFP1, NOD2, EML4, GPR35, PDE11A,* |

| | | |
|---|---|---|
| | *BCL2L10, IL23A, RTN4, KCNQ5, TLR9, CYSLTR2, SLC22A11, BMP2K, TDP1, IRAK4, SIRT5, P4HTM, SCN3A, CACNA2D2, SLC5A4, TLR7, MAP3K20, IL20, CISD1, CACNA1I, STK26, RCOR3, EIF2AK4, DHCR7, RPS6KB2, MALT1, STK17A, EPHA6, CHRNA9, PARP2, NPC1L1, TBK1, STEAP1, EGFL7, SCN11A, DAPK2, TNIK, UTS2R, IKZF3, HDAC9, ALK, CELA1, PSMD13, FZD1, TFR2, AURKC, SCN8A, HDAC5, CAMK2A, PDE10A, HPSE, CYSLTR1, CA5B, NISCH, SIK3, MAP3K2, DYRK1B, L3MBTL1, DAGLA, LOXL2, MAP4K5, TNFRSF18, PTGDR2, IRAK3, NCOR2, STK24, PARP3, DNMT3A, MAP3K4, TNFRSF10A, TNFRSF10B, EGFR, REN, ADA, TGFB1, IL2RA, TFRC, MMP1, CD5, MMP7, CSF1, SPP1, IL1R1, TNFRSF1A, IL4R, F2R, CD40, CD38, AXL, CA5A, LTBR, ACVRL1, CD200, SULT1A1, EPHB4, IL10RB, MERTK, CASP8, ANGPT1, NTRK3, PCSK9, CD274, TNFSF13B, F2, PLG, CD14, TLR4, NRP1, MAPK13, ICOSLG, DKK1, ICAM1, FCER2, CA3, RET, GSTP1, PDGFRB, GHR, PTHLH, TDGF1, SELL, ATP1B2, PLA2G2A, MMP9, IDO1, AKR1B1, NQO1, NQO2, CBR1, IFNAR1, CD33, EPHA1, CA6, TNC, EPHB2, CSF2RB, TIE1, FLT4, KDR, SNCA, PTGDS, PRCP, LEPR, VEGFC, IL11RA, GPNMB, RRM2B, LY96, EPHB6, TNFSF12, VEGFA, SELP, MMP12, GFRA1, DLK1, TEK, PDCD1LG2, SLAMF7, F10, MFGE8, IL6R, IL17RA,* | |
| 703 human druggable proteins have evidence for involvement in COVID-19 | *SIGMAR1, DNMT1, SIRT5, BRD4, BRD2, IL17RA, TLR9, TLR7, ESR1, ESR2, CSNK2A1, RET, FLT3, PIK3CD, TOP2A, PIK3R1, AXL, PIK3CB, PIK3CG, EBP, ALK, PIK3R2, CCNT1, CDK1, ROS1, CDK4, CCND1, CCNE1, CDK2, CCND3, PIK3CA, CDK9, CDK6, CDK5, DHCR24, MAPK14, PIK3R5, PIK3R3, ABL1, LMNA, LYN, CHRM4, CHRM5, ADRA2A, PDGFRB, KIT, BCR, FGFR1, DRD2, PDGFRA, FLT1, ADRA2C, CHRM3, IMPDH1, FGFR2, FGFR4, FGFR3, ADRA1D, HTR2A, HTR2C, HRH1, DRD3, KDR, HTR2B, ABL2, HTR6, SMN1, SMN2, PLK4, GAK, NPC1, JAK2, CACNA1F, ROCK2, EGFR, F10, CA1, CA2, NR3C1, ERBB2, TP53, INSR, LCK, FYN, PGR, EIF4E, CSF1R, YES1, IGF1R, ABCB1, HCK, CYP3A4, FGR, RARA, RARB, CHRM1, VDR, CYP2C9, RARG, NQO2, ADRA2B, ATP4A, DRD1, DRD4, PPIB, SLC6A2, AGTR1, SLC6A4, CYP2C19, HTR7, PTGS2, OPRM1, FLT4, THPO, CSK, MTOR, TEC, TXK, PTGIR, RAB9A, BLK, BMX, JAK3, BLM, ESRRG, PPIA, FKBP1A, FKBP1B, CACNA1D, FKBP4, GRIN1, BTK, PPP3CA, DDR1, PPID, KCNH2, STK4, MAP2K5, STK3, GRIN2B, FKBP5, ROCK1, PRPF4B, CAMK2D, CACNA1S, PTK6, CACNA1C, ERBB4, CA9, DDR2, AAK1, PDCD4, MAP3K19, SLC47A1, SRPK1, EBPL, NUAK2, CLK4, TDP1, DHCR7, SRPK3, CAMK2A, RIPK2, IMPDH2, SRC, BMPR1B, AURKA, EPHB6, MAPK13, PRPF4, NUAK1, PAK4, ALDH1A1, RAF1, ATP1A1, ATP1B1, CYP17A1, PRKCB, CHRM2, MET, ARAF, CYP2D6, THRB, PIM1, CYP19A1, CFTR, ATP1A3, ATP1B2, BRAF, CYP11B2, EPHA1, ERBB3, DRD5, CPT2, MAPK3, MARK3, MAPK1, EPHA2, EPHA3, EPHA8, EPHB2, CCND2, AKT1, AKT2, EPHX2, PTGER1, SCN1A, SCN4A, TIE1, MPL, GRIA1, GRIA2, GRIA3, FRK, PTGER2, SYK, MAPK8, MAPK9, GRIA4, MAPKAPK2, CDK8, FNTA, FNTB, CDK7, ATP1A2, PLK1, LIMK2, MAPK12, MAPK10, ATP1B3, FXYD2, EPHB3, EPHA5, EPHB4, EPHB1, EPHA4, SIK1, KCNJ2, MAP3K9, SCN7A, MYT1, TEK, ACVR1, TNK2, CYP24A1, PRKG2, TNK1, NAE1, DYRK1A, ATP1A4, SCN5A, TESK1, MAPK11, GRK1, SCN9A, PKN1, PKN2, MAP3K11, CYP51A1, TNNI3K, MARK2, PIM3, HIPK4, UBA3, MAPK15, GRK7, CPT1B, AURKB, CAMKK2, SCN2A, PKMYT1, RIOK2, TSSK1B, SIK2, SRMS, EML4, SCN3A, MAP3K20, PIM2, EIF2AK4, TBK1, SCN11A, SCN8A, PDE10A, AKT3, SIK3, HCN4, DYRK1B, MAP4K5, SCN10A, RBX1, IKZF1, CUL4A, DDB1, CRBN, IKZF3, KCNK3, RIOK3, XPO1, DCLK1, GRIN2D, TLR3, DAPK3, MAP3K7, CA12, PDE6D, GRIN3B, GUCY1B2, GMNN, STK16, PDE5A, STK10, KCNK2, S1PR2, ESRRB, S1PR4, DHFR, F2, PLG, REN, SERPINC1, C5, TNF, IFNG, IL1B, KLKB1, MMP1, HMGCR, CSF2, MAS1, SHBG, TUBB4A, NTRK1, TYMS, ITGB3, CYP1A2, IL6, TUBB, ADRB2, SERPINA6, NR3C2, ITGA2B, CD14, ADRB1, IL6R, MMP7, HMGB1, LTA4H, ADORA3, TUBA3C, CXCL8, AR, ESRRA, ACE, SLC5A1, ADRB3, INSRR, IL1R1, CSF2RA, VEGFA, ALOX15, SELP, TSHR, PDE6A, FER, IFNAR1, SRD5A1, PDE6G, NFKB1, CYP3A5, S1PR1, CNR1, GART, ACHE, MMP8, PTGS1, CA6, GLRA1, JAK1, CYP3A7, EDNRB, TBXAS1, HRH2, EDNRA, TACR1, DPP4, HTR1D, ADORA2A, ADORA2B, CCN2, TYK2, CD6, AVPR2, OXTR, GNRHR, SRD5A2, SLC5A2, ATIC, GRK4, CSF2RB, CD80, TTK, ADRA1A, ADRA1B, PDE6B, MAP2K2, AVPR1A, OPRD1, OPRK1, SLC19A1, CD86, AKR1C3, CA7, MMP13, AVPR1B, MTNR1A, IFNAR2, MTNR1B, CLK1, GSK3B, AGTR2, PDE6C, CYP2J2, CCR5, RPS6KA3, DAPK1, PRKAA2, KCNK10, TUBA1B, TUBA4A, TUBB4B, PIP4K2B, SRPK2, CDK16, CDK17, SLC6A3, DHODH, GUCY1B1, MAP2K1, TOP2B, AOX1, IL10RB, ITK, MERTK, GRIN2A, FAP, PRKAA1, TUBB3, CAMK2G, BMPR2, TUBB2A, PDE6H, PRKG1, PTK2B,* | PMID: 33837377 |

| | |
|---|---|
| | *MELK, GRIN2C, PDCD1, RPS6KA2, SF3B3, STK11, NTRK3, IL17A, NTRK2, HIF1A, TUBB8, LRRK2, TUBA3E, AC091230.1, NPSR1, TUBA1A, KCNK18, IFNLR1, MINK1, GRIN3A, MAP4K1, S1PR3, GPER1, HSD17B10, PIP5K1A, MAP3K3, SLC29A1, TUBA1C, RIOK1, TUBB6, TUBB2B, DCLK3, S1PR5, P2RY12, SLK, TUBB1, SRD5A3, CYP3A43, KCNK9, CYSLTR2, BMP2K, SLC5A4, STK26, DAPK2, TNIK, CYSLTR1, MAP3K2, PTGDR2, GLA, COMT, BRD3, F8, IL2RA, FGB, VWF, CD2, CD3E, ALOX5, MS4A1, ITGA4, PKM, IL2RB, GLUL, IFNGR1, NQO1, ALOX12, ITGAL, CXCR1, CXCR2, F2R, ITGB7, HSD3B2, PDE4A, IL12B, CD52, PPARG, IFNGR2, CCR2, RECQL, CXCR4, ADAM17, PPAT, PDE4B, PDE4C, AC008397.2, PDE4D, PTGDR, GPR17, KEAP1, PDE3A, IL11RA, TXNRD1, CA13, CSF3R, VKORC1, HRH4, NOD2, GPR35, IL23A, TNFSF13B, CA5B, HRH3, ACE2, TLR4, PSMD11, CDC7, PSMD14, STK25, TNFRSF10B, PSMA7, PDE8A, CTSV, NCOR1, ULK1, IDH1, TNFSF13, CHEK2, PLAT, ADA, IL1A, TTR, AFP, CXCL10, TFRC, APOB, TK1, CD74, APP, ALDH2, IL4, MPO, ITGB1, CEACAM5, NPM1, ITGAV, H1-0, HSP90AA1, HSP90AB1, ITGA5, VIM, GSTP1, CSF1, PARP1, CALM1, BCL2, SPP1, HSPA8, TOP1, ADH5, CCL2, EEF2, PLA2G2A, AKR1B1, FOLR1, CD19, PHKG2, MUC1, CD44, EPCAM, PRKCA, PRKACA, TNNI3, DDC, IL10, PRKACG, PRKACB, RPS6KB1, RRM1, PSMA2, PSMA3, PSMA4, PSMB9, PSMA5, PSMB5, BDKRB2, CPS1, RRM2, KIF5B, GUCY1A2, ABCC1, SOAT1, BSG, PSMC2, SNCA, GGCX, IL6ST, HTT, GLP1R, GPD2, PSMC4, BDKRB1, PSMD8, IDH2, PSMB3, PSEN1, GSK3A, PSMD7, KIF11, NR1H2, GFER, HDAC4, PSMA6, PSMC1, PSMC5, HBA2, HBA1, PRKDC, NFKB2, KMT2A, PTK2, PRKCZ, PRKCD, BCL2L1, MCL1, MFGE8, PAK2, PSMD2, MAP3K1, HDAC1, IL18, PSMD6, PDK1, RPS6KA1, SHH, MYLK, ADRM1, CCDC6, BCL2A1, TMEM97, RRM2B, DPP9, MAPKAPK5, ABHD12, PCSK9, TRPV1, HDAC7, MUC16, HDAC2, BCL2L2, BCL2L2-PABPN1, PSMD1, GPA33, TSG101, IL22, RTN4, RPS6KB2, STK17A, PSMD13, NCOR2, STK24* |

#### Supplemental Table S14. Functional enrichment analysis of 190 up-DEGs associated with severe COVID-19 based on the Reactome database

| Description | Size | Ratio | P Value | FDR |
|---|---|---|---|---|
| Immune System | 1997 | 2.56 | 1.65E-14 | 2.86E-11 |
| Innate Immune System | 1053 | 3.24 | 1.79E-12 | 1.55E-09 |
| Neutrophil degranulation | 479 | 4.07 | 3.39E-09 | 1.95E-06 |
| Negative regulators of DDX58/IFIH1 signaling | 34 | 16.71 | 1.49E-07 | 6.45E-05 |
| Cellular responses to external stimuli | 503 | 3.23 | 3.15E-06 | 1.09E-03 |
| DDX58/IFIH1-mediated induction of interferon-alpha/beta | 78 | 8.33 | 4.86E-06 | 1.40E-03 |
| Interleukin-4 and Interleukin-13 signaling | 108 | 6.77 | 6.92E-06 | 1.71E-03 |
| Adaptive Immune System | 756 | 2.58 | 1.45E-05 | 3.13E-03 |
| Interferon alpha/beta signaling | 69 | 8.24 | 2.09E-05 | 4.02E-03 |
| Cellular responses to stress | 426 | 3.05 | 6.63E-05 | 1.15E-02 |
| Regulation of TLR by endogenous ligand | 19 | 17.09 | 7.38E-05 | 1.16E-02 |
| Cytokine Signaling in Immune system | 688 | 2.48 | 9.46E-05 | 1.36E-02 |
| Interferon gamma signaling | 92 | 6.18 | 1.34E-04 | 1.79E-02 |

| | | | | |
|---|---|---|---|---|
| Interferon Signaling | 197 | 4.12 | 1.56E-04 | 1.93E-02 |
| Class I MHC mediated antigen processing & presentation | 371 | 3.06 | 1.86E-04 | 2.14E-02 |
| Antigen processing-Cross presentation | 99 | 5.74 | 2.13E-04 | 2.30E-02 |
| IRF3-mediated induction of type I IFN | 13 | 18.74 | 4.77E-04 | 4.85E-02 |

**Supplemental Table S15. Disease-based enrichment analysis of 190 up-DEGs associated with severe COVID-19 among *CCR1*+ CD16+monocytes based on the GLAD4U database.**

| Disease terms | Size | Expect | Enrichment Ratio | P Value | FDR |
|---|---|---|---|---|---|
| Infection | 643 | 5.19 | 5.40 | 2.90E-13 | 7.88E-10 |
| Virus Diseases | 580 | 4.68 | 5.56 | 1.17E-12 | 1.59E-09 |
| Inflammation | 565 | 4.56 | 5.48 | 4.42E-12 | 4.01E-09 |
| Hepatitis | 253 | 2.04 | 7.84 | 2.45E-10 | 1.67E-07 |
| Immune System Diseases | 806 | 6.50 | 3.84 | 7.35E-09 | 4.00E-06 |
| Necrosis | 371 | 2.99 | 5.35 | 5.79E-08 | 2.62E-05 |
| Hyperoxia | 33 | 0.27 | 22.54 | 2.32E-07 | 9.03E-05 |
| HIV | 862 | 6.95 | 3.31 | 4.60E-07 | 1.37E-04 |
| Respiratory Tract Infections | 281 | 2.27 | 5.73 | 4.85E-07 | 1.37E-04 |
| virological response | 282 | 2.28 | 5.71 | 5.05E-07 | 1.37E-04 |
| Sexually Transmitted Diseases | 496 | 4.00 | 4.25 | 5.76E-07 | 1.42E-04 |
| Encephalitis, Viral | 66 | 0.53 | 13.15 | 1.02E-06 | 2.31E-04 |
| Bacterial Infections | 260 | 2.10 | 5.72 | 1.39E-06 | 2.91E-04 |
| Retroviridae Infections | 494 | 3.99 | 4.01 | 2.62E-06 | 4.87E-04 |
| HIV Infections | 495 | 3.99 | 4.01 | 2.69E-06 | 4.87E-04 |
| Lentivirus Infections | 498 | 4.02 | 3.98 | 2.90E-06 | 4.93E-04 |
| Hepatitis B | 190 | 1.53 | 6.52 | 3.39E-06 | 5.42E-04 |
| Mouth Diseases | 246 | 1.98 | 5.54 | 5.25E-06 | 7.93E-04 |
| Reperfusion Injury | 86 | 0.69 | 10.09 | 6.15E-06 | 8.80E-04 |
| Encephalitis | 89 | 0.72 | 9.75 | 7.73E-06 | 1.05E-03 |
| West Nile Fever | 35 | 0.28 | 17.71 | 8.58E-06 | 1.11E-03 |

| | | | | | |
|---|---|---|---|---|---|
| Immunologic Deficiency Syndromes | 500 | 4.03 | 3.72 | 1.36E-05 | 1.66E-03 |
| Rhinitis | 135 | 1.09 | 7.35 | 1.40E-05 | 1.66E-03 |
| Tumor Virus Infections | 186 | 1.50 | 6.00 | 2.07E-05 | 2.35E-03 |
| Psoriasis | 243 | 1.96 | 5.10 | 2.90E-05 | 3.16E-03 |
| Autoimmune Diseases | 546 | 4.40 | 3.41 | 3.76E-05 | 3.93E-03 |
| Arthritis, Reactive | 78 | 0.63 | 9.54 | 4.00E-05 | 3.99E-03 |
| Skin and Connective Tissue Diseases | 617 | 4.98 | 3.21 | 4.11E-05 | 3.99E-03 |
| Skin Diseases, Viral | 85 | 0.69 | 8.75 | 6.50E-05 | 6.07E-03 |
| Stress | 643 | 5.19 | 3.08 | 6.70E-05 | 6.07E-03 |
| Connective Tissue Diseases | 392 | 3.16 | 3.79 | 8.40E-05 | 7.32E-03 |
| Hepatitis, Chronic | 174 | 1.40 | 5.70 | 8.61E-05 | 7.32E-03 |
| Liver Neoplasms | 396 | 3.19 | 3.76 | 9.25E-05 | 7.61E-03 |
| Periodontitis | 91 | 0.73 | 8.17 | 9.52E-05 | 7.61E-03 |
| Respiratory Syncytial Virus Infections | 229 | 1.85 | 4.87 | 1.04E-04 | 8.09E-03 |
| Gram-Positive Bacterial Infections | 181 | 1.46 | 5.48 | 1.13E-04 | 8.56E-03 |
| Periodontal Diseases | 96 | 0.77 | 7.75 | 1.28E-04 | 9.41E-03 |
| Dermatitis, Atopic | 140 | 1.13 | 6.20 | 1.43E-04 | 1.02E-02 |
| Dermatomyositis | 34 | 0.27 | 14.58 | 1.57E-04 | 1.09E-02 |
| Hepatitis C | 195 | 1.57 | 5.09 | 1.89E-04 | 1.27E-02 |
| Lipidoses | 67 | 0.54 | 9.25 | 2.07E-04 | 1.31E-02 |
| Pneumonia | 109 | 0.88 | 6.82 | 2.57E-04 | 1.45E-02 |
| Adenocarcinoma | 522 | 4.21 | 3.09 | 3.27E-04 | 1.81E-02 |
| Chondrodysplasia Punctata | 83 | 0.67 | 7.47 | 5.62E-04 | 2.88E-02 |
| Lupus erythematosus | 294 | 2.37 | 3.79 | 6.56E-04 | 3.08E-02 |
| Leukemia | 565 | 4.56 | 2.85 | 6.87E-04 | 3.11E-02 |
| Pharyngeal Neoplasms | 137 | 1.11 | 5.43 | 8.66E-04 | 3.62E-02 |

| | | | | | |
|---|---|---|---|---|---|
| Frostbite | 6 | 0.05 | 41.32 | 9.50E-04 | 3.64E-02 |
| Actinomycetales Infections | 145 | 1.17 | 5.13 | 1.16E-03 | 4.00E-02 |
| Measles | 62 | 0.50 | 8.00 | 1.58E-03 | 4.73E-02 |

**Supplemental Table S16. 190 up-DEGs associated with severe COVID-19 among *CCR1*+ CD16+monocytes matched in druggable gene categories based on the DGIdb resource**

| Druggable Gene Category | Matching Gene Count | Matching Gene(s) |
|---|---|---|
| Druggable genome | 65 | *AHR, APOBEC3A, ASGR1, C1QB, CD14, CD163, CD300E, CD36, CD53, CD63, CD84, CD99, CDKN1A, CTSA, CTSD, CTSL, CXCL8, CYP1B1, FKBP5, FOLR3, FPR1, FPR2, GLUL, HBEGF, HIF1A, HRH2, HSPA5, ICAM1, IFITM1, IGFBP7, IL4R, ISG15, LAIR1, LDHA, LGALS3BP, MGST1, NAMPT, PARP9, PGD, PIM1, PIM3, PLBD1, PLSCR1, PPIF, PSMA1, PXK, RNASE2, S100A12, S100A8, S100A9, SELL, SERPING1, SGK1, SIGLEC1, SLC12A7, SLC25A37, SLC2A3, SMPDL3A, TCN2, TNFAIP2, TNFSF10, TXN, TXNRD2, UPP1, VCAN* |
| Enzyme | 35 | *AHR, APOBEC3A, ATG12, BLVRB, CTSA, CTSD, CTSL, CYP1B1, DYNLL1, GLRX, GLUL, GM2A, HIF1A, HSPA5, IFIH1, ISG15, KRTCAP2, NCF1, PARP9, PGD, PIM1, PIM3, PXK, S100A8, S100A9, SGK1, SMPDL3A, SUMO3, SUPT5H, TNFAIP3, UBE2B, UBE2K, UBE2L3, UBE2V2, UPP1* |
| Kinase | 21 | *CCNL1, CD163, CD300E, CD63, CDKN1A, GADD45GIP1, HRH2, IFIH1, IRF3, MPLKIP, PIM1, PIM3, PXK, RGCC, S100A12, S100A8, S100A9, SGK1, SOCS3, STING1, TNFAIP3* |

| | | |
|---|---|---|
| Clinically actionable | 18 | *BCL3, CAMTA1, CD36, CDKN1A, CYP1B1, ETV6, HIF1A, H2BC12, IRF2, MAML2, NOP10, PER1, PIM1, PRCC, SGK1, SOCS3, STING1, TNFAIP3* |
| Transcription factor | 14 | *AHR, ATF3, CAMTA1, CEBPB, CEBPD, ETV6, HIF1A, ID1, IFI16, IRF2, IRF3, PER1, SCAND1, SGK1* |
| Cell surface | 8 | *C1QB, CD36, CD53, CD63, HBEGF, HSPA5, ICAM1, VAMP5* |
| Transporter | 8 | *CD36, EMB, HIF1A, HSPA5, SGK1, SLC12A7, SLC25A37, SLC2A3* |
| Transcription factor binding | 7 | *AHR, BCL3, HIF1A, ID1, IFI16, PIM1, STING1* |
| Drug resistance | 6 | *CDKN1A, ICAM1, LDHA, MGST1, SGK1, UBE2B* |
| External side of plasma membrane | 6 | *CD14, CD163, CD36, FLOT1, ICAM1, SLC12A7* |
| Protease | 5 | *CTSA, CTSD, CTSL, PSMA1, TNFAIP3* |
| Protease inhibitor | 4 | *APLP2, HSPA5, NAIP, SERPING1* |
| Serine threonine kinase | 4 | *PIM1, PIM3, PXK, SGK1* |
| G protein coupled receptor | 3 | *FPR1, FPR2, HRH2* |
| Nuclear hormone receptor | 3 | *AHR, GADD45GIP1, PER1* |

**Supplemental Table S17. Highly-expressed inflammatory and cytokine genes among *ABO*+ megakaryocytes**

| Gene name | T score | Fold change | P value | FDR |
|---|---|---|---|---|
| *ADORA2B* | 2.80 | 2.27 | 5.29E-03 | 2.227E-02 |
| *ADRM1* | 3.02 | 1.66 | 2.72E-03 | 1.301E-02 |
| *AHR* | 2.52 | 2.25 | 1.22E-02 | 4.349E-02 |
| *GNA15* | 3.49 | 1.82 | 5.35E-04 | 3.461E-03 |
| *IRAK2* | 3.20 | 2.85 | 1.48E-03 | 7.919E-03 |
| *KCNA3* | 3.29 | 1.89 | 1.10E-03 | 6.179E-03 |

| | | | | |
|---|---|---|---|---|
| *PDGFA* | 4.26 | 1.71 | 2.53E-05 | 2.666E-04 |
| *PTGIR* | 4.05 | 1.56 | 6.18E-05 | 5.671E-04 |
| *SPHK1* | 3.71 | 1.57 | 2.36E-04 | 1.745E-03 |
| *BMP6* | 2.90 | 2.44 | 3.96E-03 | 1.766E-02 |
| *TNFSF4* | 4.81 | 2.67 | 2.18E-06 | 3.138E-05 |

**Supplemental Table S18. Pathway enrichment analysis of 424 highly-expressed genes among *ABO*+ megakaryocytes.**

| **Pathway name** | **Gene size** | **Enrichment ratio** | **P Value** | **FDR** |
|---|---|---|---|---|
| Systemic lupus erythematosus | 133 | 4.79 | 4.55E-07 | 1.48E-04 |
| Alcoholism | 180 | 4.01 | 9.82E-07 | 1.60E-04 |
| Platelet activation | 123 | 3.80 | 1.42E-04 | 1.55E-02 |
| SNARE interactions in vesicular transport | 34 | 6.24 | 1.10E-03 | 8.94E-02 |
| Endocytosis | 244 | 2.43 | 1.80E-03 | 1.17E-01 |
| Viral carcinogenesis | 201 | 2.53 | 2.74E-03 | 1.28E-01 |
| VEGF signaling pathway | 59 | 4.32 | 2.52E-03 | 1.28E-01 |
| Necroptosis | 162 | 2.62 | 4.83E-03 | 1.61E-01 |
| Mitophagy | 65 | 3.92 | 4.11E-03 | 1.61E-01 |
| Chemokine signaling pathway | 189 | 2.47 | 4.95E-03 | 1.61E-01 |
| Transcriptional misregulation in cancer | 186 | 2.28 | 1.23E-02 | 3.65E-01 |
| Axon guidance | 175 | 2.18 | 2.23E-02 | 6.05E-01 |
| Bacterial invasion of epithelial cells | 74 | 2.87 | 2.99E-02 | 6.51E-01 |
| Regulation of actin cytoskeleton | 213 | 1.99 | 2.88E-02 | 6.51E-01 |
| Adherens junction | 72 | 2.95 | 2.70E-02 | 6.51E-01 |
| Human cytomegalovirus infection | 225 | 1.89 | 3.97E-02 | 8.08E-01 |
| Proteoglycans in cancer | 201 | 1.90 | 4.77E-02 | 8.64E-01 |
| Tight junction | 170 | 2.00 | 4.73E-02 | 8.64E-01 |

**Supplemental Table S19. Disease-term enrichment analysis of 35 up-DEGs associated with severe COVID-19 among *ABO*+ megakaryocytes based on the GLAD4U database**

| **Term ID** | **Disease-terms** | **Size** | **Expect** | **Enrichment Ratio** | **P Value** |
|---|---|---|---|---|---|
| PA443490 | Bernard-Soulier Syndrome | 9 | 0.01 | 140.31 | 8.69E-05 |

| | | | | | |
|---|---|---|---|---|---|
| PA445644 | Shock | 367 | 0.58 | 8.60 | 2.59E-04 |
| PA443842 | Death | 417 | 0.66 | 7.57 | 4.65E-04 |
| PA445846 | Thrombocytopenia | 96 | 0.15 | 19.73 | 4.67E-04 |
| PA443382 | Anoxia | 251 | 0.40 | 10.06 | 6.36E-04 |
| PA445457 | Pterygium | 25 | 0.04 | 50.51 | 7.13E-04 |
| PA165108957 | Venous ulcer of leg | 26 | 0.04 | 48.57 | 7.71E-04 |
| PA166048906 | Blood Platelet Disorders | 118 | 0.19 | 16.05 | 8.51E-04 |
| PA444668 | Keratitis | 31 | 0.05 | 40.74 | 1.10E-03 |
| PA166129556 | disease activity score 28 joint in rheumatoid arthritis | 31 | 0.05 | 40.74 | 1.10E-03 |
| PA443429 | Arteritis | 34 | 0.05 | 37.14 | 1.32E-03 |
| PA443882 | Dermatomyositis | 34 | 0.05 | 37.14 | 1.32E-03 |
| PA445850 | Thrombosis | 141 | 0.22 | 13.43 | 1.42E-03 |
| PA166123766 | platelet aggregation | 153 | 0.24 | 12.38 | 1.80E-03 |
| PA446220 | Abdominal Pain | 43 | 0.07 | 29.37 | 2.11E-03 |
| PA445793 | Synovitis | 47 | 0.07 | 26.87 | 2.51E-03 |
| PA443433 | Arthritis, Juvenile Rheumatoid | 48 | 0.08 | 26.31 | 2.62E-03 |
| PA445051 | Necrosis | 371 | 0.59 | 6.81 | 2.68E-03 |
| PA446477 | Polymyositis | 49 | 0.08 | 25.77 | 2.73E-03 |
| PA166048887 | Irritable Bowel Syndrome | 50 | 0.08 | 25.26 | 2.84E-03 |
| PA445752 | Stress | 643 | 1.02 | 4.91 | 3.17E-03 |
| PA445593 | Sarcoidosis | 56 | 0.09 | 22.55 | 3.55E-03 |
| PA444034 | Endocarditis | 67 | 0.11 | 18.85 | 5.04E-03 |
| PA443829 | Cystic Fibrosis | 223 | 0.35 | 8.49 | 5.20E-03 |
| PA445044 | Nasal Polyps | 75 | 0.12 | 16.84 | 6.27E-03 |
| PA444035 | Endocarditis, Bacterial | 76 | 0.12 | 16.62 | 6.44E-03 |
| PA445512 | Arthritis, Reactive | 78 | 0.12 | 16.19 | 6.77E-03 |
| PA446020 | Varicose Veins | 79 | 0.13 | 15.99 | 6.94E-03 |

| | | | | | |
|---|---|---|---|---|---|
| PA165108622 | Drug interaction with drug | 494 | 0.78 | 5.11 | 7.38E-03 |
| PA165374639 | suicide | 83 | 0.13 | 15.22 | 7.63E-03 |
| PA446303 | Tooth Loss | 84 | 0.13 | 15.03 | 7.81E-03 |
| PA444942 | Methemoglobinemia | 5 | 0.01 | 126.28 | 7.89E-03 |
| PA447296 | nondiabetic proteinuric nephropathy | 5 | 0.01 | 126.28 | 7.89E-03 |
| PA443474 | Bacterial Infections | 260 | 0.41 | 7.29 | 7.93E-03 |
| PA445672 | Sinusitis | 89 | 0.14 | 14.19 | 8.73E-03 |
| PA443690 | Charcot-Marie-Tooth Disease | 95 | 0.15 | 13.29 | 9.90E-03 |
| PA445382 | Polyps | 95 | 0.15 | 13.29 | 9.90E-03 |
| PA445296 | Periodontal Diseases | 96 | 0.15 | 13.15 | 1.01E-02 |
| PA447230 | HIV | 862 | 1.37 | 3.66 | 1.08E-02 |
| PA165108641 | Acatalasia | 7 | 0.01 | 90.20 | 1.10E-02 |
| PA165108334 | Ulnar neuropathy | 8 | 0.01 | 78.93 | 1.26E-02 |
| PA446833 | Pouchitis | 8 | 0.01 | 78.93 | 1.26E-02 |
| PA445355 | Pneumonia | 109 | 0.17 | 11.59 | 1.29E-02 |
| PA446540 | Cytomegalovirus Retinitis | 9 | 0.01 | 70.16 | 1.42E-02 |
| PA447149 | Chills | 9 | 0.01 | 70.16 | 1.42E-02 |
| PA444231 | Fructose Intolerance | 10 | 0.02 | 63.14 | 1.57E-02 |
| PA443820 | Cryptococcosis | 11 | 0.02 | 57.40 | 1.73E-02 |

**Supplemental Table S20. 35 up-DEGs significantly associated with severe COVID-19 among *ABO*+ megakaryocytes matched in druggable gene categories**

| Druggable Gene Category | Matching Gene Count | Matching Gene(s) |
|---|---|---|
| Druggable genome | 11 | *ACP1, CFLAR, CYB5R3, FKBP8, HSPB1, MPIG6B, P2RX1, S100A8, S100A9, SMOX , TUBB1* |
| Enzyme | 9 | *ACP1, ATP6V1E1, CYB5R3, ENO1, HK1, S100A8, S100A9, SMOX, SUPT4H1* |
| Kinase | 5 | *HCST, HK1, S100A8, S100A9, SKP1* |

| | | |
|---|---|---|
| Transporter | 4 | *ATF4, ATP6V1E1, SEC14L1, SLC44A2* |
| Cell surface | 2 | *ENO1, HCST* |
| Clinically actionable | 1 | *MYH9* |
| Drug resistance | 1 | *CFLAR* |
| External side of plasma membrane | 1 | *P2RX1* |
| Ion channel | 1 | *P2RX1* |
| Protease | 1 | *CFLAR* |
| Protein phosphatase | 1 | *ACP1* |
| Transcription factor | 1 | *ATF4* |

**Supplemental Table S21. Pathway enrichment analysis of 158 highly-expressed genes among *CXCR6*+ memory CD8+T cells.**

| Pathway name | Size | Enrichment Ratio | P Value | FDR |
|---|---|---|---|---|
| Cytokine-cytokine receptor interaction | 294 | 5.65 | 1.24E-08 | 4.04E-06 |
| Inflammatory bowel disease (IBD) | 65 | 9.58 | 3.43E-05 | 5.58E-03 |
| NF-kappa B signaling pathway | 95 | 5.46 | 2.14E-03 | 2.32E-01 |
| Toxoplasmosis | 113 | 4.59 | 4.54E-03 | 2.85E-01 |
| Necroptosis | 162 | 3.84 | 4.55E-03 | 2.85E-01 |
| Allograft rejection | 38 | 8.19 | 5.69E-03 | 2.85E-01 |
| Pyruvate metabolism | 39 | 7.98 | 6.12E-03 | 2.85E-01 |
| ABC transporters | 44 | 7.07 | 8.57E-03 | 3.49E-01 |
| Apoptosis | 136 | 3.81 | 9.81E-03 | 3.55E-01 |
| Intestinal immune network for IgA production | 49 | 6.35 | 1.15E-02 | 3.76E-01 |
| Autoimmune thyroid disease | 53 | 5.87 | 1.43E-02 | 4.23E-01 |
| Chagas disease (American trypanosomiasis) | 102 | 4.07 | 1.66E-02 | 4.51E-01 |
| Human cytomegalovirus infection | 225 | 2.77 | 2.09E-02 | 5.25E-01 |
| Kaposi sarcoma-associated herpesvirus infection | 186 | 2.79 | 3.32E-02 | 6.83E-01 |
| Citrate cycle (TCA cycle) | 30 | 6.92 | 3.35E-02 | 6.83E-01 |

| | | | | |
|---|---|---|---|---|
| Chemokine signaling pathway | 189 | 2.74 | 3.52E-02 | 6.83E-01 |
| Circadian rhythm | 31 | 6.69 | 3.56E-02 | 6.83E-01 |
| Measles | 132 | 3.14 | 3.81E-02 | 6.85E-01 |
| Apoptosis | 33 | 6.29 | 3.99E-02 | 6.85E-01 |
| Phenylalanine, tyrosine and tryptophan biosynthesis | 5 | 20.75 | 4.73E-02 | 7.71E-01 |

**Supplemental Table S22. GO-terms enrichment analysis of 108 up-DEGs associated with COVID-19 among *CXCR6*+ memory CD8+T cells.**

| GO-terms | Size | Enrichment ratio | P Value | FDR |
|---|---|---|---|---|
| Immune response-activating cell surface receptor signaling pathway | 300 | 6.79 | 6.30E-07 | 2.83E-03 |
| Positive regulation of immune system process | 979 | 3.59 | 9.22E-07 | 2.83E-03 |
| Immune response-activating signal transduction | 454 | 5.30 | 9.34E-07 | 2.83E-03 |
| Immune response-regulating cell surface receptor signaling pathway | 330 | 6.17 | 1.60E-06 | 3.54E-03 |
| Immune response-regulating signaling pathway | 485 | 4.96 | 1.95E-06 | 3.54E-03 |
| Post-translational protein modification | 360 | 5.66 | 3.71E-06 | 4.83E-03 |
| Regulation of immune system process | 1400 | 2.91 | 3.72E-06 | 4.83E-03 |
| Positive regulation of immune response | 706 | 3.93 | 5.21E-06 | 5.63E-03 |
| Activation of immune response | 534 | 4.51 | 5.58E-06 | 5.63E-03 |
| Leukocyte differentiation | 496 | 4.48 | 1.41E-05 | 1.28E-02 |
| Hemopoiesis | 791 | 3.51 | 2.01E-05 | 1.66E-02 |
| Hematopoietic or lymphoid organ development | 833 | 3.33 | 3.67E-05 | 2.78E-02 |
| Regulation of hemopoiesis | 389 | 4.76 | 4.64E-05 | 3.25E-02 |
| Immune response-regulating cell surface receptor signaling pathway involved in phagocytosis | 76 | 12.18 | 5.61E-05 | 3.28E-02 |
| Fc-gamma receptor signaling pathway involved in phagocytosis | 76 | 12.18 | 5.61E-05 | 3.28E-02 |
| Immune response | 1919 | 2.32 | 5.76E-05 | 3.28E-02 |
| Antigen receptor-mediated signaling pathway | 184 | 7.04 | 6.16E-05 | 3.29E-02 |
| Fc-gamma receptor signaling pathway | 79 | 11.72 | 6.75E-05 | 3.32E-02 |
| Immune system development | 881 | 3.15 | 6.94E-05 | 3.32E-02 |
| Fc receptor mediated stimulatory signaling pathway | 81 | 11.43 | 7.61E-05 | 3.46E-02 |

| | | | | |
|---|---|---|---|---|
| Regulation of leukocyte differentiation | 263 | 5.63 | 8.77E-05 | 3.80E-02 |
| Regulation of immune response | 909 | 3.06 | 9.86E-05 | 4.07E-02 |
